# Post-pandemic mortality patterns and COVID-19 burden considering multiple death causes

**DOI:** 10.1101/2025.09.01.25334892

**Authors:** Uwe Riedmann, Michael Levitt, Stefan Pilz, John PA Ioannidis

**Affiliations:** Department of Internal Medicine, Division of Endocrinology and Diabetology, Medical University of Graz, 8036 Graz, Austria.; Department of Structural Biology, Stanford University, Stanford, CA, USA; Departments of Medicine, Epidemiology and Population Health, and Biomedical Data Science, and Meta-Research Innovation Center at Stanford (METRICS), Stanford University, Stanford, CA, USA

**Keywords:** COVID-19, Mortality patterns, Vaccination, Number needed to vaccinate, Multiple cause mortality analysis

## Abstract

**Background:** Post-pandemic mortality rates can explore the residual COVID-19 burden and changes in other causes of death. Considering weighted multiple causes of death from death certificates (underlying and others) may help compare post-versus pre-pandemic mortality patterns, while potentially reducing the impact of cause misattribution. Estimates of post-pandemic impact are critical also for proper continuing public health policies (e.g. vaccinations).

**Methods:** We retrospectively analyse national all-cause mortality rate ratios between 2024 and pre-pandemic years (2017-2019) for sex-stratified 10-year age groups in Austria. In weighted analyses, the underlying death cause was weighted 50% and other causes shared the remaining 50%. Sensitivity analyses explored different weightings. Cause-specific weightings were also compared between 2024 and 2019.

**Results:** Despite 1,212 reported COVID-19 deaths in 2024, all-cause mortality rates were equal or lower in 2024 compared to 2019 in all strata at risk from COVID-19 (i.e., aged 60 years and over). All-cause mortality rates in 2024 were higher than in 2019 in adolescent and young adult strata. The ratio of weighted over unweighted COVID-19 death rates was 0.51-0.58 for age strata 60 years and older and even lower in sensitivity analyses, indicating that COVID-19 deaths were possibly overestimated.

**Conclusions:** Post-pandemic COVID-19 deaths had no visible impact on mortality patterns in Austria and were possibly overcounted. Increased post-pandemic mortality patterns in the young are particularly worrisome.

## INTRODUCTION

Evaluation of the continued, post-pandemic impact of COVID-19 is challenging. The effort to thoroughly document SARS-CoV-2 infections through intensive testing has been discontinued worldwide. Nevertheless, indirect ways may be used to evaluate the impact of SARS-CoV2 in the post-pandemic period. One such approach is based on all-cause mortality data. During the COVID-19 pandemic, most countries reported excess mortality compared to pre-pandemic years [1,2], with large differences across countries [3]. However, excess death calculations have limitations [3,4], especially in western countries where decrease in mortality trends slowed and sometimes even reversed in the years before the pandemic [5]. A comparison of mortality rates in different age and sex strata between post- and pre-pandemic years may provide some insight into the current disease burden of COVID-19 as well as any potential long-term consequences of the pandemic. If SARS-CoV-2 infections continued to have a serious health impact in 2024, strata that are disproportionately at risk of COVID-19 death (e.g., 60+ year olds) should have higher mortality rates compared to the pre-pandemic years. Conversely, other long-term consequences of the pandemic and of the pandemic response may be reflected in increased mortality rates in younger populations.

COVID-19 disease burden may also be evaluated directly by investigating reported COVID-19 deaths. However, this approach depends heavily on how deaths are reported and classified. In many cases, individuals who die with a positive SARS-CoV-2 test may be recorded as COVID-19 deaths, even when other conditions are more relevant [6–8]. Differentiating between deaths caused by COVID-19 and deaths where COVID-19 was merely yet another co-existing factor is crucial for understanding the true burden of the disease, particularly in older or chronically ill populations [9].

Accurate estimates of post-pandemic COVID-19 impact are also essential for proper formulation of COVID-19 vaccination recommendations. Current COVID-19 vaccination recommendations vary starkly across western countries (e.g. age recommendation for yearly vaccination in 2024: 12 years and older in Austria; 65 years and older in Denmark) [10,11]. The potential benefits of continued COVID-19 vaccinations have come under scrutiny in the context of widespread prior SARS-CoV-2 exposure and unclear vaccine effectiveness (VE) in previously infected individuals [12–16]. Against this background, quantification of post pandemic impact is crucial to inform vaccination policy.

In this study, we assess whether age- and sex-adjusted mortality in Austria has returned to pre-pandemic levels (or lower) by comparing all-cause mortality rates in 2024 with the average rates from 2017 to 2019. Furthermore, we apply a weighted death count methodology to estimate a potentially more accurate contribution of COVID-19 to total mortality, accounting for co-occurring causes of death and comparing death rates from different causes from 2019 to 2024.

## METHODS

### Study design and data description

This is a retrospective study using national data from Austria. We compared mortality rates between pre- and post-COVID-19 pandemic time periods and analysed 2024 death certificates to estimate the potential contribution of COVID-19 deaths to total mortality when both underlying and other contributing causes are considered. Population and mortality data were publicly available [17–19]. Data on death certificates was acquired from Statistics Austria [20].

We followed the Strengthening the Reporting of Observational Studies in Epidemiology (STROBE) checklist (Table S1). The study was approved by the ethics committee at the Medical University of Graz (no. 33-144 ex 20/21). The statistical analyses were conducted using R (version 4.4.2) [21].

### Analyses

#### Pre- and post-pandemic mortality

We compared mortality rates in 2024 with the average rates observed in the three years preceding the pandemic (2017–2019). Specifically, we calculated age- and sex-stratified mortality rates in each 10-year age and sex (binary) stratum (up to 90+ year olds) [17–19]. For each stratum, we then computed the ratio of the 2024 mortality rate to the average mortality rate across 2017–2019, with 95% confidence intervals derived via the delta method on the log scale. Ratios above 1.00 in age groups that exhibit deaths due to COVID-19 (practically those 60+ year old) suggest higher mortality in 2024 relative to the pre-pandemic years and may indicate a persistent COVID-19 disease burden, whereas ratios below 1.00 would argue against this.

One may argue whether in the absence of the COVID-19 pandemic, mortality rates might have decreased (and life expectancy increased) rather than simply remain stable. Long-term trends in high-income countries (including Austria) showed remarkable improvements in life expectancy over many decades, but the rate of improvement declined markedly (and in some countries life expectancy even deteriorated) in the decade preceding the pandemic [5]. In Austria specifically, life expectancy changed minimally by 0.41 years (from 81.49 years to 81.90 years) between 2014 and 2019 (as compared to 1.16 years in 2009-2014, 1.15 years in 2004-2009 and 1.30 years in 1999-2004) [22]. To account for this uncertainty, we also considered a sensitivity analysis interpretation using the value of 0.929 (based on the extrapolation of the age-standardised mortality rate ratio between 2019 and the average of 2012-2014) instead of 1.00 for the mortality ratio in 2024 versus 2017-2019 as an optimistic “expected” value under an assumption of continued improvements absent the pandemic without the deceleration in improvement rates that had been seen in pre-pandemic years.

#### Weighted death counts

To address possible misattribution of death causes, we used death certificate data to calculate the weighted death count, which encompasses both the prevalence of a cause mentioned as the underlying cause of death (UC) as well other mentioned causes (OC) on the death certificate [9,23]. This is based on a previously published methodology which used contributing causes (CC) instead of all other mentions [9,23]. However, Austrian death certificate data states the UC and all other causes mentioned on the death certificates, but not whether causes are immediate, intermediate, or contributing causes [9]. We tried to address this issue in the sensitivity analyses. To calculate weighted estimates, we weighed both the UC and all OC mentions equally (50% UC; 50% all OCs). When no OC was reported, UC was weighted with 100%. The method includes all mentioned causes while addressing the issue with the classic “any mention” method, that the sum of weights per death certificate usually don’t add to one. Duplicates and ill-defined causes in OC were excluded. The data may present an external and non-external UC. When an external UC was present, it was used as the UC.

We calculated both age standardised UC (ASRuc), i.e. unweighted death counts, and age standardised weighted counts (ASRw) for COVID-19 in 2020 to 2024, for age groups 0-19, 20-39, 40-59, 60-74, 75-84 and 85+ years. We applied the Dobson variance estimator to calculate 95% confidence intervals for directly age-standardised rates [24]. Lower ASRw than ASRuc (i.e., a ratio of ASRw/ASRuc below 1.00) suggests that COVID-19 was more commonly chosen to be placed as UC rather than OC, compared to other causes. Very low values may potentially indicate overcounting of COVID-19 deaths. Additionally, we compared the 10 major causes of death across all years from 2019 to 2024 using ASRw and ASRuc. The main comparison was for 2024 (post-pandemic) versus 2019 (pre-pandemic, death certificates causes other than UC were not available before 2019). Similarly, we ran the same comparison for the 19 high-level groupings of death causes (such as all infectious diseases, all neoplasms, all external etc – see supplement for full list derived from Bishop et al. [23]). For 2020 to 2024, we also reported the ten most frequent causes on death certificates co-occurring with COVID-19, and the percentage of those in which COVID-19 was the UC.

We performed sensitivity analyses by applying two different weighting schemes. (1) all causes are weighted the same (including UC) and (2) the UC was weighted double the weight of a single OC in the respective death certificate [23].

Additionally, we reran the analyses using more stringent exclusion criteria to address noisy data [25]. See Supplements for full list of exclusions.

## RESULTS

### Pre- and post-pandemic mortality

Mortality rate ratios show that in all strata aged 40 years or older, the post pandemic mortality rate was either equal or lower than before the pandemic (Figure 1 and Table S2). The mortality ratios were numerically better than 0.929 (based on the extrapolation of the age-standardised mortality rate ratio between 2019 and the average of 2012-2014) in 5 of the 12 age-sex strata 40 years and older.

**Fig. 1:**
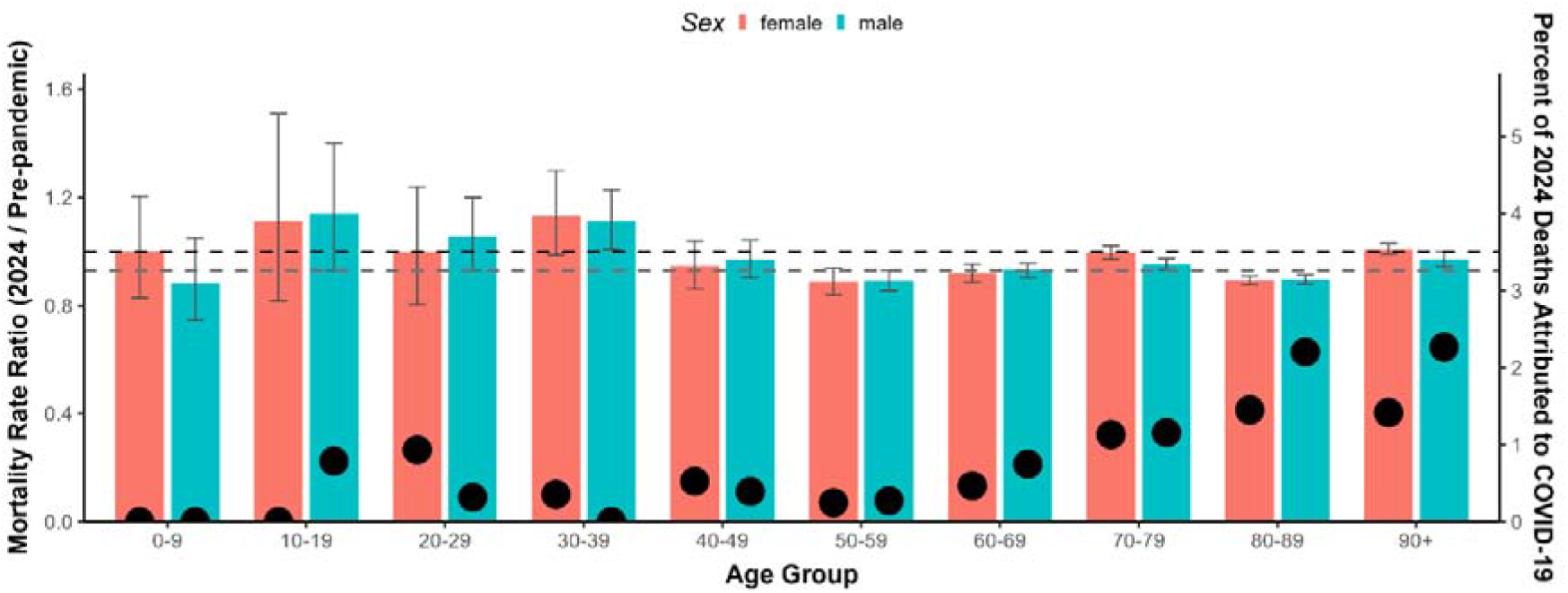
Mortality rate ratios between 2024 and pre-pandemic by age-group and sex. Pre-pandemic is based on years 2017-2019. Rates under 1.00 (black dashed line) indicate fewer relative deaths in 2024 than in the pre-pandemic period. The grey dashed line shows the value 0.929, which represents the optimistic “expected” value under an assumption of continued improvements absent the pandemic without the deceleration in improvement rates that had been seen in pre-pandemic years. Right axis shows percentage of all-cause deaths attributed to COVID-19 in 2024 (black dots). Note that COVID-19 death percentages in age groups under 40 years are based on one (or zero) COVID-19 death within the respective groups (Table S3). See exact values in Table S2.

Conversely, some mortality rate ratios were numerically elevated in younger groups (i.e. 10-19, 20-29 (male only) and 30-39 years old). Significantly so in the male 30-39 year old group (Figure S1, Table S4). In these groups only one individual was categorized to have died due to COVID-19 in 2024 (Table S3). To explore potential reasons for the increased youth mortality, we additionally investigated the mortality rate ratios of these age groups in the high-level causes previously described (see list of causes in supplements), based on the unfiltered death certificate data. Higher mortality rates in 2024 as compared to pre-pandemic in the 10-39 year olds were due to moderate increases in multiple high level causes (Figure S1, Table S4). There were consistent increases in particular in mental & behavioural diseases, external causes and nervous system causes, though only external causes in the 10-19 year olds were significant.

#### Weighted death counts

There was a total of 538,525 death certificates in Austria for the years 2019 to 2024 (2019: 83,386; 2024: 88,486), the years with full information on specific death causes. This included 2,502,281 non-duplicate ICDs code mentions (2019: 369,844; 2024: 424,707). We excluded 259,041 (2019: 37,927; 2024: 44,968) ill-defined causes in OC (see lists below for definitions of intermediate, immediate and ill-defined causes). We also removed all duplicate causes that were created when mapping ICD codes to the causes list. The final dataset for this sensitivity analysis had 1,888,980 mentions of causes (2019: 281,185; 2024: 316,291). The allocation to high-level causes led to further exclusion of OCs due to multiple causes being allocated to the same high-level cause. After excluding duplicates, the final high-level analysis had a total of 1,340,943 mentions of causes (2019: 199,781; 2024: 224,967).

The ASRw of COVID-19 was much lower than ASRuc (Figure 2 and Table S5). While mentions of COVID-19 in death certificates decreased drastically from 2020 to 2024, the ratio between ASRw and ASRuc in age groups older than 60 barely changed (0.52 in 2020 versus 0.54-0.58 in 2024). In those 40-59 years old, the ratio increased from 0.53 in 2020 to 0.75 in 2024.

**Fig. 2:**
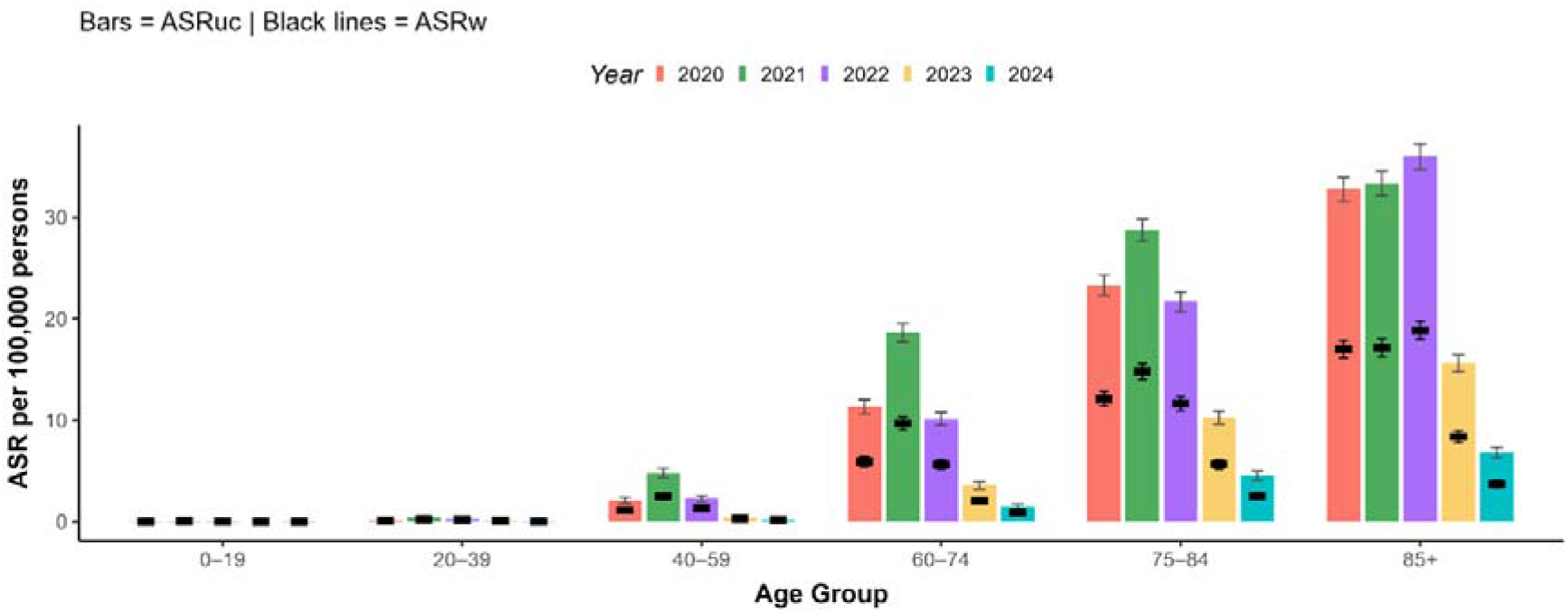
Stratified age standardised unweighted (ASRuc) and weighted COVID-19 death counts (ASRw) from 2020 to 2024. Bars indicate 95% confidence intervals. ASRuc = age standardised underlying causes; ASRw = age standardised weighted counts

When comparing the 10 most frequent causes of death between 2024 and 2019, age standardised estimates indicated a relative decrease in reported “Ischaemic heart disease” and a relative increase in “Alzheimer’s disease and dementia” as causes of death (Figure 3 and Table S6). While many of the top 10 causes had a ratio of ASRw/ASRuc lower than 1.00, COVID-19 had an extremely low ratio comparable only to “Colorectal cancer”, “Lung cancer” and “Residual - external causes” (Table S6).

**Fig. 3:**
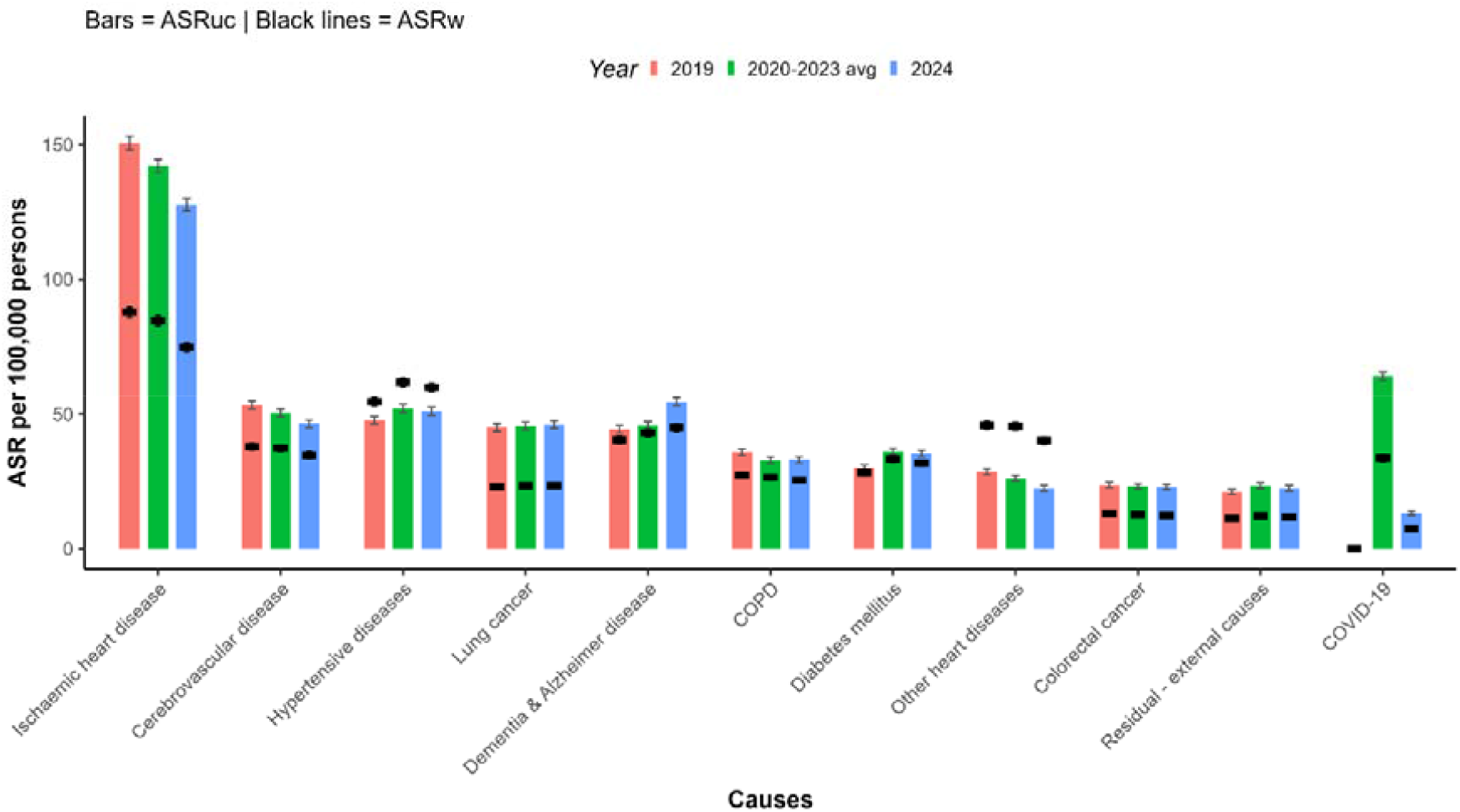
Age standardised underlying causes (ASRuc) and weighted death counts (ASRw) of the top 10 causes of death (as of 2019) and COVID-19 for 2019, 2020-2023 and 2024. The category “Ill-defined causes” is not shown. Bars indicate 95% confidence intervals. ASRuc = age standardised underlying causes; ASRw = age standardised weighted counts.

High-level causes indicated an increase in “Respiratory diseases” between 2020 and 2023 as compared to 2024 and 2019, when COVID-19 was included in this group (Figure 4 & Table S7). Conversely, when COVID-19 was not included, “Respiratory diseases” showed a marked decrease in 2020-2023 (ASRuc 52.44 versus 60.72 in 2019 and 58.26 in 2024); moreover, the ASRw/ASRuc ratio (95% CI) was highest in 2020-2023 (1.64 (1.59 – 1.71) versus 1.47 (1.41 – 1.52) in 2019 and 1.40 (1.35 – 1.45) in 2024).

**Fig. 4:**
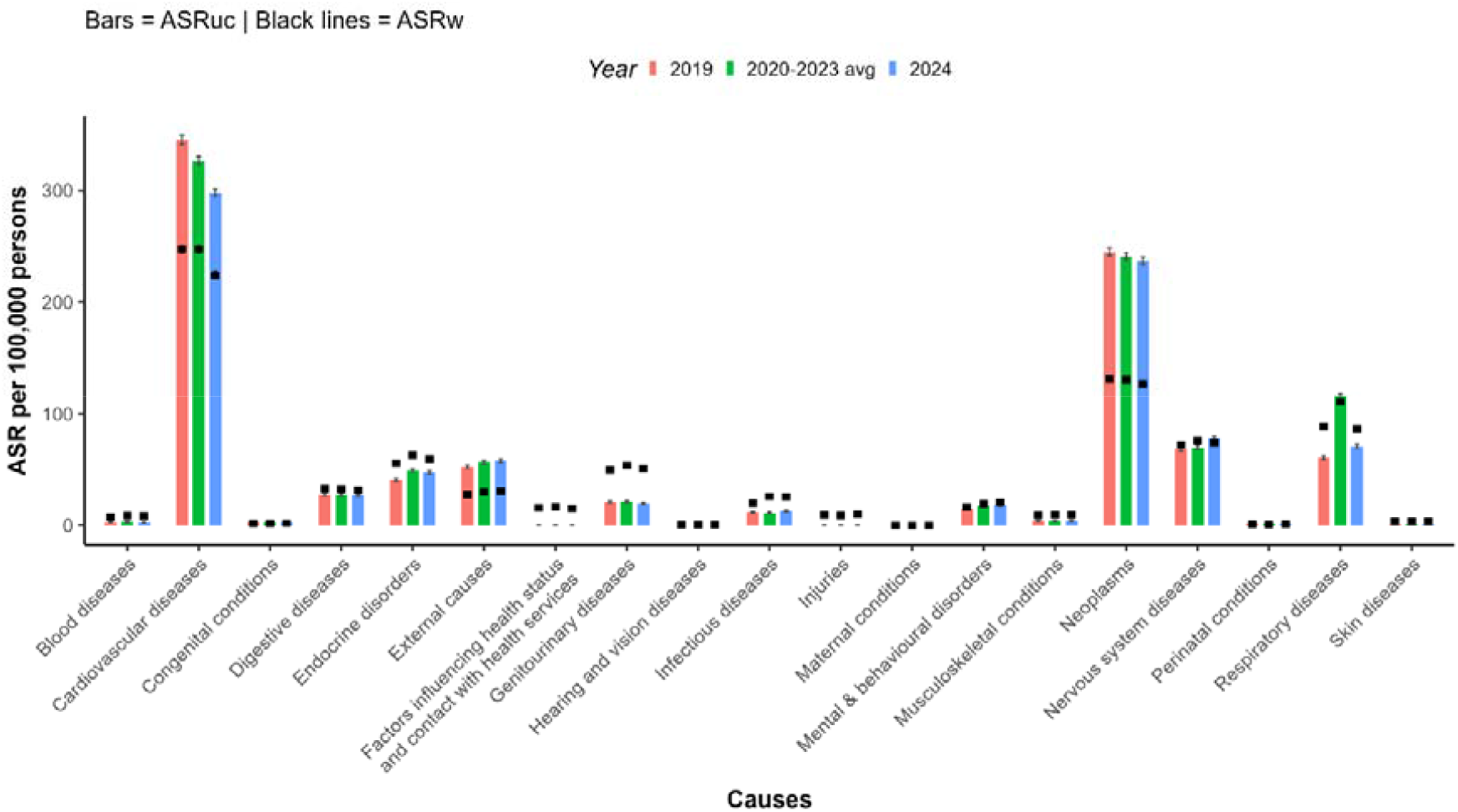
Age standardised underlying causes (ASRuc) and weighted death counts (ASRw) of the 19 high-level factors for 2019, 2020-2023 and 2024. COVID-19 is included in “Respiratory diseases”. The category “Ill-defined causes” is not shown. Bars indicate 95% confidence intervals. ASRuc = age standardised underlying causes; ASRw = age standardised weighted counts.

Causes most mentioned together with COVID-19 in 2024 were: “Pneumonia”, “Residual – infections”, “Hypertensive Disease”, “Ischaemic heart disease”, and “Renal failure” (Table 1). Previous pandemic years mostly showed the same causes, though in a slightly different order (Table S8). When co-mentioned with these causes, COVID-19 was listed as the UC in 88-98% of these deaths in 2020. While this gradually decreased over time, in 2024 COVID-19 was still listed as the UC in 68-85% of these deaths.

**Table 1:** Frequencies of causes most co-occurring with COVID-19 (in 2024) by year, and percentage where COVID-19 was determined the underlying cause.

| COVID as underlying cause | 2024 | 2023 | 2022 | 2021 | 2020 |
| --- | --- | --- | --- | --- | --- |
|  | frequency co-occurring (COVID-19 as UC percentage) |  |  |  |  |
| Residual - infections | 1,031<br>(70.2%) | 1,315<br>(77.9%) | 2,615<br>(81.6%) | 2,756<br>(90.4%) | 2,134<br>(97.7%) |
| Pneumonia | 808<br>(80.3%) | 1,587<br>(87%) | 2,957<br>(93%) | 4,848<br>(94.7%) | 3,069<br>(97.8%) |
| Hypertensive diseases | 535<br>(72.1%) | 1,049<br>(78%) | 2,257<br>(84.8%) | 2,578<br>(91.8%) | 2,017<br>(93.9%) |
| Renal failure | 415<br>(69.9%) | 791<br>(74.1%) | 1,671<br>(85.3%) | 1,580<br>(94.3%) | 1,326<br>(93.8%) |
| Ischaemic heart disease | 287<br>(79.1%) | 616<br>(85.7%) | 1,301<br>(87.4%) | 1,481<br>(94.3%) | 1,194<br>(96.9%) |
| Dementia & Alzheimer disease | 292<br>(84.2%) | 643<br>(83.2%) | 1,439<br>(92.2%) | 1,109<br>(92.2%) | 1,230<br>(93.2%) |
| Atrial fibrillation | 299<br>(69.9%) | 588<br>(79.4%) | 1,049<br>(85.8%) | 1,086<br>(89.2%) | 818<br>(91.3%) |
| Diabetes mellitus | 291<br>(73.9%) | 511<br>(79.5%) | 1,222<br>(87.2%) | 1,517<br>(92.1%) | 1,183<br>(94.5%) |
| Other heart diseases | 298<br>(68.8%) | 633<br>(68.7%) | 1,243<br>(82.4%) | 1,134<br>(87.8%) | 970<br>(88.8%) |
| Cerebrovascular disease | 193<br>(72.5%) | 401<br>(79.1%) | 848<br>(87.5%) | 643<br>(93.6%) | 624<br>(93.4%) |

Sensitivity analyses showed that alternative weighing methods did not change the findings on COVID-19 (Table S9). ASRw for the alternative weighting methods was even lower than for the weighting used in the main analysis (Table S9; e.g. 2020: main 36.49, equally weighted causes 21.00, UC double the weight of OC 30.56; 2024: main 7.12 equally weighted causes 4.15, UC double the weight of OC 5.68).

Using more stringent exclusion criteria did not change the findings concerning COVID-19 either (Table S10, Figure S2 & S3). However, it did affect ASRw/ASRuc ratios of some other prevalent causes such as “Hypertensive diseases” and changed the overall prevalence of multiple top-10 causes (Figure S3). It also led to changes in co-occurrences, in particular for “Pneumonia” (Table S8, S11, & S12).

## DISCUSSION

We showed that in Austria the relative mortality of age groups of 60 years or older (i.e. those at substantive risk to die of COVID-19) was lower (even substantially lower in many strata) in 2024 than it was pre-pandemic (2017-2019). Conversely, increased post-pandemic mortality was documented for young age groups. Analysis of multiple causes of death via death certificates, offered hints that COVID-19 deaths were possibly overestimated. The over-estimation may have apparently occurred not only in 2024, but throughout the entire pandemic. When COVID-19 was listed as a cause, it was almost always listed as the underlying cause during the pandemic. While gradually a larger share of COVID-19 was listed as “other cause”, even in 2024 it was mostly listed as the underlying cause.

SARS-CoV2 infections seemingly no longer had any substantial impact on mortality patterns in 2024. Age strata that had significant or more than 5% increased mortality in 2024 compared to the pre-pandemic (mortality ratio >1.05), were low-risk strata of young individuals with a single, or no reported COVID-19 death. It is unlikely that those differences are directly caused by recent undiscovered SARS-CoV-2 infections. Conversely, the increase in mortality in young individuals should raise concerns about the advent of non-COVID-19 reasons and/or reasons related to adverse consequences of the pandemic response (e.g., mental health, external causes and nervous system diseases). Escalating problems of violence, alcohol, drug overdose, and suicides in young age strata have been described already in different countries [26–29] and may create a persisting legacy of the pandemic that needs to be tracked and addressed.

We recently estimated 2.5 million SARS-CoV-2 infections in 2024 across Austria [30]. In this context the total reported COVID-19 death count of 1,212 is already low, corresponding to an infection fatality rate of 0.048%. However, given our findings on death certificates we are possibly still overestimating COVID-19 deaths and hence also the infection fatality rate. ASRw was lower than ASRuc throughout the pandemic, with no major changes in their ratios between 2020 and 2024 for those 60 years and older. This indicates that even with lower UC counts, COVID-19 was, and still is potentially overcounted as an UC. If no overcounting took place in 2024, the inconspicuous mortality rate ratios suggest that deaths mainly occurred in populations with very low life expectancy. This aligns with our previously published study estimating number of people needed to vaccinate to save one life (NNV) (and one life year) in 2024 in Austria [31]. There we estimated that, assuming correct COVID-19 death counts, the only strata with a COVID-19 mortality rate high enough to indicate a NNV below 1000 (85+ year old nursing home residents) had average life expectancies below 1 year.

Changes in the top 10 causes of death in 2019 and 2024 may reflect real changes or artefacts of death certificate compilation. The decrease in “Ischaemic heart disease” in particular, is likely too large to reflect a genuine improvement. This may partially be due to a change in UC categorization of individuals with multiple comorbidities and unclear main reason for death. Here the tendency of using “Ischaemic heart disease” as an UC may have become partially replaced by UC categorized as COVID-19.

In high-level causes the effect of COVID-19 may be seen against the group of “Respiratory diseases”. Throughout the pandemic (2020-2023), there was a decrease in other recorded respiratory diseases. Respiratory diseases have traditionally had a high ASRw/ASRuc, suggesting that they are typically listed as contributing rather than underlying causes of death. This pattern was markedly reversed with COVID-19.

Sensitivity analyses showed that our findings on COVID-19 are robust to different weighing approaches. Tested alternative approaches indicated even lower weighted estimates for COVID-19 deaths than the main analysis. COVID-19 deaths may have been both under- and over-counted during the pandemic in different locations and time periods [6]. However, overcounting is far more likely in countries like Austria. Evidence of major overcounting in high-income countries has also accumulated from audits of medical records. In Greece, only 64.9% of audited hospital deaths listed as COVID-19 were found to be attributable or related to COVID-19 upon auditing [7]. In Sweden, 24% of audited deaths that were listed as COVID-19 in death certificates were found to have absolutely no relationship to COVID-19 upon auditing. In most of the others, COVID-19 was found upon audit to be contributing rather than underlying cause. Overall, while death certificates had counted 799 deaths as having COVID-19 UC, clinical record audit found only 213 such deaths [8]. Among 55 deaths recorded as due to COVID-19 in the winter of 2021-2022 in Ireland [32], clinical opinion considered that COVID-19 was the primary cause of death in only 72.7% (40/55). The extent of over-counting of COVID-19 deaths apparently increased with Omicron variants, as documented also in a study in Denmark [33]. Audit studies are also needed in non-high-income countries; there, large under-counting of COVID-19 deaths due to low-testing is sometimes speculated. However, one medical record audit study in Colombia [34] found only 6% undercounting of COVID-19 deaths (231 in death certificates versus 247 in the audit) in 2021. Another study auditing 339 COVID-19-related death certificates in Iran in 2020 found major errors in 58% and minor errors in all of them [34]. These emerging data suggest that under-counting of COVID-19 deaths was a much lesser problem than initially believed, while over-counting has been highly prominent in several settings.

The presented estimates have limitations. First, while we stratify by age group and sex, we do not have data on comorbidities. Therefore, our findings cannot be simply generalized to specific patient populations (e.g., immunocompromised patients or nursing home residents) who might have a particularly high risk of COVID-19 deaths. Even within extremely high-risk populations, outcomes may differ. Second, the presented values are also mostly descriptive and the shortcomings of reporting in death certificates should be kept in mind [35]. Empirical studies have suggested very high rates of major errors in death certificates, exceeding 50% even in pre-pandemic years [36]. Some causes may be more likely to be chosen as UC than others due to biased perceptions and/or lack of familiarity of the person compiling the certificate. The impact of biases may have been even higher during the highly charged, pressurized conditions of the pandemic crisis and may have fuelled major over-reporting of COVID-19 as UC. Additional in-depth auditing of medical records would be helpful. While there have been multiple small updates to the Austrian ICD coding practices between 2017 and 2024 (2020, 2022 and 2024), changes were within disease categories (other than the implementation of COVID) and are thus unlikely to have affected analyses substantially. However, for COVID-19 in particular, it is possible that the coding algorithms may have favoured over-counting of COVID-19 deaths [37].

Finally, there can be debate on whether mortality rates across age strata should continue to decrease over time, and, if so, what might be a good enough rate. This would translate to mortality rate ratios of <1.00 being desirable for 2024 versus 2019. Our results and inferences hold true even if substantial reduction in mortality is assumed to be a reasonable counterfactual of what would have happened between 2017-2019 and 2024 absent the pandemic. Our optimistic sensitivity analysis assumed that mortality rates would continue to decrease linearly after 2019 (i.e. without the deceleration in improvements that had long been seen before the pandemic). In fact, under this assumption, the observed increase in mortality rates in young strata would be even more worrisome. Overall, it is unclear to what extent further decreases in mortality (and consequently increases in life expectancy) are feasible in countries that have already reached very high life expectancy. It is well documented that high-income countries (including Austria) slowed down (and occasionally even reversed) life expectancy gains for many years before the pandemic [5,38] and this may have been due to a composite of health and socioeconomic reasons, such as austerity [39]. One may speculate whether long-term austerity and socioeconomic disadvantage may have primed young age strata to be hit most hard by the pandemic and the pandemic response with perpetuating legacy of adverse impact long after the pandemic.

In summary, nationwide mortality data in Austria show no visible COVID-19 mortality burden in the post-pandemic phase and potentially indicate overcounting of COVID-19 deaths from 2020-2024. These findings should be factored in ongoing public health policies. Increased mortality patterns in young individuals are of concern, requiring confirmation and further elucidation of the underlying reasons and proper intervention approaches.

## Data Availability

Death certificate data were provided by Statistics Austria. Data are available through an application at Statistics Austria. All other data is publicly available.

https://www.statistik.at/statistiken/bevoelkerung-und-soziales/bevoelkerung/gestorbene/todesursachen

https://www.statistik.at/en/statistics/population-and-society/population/population-stock/population-at-beginning-of-year/quarter

https://www.statistik.at/en/statistics/population-and-society/population/demographische-prognosen/population-projections-for-austria-and-federal-states

## Statements & Declarations

### Author Contributions

JPAI conceptualized the study. UR wrote the original draft of the manuscript. SP acquired funding. UR wrote software. UR performed the formal analyses. SP administered the project. All authors contributed to writing, reviewing, and editing the manuscript, and approved the final version before submission.

## Acknowledgements

The authors thank all persons and organizations involved in data collection.

