## Supplements for "Post-pandemic mortality patterns and COVID-19 burden considering multiple death causes"

**Supplements: Post-pandemic mortality patterns and the potential of SARS-CoV-2 vaccinations in Austria**

### **Table S1:** STROBE Statement—Checklist of items for *cohort studies*

|  | **Item No** | **Recommendation** | **Main text page** |
| --- | --- | --- | --- |
| **Title and abstract** | 1 | (*a*) Indicate the study’s design with a commonly used term in the title or the abstract | Page 2, Abstract |
|  |  | (*b*) Provide in the abstract an informative and balanced summary of what was done and what was found | Page 2 , Abstract |
| **Introduction** | | |  |
| Background/rationale | 2 | Explain the scientific background and rationale for the investigation being reported | Page 3 and 4, Introduction |
| Objectives | 3 | State specific objectives, including any prespecified hypotheses | Page 3 and 4, Introduction |
| **Methods** | | |  |
| Study design | 4 | Present key elements of study design early in the paper | Page 4 and 5, Methods |
| Setting | 5 | Describe the setting, locations, and relevant dates, including periods of recruitment, exposure, follow-up, and data collection | Page 4 and 5, Methods |
| Participants | 6 | (*a*) Give the eligibility criteria, and the sources and methods of selection of participants. Describe methods of follow-up | Page 5 to 7, Methods |
|  |  | (*b*) For matched studies, give matching criteria and number of exposed and unexposed | Not applicable |
| Variables | 7 | Clearly define all outcomes, exposures, predictors, potential confounders, and effect modifiers. Give diagnostic criteria, if applicable | Page 5 to 7, Methods |
| Data sources/ measurement | 8* | For each variable of interest, give sources of data and details of methods of assessment (measurement). Describe comparability of assessment methods if there is more than one group | Page 5 to 7, Methods |
| Bias | 9 | Describe any efforts to address potential sources of bias | Page 6 and 7, Methods |
| Study size | 10 | Explain how the study size was arrived at | Only partially applicable,  Page 6 and 7, Methods |
| Quantitative variables | 11 | Explain how quantitative variables were handled in the analyses. If applicable, describe which groupings were chosen and why | Page 5 to 7, Methods |
| Statistical methods | 12 | (*a*) Describe all statistical methods, including those used to control for confounding | Not applicable. |
|  |  | (*b*) Describe any methods used to examine subgroups and interactions | Page 5 to 7, Methods |
|  |  | (*c*) Explain how missing data were addressed | Page 6 and 7, Methods |
|  |  | (*d*) If applicable, explain how loss to follow-up was addressed | Not applicable |
|  |  | (*e*) Describe any sensitivity analyses | Not applicable |
| **Results** | | |  |
| Participants | 13* | (a) Report numbers of individuals at each stage of study—eg numbers potentially eligible, examined for eligibility, confirmed eligible, included in the study, completing follow-up, and analysed | Page 7 to 12 and Supplement Tables |
|  |  | (b) Give reasons for non-participation at each stage | Page 8, Results |
|  |  | (c) Consider use of a flow diagram | Not applicable |
| Descriptive data | 14* | (a) Give characteristics of study participants (eg demographic, clinical, social) and information on exposures and potential confounders | Not applicable |
|  |  | (b) Indicate number of participants with missing data for each variable of interest | Not applicable |
|  |  | (c) Summarise follow-up time (eg, average and total amount) | Not applicable |
| Outcome data | 15* | Report numbers of outcome events or summary measures over time | Page 7 to 12, Results |
| Main results | 16 | (*a*) Give unadjusted estimates and, if applicable, confounder-adjusted estimates and their precision (eg, 95% confidence interval). Make clear which confounders were adjusted for and why they were included | Not applicable |
|  |  | (*b*) Report category boundaries when continuous variables were categorized | Not applicable |
|  |  | (*c*) If relevant, consider translating estimates of relative risk into absolute risk for a meaningful time period | Not applicable |
| Other analyses | 17 | Report other analyses done—eg analyses of subgroups and interactions, and sensitivity analyses | Page 9 to 12, Results |
| **Discussion** | | |  |
| Key results | 18 | Summarise key results with reference to study objectives | Page 12, Discussion |
| Limitations | 19 | Discuss limitations of the study, taking into account sources of potential bias or imprecision. Discuss both direction and magnitude of any potential bias | Page 15 and 16, Discussion |
| Interpretation | 20 | Give a cautious overall interpretation of results considering objectives, limitations, multiplicity of analyses, results from similar studies, and other relevant evidence | Page 12 to 15, Discussion |
| Generalisability | 21 | Discuss the generalisability (external validity) of the study results | This is a nationwide survey |
| **Other information** | | |  |
| Funding | 22 | Give the source of funding and the role of the funders for the present study and, if applicable, for the original study on which the present article is based | Page 16 |

*Give information separately for exposed and unexposed groups.

**Note:** An Explanation and Elaboration article discusses each checklist item and gives methodological background and published examples of transparent reporting. The STROBE checklist is best used in conjunction with this article (freely available on the Web sites of PLoS Medicine at http://www.plosmedicine.org/, Annals of Internal Medicine at http://www.annals.org/, and Epidemiology at http://www.epidem.com/). Information on the STROBE Initiative is available at <http://www.strobe-statement.org>.

### **Supplementary Methods**

#### **Age-standardised rate:**

Age standardises rates are calculated per age group:

$$ASR=\sum_{x} \left( \frac{d_{x}}{P_{x}}*w_{x} \right)$$

Where x is the age group, d is deaths (either UC mentions for ASRuc; or the 50%/50% UC and CC weighted outcomes for ASRw), P is the population, and w is the proportion of the age group in the total population in Austria across all years.^3^

#### **Stringent exclusion criteria sensitivity analysis**

All exclusions were performed on both UC and OC. If an excluded ICD was present as an UC, the whole certificate was removed from the analysis. First, we excluded all causes that were previously categorized as “immediate or intermediate causes” (see page 35 in supplements). This may address the issue that we only have access to OC, which is a combination of CC, immediate and intermediate causes. As the method was originally designed to use CC, the use of OC might lead to a relative underweighting of CCs. As previously mentioned, there may be both an external and non-external UC documented. In those cases, we excluded the associated non-external UC (always injury) from the respective death certificate, to avoid double counting the same ailments twice. We also assume that co-occurrence of external causes and injuries in OC represent the same cause (i.e., the injury following from the external cause). Thus, in case of co-occurrence, we excluded the stated injury. Lastly, we excluded ill-defined causes.

### **Supplementary Results**

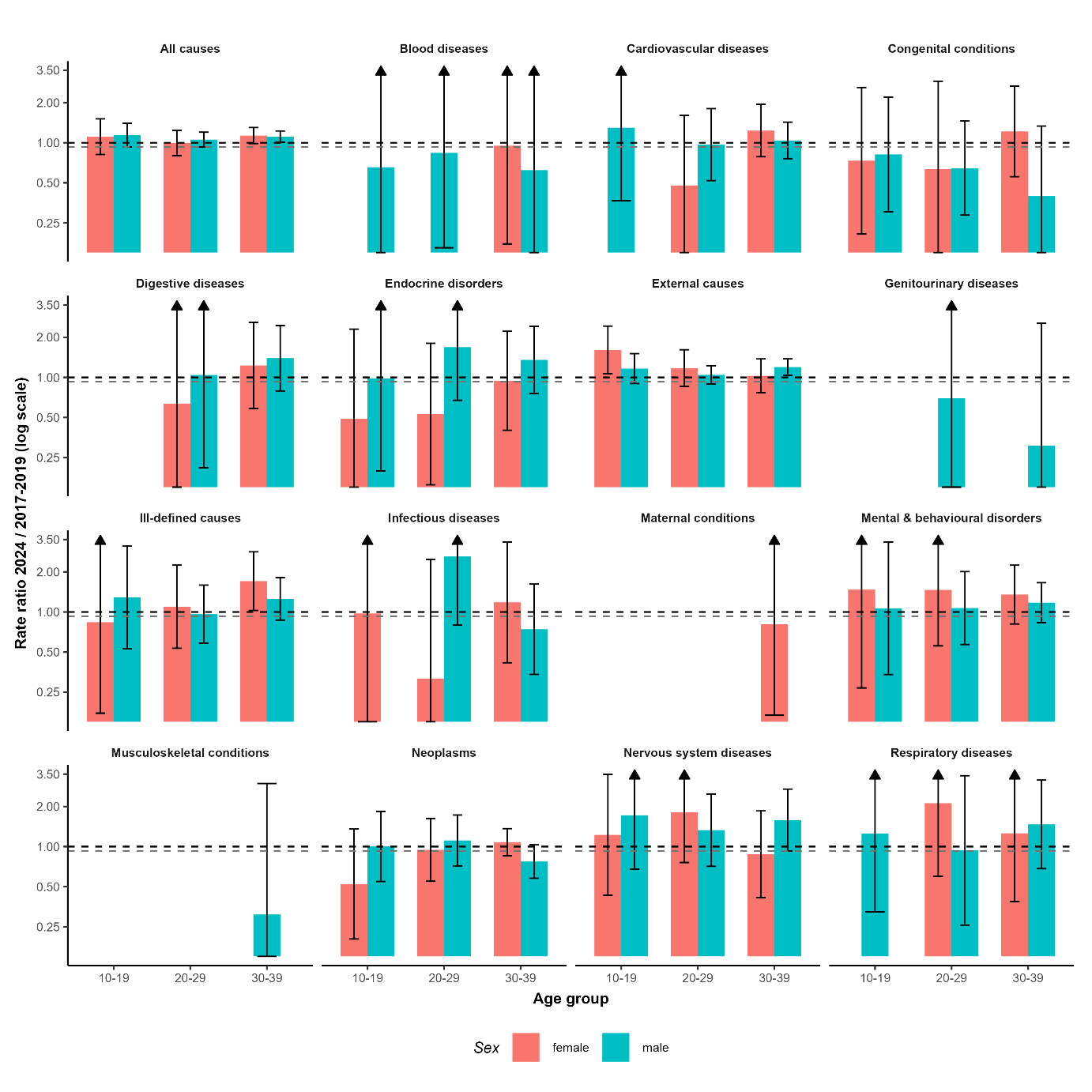

**Figure S1:** Mortality rate ratios and 95% confidence intervals of high-level causes for age groups 10-19, 20-29 and 30-39 by sex. Arrows indicate upper CI higher than 3.5 which are not shown for illustrative purposes. The grey dashed line shows the value 0.929, which represents the optimistic “expected” value under an assumption of continued improvements absent the pandemic without the deceleration in improvement rates that had been seen in pre-pandemic years. See Table S4 for specific values.

| Table S2: Pre- and post-pandemic mortality ratios and percentages of deaths categorized as COVID-19 in 2024 stratified by age group and sex. | | | | |
| --- | --- | --- | --- | --- |
| age group | mortality ratios (95% CI) | | Covid death (% in 2024) | |
|  | male | female | male | female |
| 0-9 | 0.884 (0.745 - 1.049) | 0.999 (0.829 - 1.204) | 0 | 0 |
| 10-19 | 1.141 (0.929 - 1.4) | 1.11 (0.816 - 1.51) | 0.787 | 0 |
| 20-29 | 1.056 (0.93 - 1.2) | 0.998 (0.803 - 1.24) | 0.317 | 0.935 |
| 30-39 | 1.112 (1.008 - 1.227) | 1.133 (0.986 - 1.3) | 0 | 0.355 |
| 40-49 | 0.971 (0.905 - 1.041) | 0.946 (0.862 - 1.04) | 0.391 | 0.525 |
| 50-59 | 0.892 (0.856 - 0.93) | 0.888 (0.84 - 0.939) | 0.274 | 0.25 |
| 60-69 | 0.93 (0.905 - 0.956) | 0.92 (0.887 - 0.955) | 0.749 | 0.467 |
| 70-79 | 0.953 (0.932 - 0.974) | 0.996 (0.971 - 1.022) | 1.156 | 1.132 |
| 80-89 | 0.897 (0.88 - 0.914) | 0.893 (0.877 - 0.909) | 2.203 | 1.452 |
| 90+ | 0.972 (0.944 - 1.001) | 1.01 (0.99 - 1.03) | 2.267 | 1.417 |

| Table S3: Deaths, populations and Covid deaths for different years stratified by 10-year age groups and sex | | | | | | |
| --- | --- | --- | --- | --- | --- | --- |
| age group | deaths all cause | | | | | |
|  | 2017 | | 2018 | | 2019 | |
|  | male | female | male | female | male | female |
| 0-9 | 188 | 152 | 174 | 131 | 200 | 146 |
| 10-19 | 109 | 56 | 106 | 47 | 112 | 45 |
| 20-29 | 312 | 107 | 321 | 109 | 301 | 125 |
| 30-39 | 444 | 220 | 446 | 261 | 493 | 224 |
| 40-49 | 1094 | 612 | 1096 | 649 | 1055 | 605 |
| 50-59 | 3335 | 1766 | 3393 | 1850 | 3153 | 1738 |
| 60-69 | 6062 | 3548 | 6205 | 3472 | 6060 | 3536 |
| 70-79 | 10849 | 7690 | 11272 | 7880 | 11055 | 7892 |
| 80-89 | 12602 | 15553 | 12509 | 15064 | 12752 | 14763 |
| 90+ | 4907 | 13664 | 5329 | 13661 | 5593 | 13538 |
| age group | population | | | | | |
|  | 2017 | | 2018 | | 2019 | |
|  | male | female | male | female | male | female |
| 0-9 | 432648 | 407642 | 438432 | 412479 | 440432 | 414987 |
| 10-19 | 454327 | 423108 | 448661 | 419927 | 444009 | 418268 |
| 20-29 | 590860 | 560651 | 588722 | 558439 | 582129 | 552342 |
| 30-39 | 592093 | 578958 | 600406 | 586672 | 609169 | 593955 |
| 40-49 | 632288 | 634557 | 617051 | 620928 | 602334 | 606882 |
| 50-59 | 667321 | 667915 | 680322 | 679680 | 692191 | 690277 |
| 60-69 | 457792 | 500102 | 465495 | 506252 | 474962 | 514324 |
| 70-79 | 331907 | 407196 | 342816 | 420619 | 350075 | 429922 |
| 80-89 | 133979 | 220282 | 136129 | 218845 | 140081 | 220825 |
| 90+ | 19059 | 59032 | 20337 | 59036 | 21520 | 59085 |
| age group | deaths all cause | | deaths covid | | population | |
|  | 2024 | | 2024 | | 2024 | |
|  | male | female | male | female | male | female |
| 0-9 | 171 | 148 | 0 | 0 | 451310 | 426489 |
| 10-19 | 127 | 56 | 1 | 0 | 458610 | 429797 |
| 20-29 | 315 | 107 | 1 | 1 | 562462 | 525664 |
| 30-39 | 552 | 282 | 0 | 1 | 646623 | 621474 |
| 40-49 | 1022 | 571 | 4 | 3 | 600871 | 602103 |
| 50-59 | 2920 | 1603 | 8 | 4 | 675740 | 687028 |
| 60-69 | 6943 | 3854 | 52 | 18 | 569452 | 603516 |
| 70-79 | 10464 | 7687 | 121 | 87 | 339252 | 413789 |
| 80-89 | 15391 | 16945 | 339 | 246 | 185971 | 275921 |
| 90+ | 6131 | 13197 | 139 | 187 | 24275 | 56647 |

| Table S4: Mortality rate ratios between 2024 and 2017-2019 per cause, age group and sex, for age groups between 10 and 39. Death rates and number of deaths for 2024 and 2017-2019 included. | | | | | | | |
| --- | --- | --- | --- | --- | --- | --- | --- |
| age group | sex | cause | death in 2024 (n) | rate in 2024 | avg deaths 2017-19 (avg n) | rate in 2017-19 | ratio of rates (2024/2017-19) (95% CI) |
| 10-19 | female | All causes | 56 | 13.03 | 49.3333333 | 11.73 | 1.111 (0.817 - 1.511) |
| 10-19 | female | Blood diseases | 1 | 0.23 |  |  |  |
| 10-19 | female | Congenital conditions | 3 | 0.70 | 4 | 0.95 | 0.734 (0.207 - 2.601) |
| 10-19 | female | Endocrine disorders | 2 | 0.47 | 4 | 0.95 | 0.489 (0.104 - 2.301) |
| 10-19 | female | External causes | 35 | 8.14 | 21.3333333 | 5.07 | 1.606 (1.064 - 2.425) |
| 10-19 | female | Ill-defined causes | 2 | 0.47 | 2.33333333 | 0.56 | 0.838 (0.174 - 4.033) |
| 10-19 | female | Infectious diseases | 1 | 0.23 | 1 | 0.24 | 0.975 (0.088 - 10.754) |
| 10-19 | female | Mental & behavioural disorders | 2 | 0.47 | 1.33333333 | 0.32 | 1.470 (0.269 - 8.024) |
| 10-19 | female | Neoplasms | 5 | 1.16 | 9.33333333 | 2.22 | 0.524 (0.202 - 1.358) |
| 10-19 | female | Nervous system diseases | 5 | 1.16 | 4 | 0.95 | 1.224 (0.431 - 3.475) |
| 10-19 | male | All causes | 127 | 27.69 | 109 | 24.28 | 1.141 (0.929 - 1.400) |
| 10-19 | male | Blood diseases | 1 | 0.22 | 1.5 | 0.33 | 0.655 (0.068 - 6.296) |
| 10-19 | male | Cardiovascular diseases | 4 | 0.87 | 3 | 0.67 | 1.298 (0.366 - 4.598) |
| 10-19 | male | Congenital conditions | 5 | 1.09 | 6 | 1.33 | 0.817 (0.303 - 2.200) |
| 10-19 | male | Endocrine disorders | 2 | 0.44 | 2 | 0.44 | 0.983 (0.198 - 4.869) |
| 10-19 | male | External causes | 80 | 17.44 | 67.3333333 | 15.00 | 1.163 (0.898 - 1.506) |
| 10-19 | male | Ill-defined causes | 7 | 1.53 | 5.33333333 | 1.19 | 1.283 (0.528 - 3.119) |
| 10-19 | male | Mental & behavioural disorders | 4 | 0.87 | 3.66666667 | 0.82 | 1.066 (0.339 - 3.347) |
| 10-19 | male | Neoplasms | 14 | 3.05 | 13.6666667 | 3.04 | 1.004 (0.547 - 1.841) |
| 10-19 | male | Nervous system diseases | 7 | 1.53 | 4 | 0.89 | 1.719 (0.677 - 4.365) |
| 10-19 | male | Respiratory diseases | 3 | 0.65 | 2.33333333 | 0.52 | 1.255 (0.324 - 4.852) |
| 20-29 | female | All causes | 107 | 20.36 | 113.666667 | 20.41 | 0.997 (0.803 - 1.239) |
| 20-29 | female | Cardiovascular diseases | 3 | 0.57 | 6.66666667 | 1.19 | 0.478 (0.142 - 1.609) |
| 20-29 | female | Congenital conditions | 2 | 0.38 | 3.33333333 | 0.60 | 0.636 (0.139 - 2.901) |
| 20-29 | female | Digestive diseases | 1 | 0.19 | 1.66666667 | 0.30 | 0.636 (0.074 - 5.441) |
| 20-29 | female | Endocrine disorders | 3 | 0.57 | 6 | 1.08 | 0.530 (0.156 - 1.801) |
| 20-29 | female | External causes | 52 | 9.89 | 47 | 8.44 | 1.172 (0.853 - 1.611) |
| 20-29 | female | Ill-defined causes | 10 | 1.90 | 9.66666667 | 1.73 | 1.097 (0.535 - 2.251) |
| 20-29 | female | Infectious diseases | 1 | 0.19 | 3.33333333 | 0.60 | 0.317 (0.041 - 2.475) |
| 20-29 | female | Mental & behavioural disorders | 6 | 1.14 | 4.33333333 | 0.78 | 1.466 (0.557 - 3.856) |
| 20-29 | female | Neoplasms | 17 | 3.23 | 19 | 3.42 | 0.946 (0.550 - 1.626) |
| 20-29 | female | Nervous system diseases | 8 | 1.52 | 4.66666667 | 0.84 | 1.812 (0.760 - 4.320) |
| 20-29 | female | Respiratory diseases | 4 | 0.76 | 2 | 0.36 | 2.118 (0.598 - 7.507) |
| 20-29 | male | All causes | 315 | 56.00 | 311.333333 | 53.01 | 1.056 (0.930 - 1.200) |
| 20-29 | male | Blood diseases | 2 | 0.36 | 2.5 | 0.42 | 0.839 (0.163 - 4.325) |
| 20-29 | male | Cardiovascular diseases | 13 | 2.31 | 14 | 2.39 | 0.969 (0.520 - 1.804) |
| 20-29 | male | Congenital conditions | 7 | 1.24 | 11.3333333 | 1.93 | 0.646 (0.286 - 1.456) |
| 20-29 | male | Digestive diseases | 2 | 0.36 | 2 | 0.34 | 1.042 (0.210 - 5.163) |
| 20-29 | male | Endocrine disorders | 7 | 1.24 | 4.33333333 | 0.74 | 1.686 (0.673 - 4.227) |
| 20-29 | male | External causes | 201 | 35.74 | 201 | 34.22 | 1.044 (0.890 - 1.225) |
| 20-29 | male | Genitourinary diseases | 1 | 0.18 | 1.5 | 0.26 | 0.697 (0.072 - 6.699) |
| 20-29 | male | Ill-defined causes | 20 | 3.56 | 21.6666667 | 3.69 | 0.964 (0.584 - 1.591) |
| 20-29 | male | Infectious diseases | 5 | 0.89 | 2 | 0.34 | 2.610 (0.797 - 8.552) |
| 20-29 | male | Mental & behavioural disorders | 13 | 2.31 | 12.6666667 | 2.16 | 1.070 (0.570 - 2.009) |
| 20-29 | male | Neoplasms | 27 | 4.80 | 25.3333333 | 4.31 | 1.113 (0.717 - 1.726) |
| 20-29 | male | Nervous system diseases | 14 | 2.49 | 11 | 1.87 | 1.330 (0.712 - 2.484) |
| 20-29 | male | Respiratory diseases | 3 | 0.53 | 3.33333333 | 0.57 | 0.937 (0.258 - 3.406) |
| 30-39 | female | All causes | 282 | 45.38 | 235 | 40.07 | 1.133 (0.986 - 1.300) |
| 30-39 | female | Blood diseases | 2 | 0.32 | 2 | 0.34 | 0.950 (0.174 - 5.186) |
| 30-39 | female | Cardiovascular diseases | 27 | 4.34 | 20.6666667 | 3.52 | 1.234 (0.785 - 1.938) |
| 30-39 | female | Congenital conditions | 9 | 1.45 | 7 | 1.19 | 1.216 (0.557 - 2.654) |
| 30-39 | female | Digestive diseases | 10 | 1.61 | 7.66666667 | 1.31 | 1.229 (0.585 - 2.582) |
| 30-39 | female | Endocrine disorders | 7 | 1.13 | 7 | 1.19 | 0.943 (0.401 - 2.219) |
| 30-39 | female | External causes | 62 | 9.98 | 57 | 9.71 | 1.027 (0.768 - 1.374) |
| 30-39 | female | Genitourinary diseases | 2 | 0.32 |  |  |  |
| 30-39 | female | Ill-defined causes | 24 | 3.86 | 13.3333333 | 2.26 | 1.705 (1.028 - 2.828) |
| 30-39 | female | Infectious diseases | 5 | 0.80 | 4 | 0.68 | 1.181 (0.416 - 3.352) |
| 30-39 | female | Maternal conditions | 2 | 0.32 | 2.33333333 | 0.40 | 0.809 (0.168 - 3.894) |
| 30-39 | female | Mental & behavioural disorders | 22 | 3.54 | 15.3333333 | 2.62 | 1.351 (0.813 - 2.246) |
| 30-39 | female | Neoplasms | 97 | 15.61 | 84.6666667 | 14.44 | 1.081 (0.855 - 1.365) |
| 30-39 | female | Nervous system diseases | 9 | 1.45 | 9.66666667 | 1.65 | 0.876 (0.415 - 1.851) |
| 30-39 | female | Respiratory diseases | 4 | 0.64 | 3 | 0.51 | 1.258 (0.387 - 4.086) |
| 30-39 | male | All causes | 552 | 85.37 | 461 | 76.73 | 1.113 (1.008 - 1.228) |
| 30-39 | male | Blood diseases | 1 | 0.15 | 1.5 | 0.25 | 0.622 (0.065 - 5.980) |
| 30-39 | male | Cardiovascular diseases | 53 | 8.20 | 47.3333333 | 7.87 | 1.041 (0.760 - 1.428) |
| 30-39 | male | Congenital conditions | 3 | 0.46 | 7 | 1.17 | 0.398 (0.119 - 1.334) |
| 30-39 | male | Digestive diseases | 18 | 2.78 | 12 | 2.00 | 1.393 (0.791 - 2.454) |
| 30-39 | male | Endocrine disorders | 17 | 2.63 | 11.6666667 | 1.94 | 1.353 (0.758 - 2.414) |
| 30-39 | male | External causes | 273 | 42.22 | 212.333333 | 35.35 | 1.194 (1.036 - 1.376) |
| 30-39 | male | Genitourinary diseases | 1 | 0.15 | 3 | 0.50 | 0.307 (0.037 - 2.547) |
| 30-39 | male | Ill-defined causes | 41 | 6.34 | 30.3333333 | 5.05 | 1.257 (0.869 - 1.817) |
| 30-39 | male | Infectious diseases | 8 | 1.24 | 10 | 1.67 | 0.742 (0.340 - 1.619) |
| 30-39 | male | Mental & behavioural disorders | 46 | 7.11 | 36.3333333 | 6.05 | 1.176 (0.833 - 1.659) |
| 30-39 | male | Musculoskeletal conditions | 1 | 0.15 | 3 | 0.50 | 0.310 (0.032 - 2.976) |
| 30-39 | male | Neoplasms | 59 | 9.12 | 71 | 11.81 | 0.772 (0.579 - 1.030) |
| 30-39 | male | Nervous system diseases | 21 | 3.25 | 12.3333333 | 2.05 | 1.583 (0.927 - 2.704) |
| 30-39 | male | Respiratory diseases | 10 | 1.55 | 6.33333333 | 1.05 | 1.468 (0.683 - 3.158) |

| Table S5: COVID-19 ASRuc, ASRw and their ratios per year and age group. | | | | | | | | | | | | | | | |
| --- | --- | --- | --- | --- | --- | --- | --- | --- | --- | --- | --- | --- | --- | --- | --- |
| age group | 2020 (95% CI) | | | 2021 (95% CI) | | | 2022 (95% CI) | | | 2023 (95% CI) | | | 2024 (95% CI) | | |
|  | ASRuc | ASRw | Ratio | ASRuc | ASRw | Ratio | ASRuc | ASRw | Ratio | ASRuc | ASRw | Ratio | ASRuc | ASRw | Ratio |
| 0–19 | 0.01 (-0.01 - 0.03) | 0.01 (-0.01 - 0.03) | 1 (0.06 - 15.99) | 0.09 (0.03 - 0.15) | 0.05 (0 - 0.09) | 0.5 (0.15 - 1.66) | 0.04 (0 - 0.09) | 0.03 (-0.01 - 0.06) | 0.62 (0.13 - 3.03) | 0 (0 - 0) | 0.01 (-0.01 - 0.02) |  | 0.01 (-0.01 - 0.03) | 0.01 (-0.01 - 0.02) | 0.5 (0.02 - 14.9) |
| 20–39 | 0.13 (0.06 - 0.21) | 0.08 (0.02 - 0.14) | 0.62 (0.25 - 1.54) | 0.43 (0.29 - 0.56) | 0.24 (0.14 - 0.34) | 0.56 (0.33 - 0.96) | 0.28 (0.17 - 0.39) | 0.18 (0.09 - 0.26) | 0.63 (0.33 - 1.18) | 0.11 (0.04 - 0.18) | 0.06 (0.01 - 0.11) | 0.56 (0.2 - 1.57) | 0.03 (0 - 0.07) | 0.02 (-0.01 - 0.06) | 0.74 (0.13 - 4.2) |
| 40–59 | 2.07 (1.77 - 2.37) | 1.1 (0.88 - 1.32) | 0.53 (0.42 - 0.68) | 4.78 (4.33 - 5.23) | 2.5 (2.17 - 2.82) | 0.52 (0.44 - 0.61) | 2.26 (1.95 - 2.57) | 1.32 (1.08 - 1.56) | 0.58 (0.47 - 0.73) | 0.41 (0.28 - 0.55) | 0.27 (0.16 - 0.38) | 0.66 (0.4 - 1.1) | 0.21 (0.12 - 0.31) | 0.16 (0.08 - 0.24) | 0.75 (0.38 - 1.49) |
| 60–74 | 11.32 (10.61 - 12.02) | 5.93 (5.42 - 6.44) | 0.52 (0.47 - 0.58) | 18.64 (17.75 - 19.53) | 9.66 (9.02 - 10.3) | 0.52 (0.48 - 0.56) | 10.14 (9.49 - 10.79) | 5.63 (5.15 - 6.11) | 0.56 (0.5 - 0.62) | 3.56 (3.18 - 3.94) | 2.06 (1.77 - 2.34) | 0.58 (0.48 - 0.69) | 1.47 (1.23 - 1.71) | 0.86 (0.67 - 1.04) | 0.58 (0.45 - 0.76) |
| 75–84 | 23.32 (22.33 - 24.32) | 12.1 (11.39 - 12.82) | 0.52 (0.48 - 0.56) | 28.73 (27.62 - 29.83) | 14.8 (14.01 - 15.59) | 0.52 (0.48 - 0.55) | 21.69 (20.73 - 22.65) | 11.62 (10.92 - 12.32) | 0.54 (0.5 - 0.58) | 10.25 (9.6 - 10.9) | 5.64 (5.16 - 6.13) | 0.55 (0.5 - 0.61) | 4.54 (4.11 - 4.96) | 2.54 (2.22 - 2.86) | 0.56 (0.48 - 0.65) |
| 85+ | 32.79 (31.6 - 33.98) | 17.03 (16.17 - 17.88) | 0.52 (0.49 - 0.55) | 33.33 (32.13 - 34.53) | 17.16 (16.29 - 18.02) | 0.51 (0.48 - 0.55) | 36 (34.75 - 37.25) | 18.86 (17.95 - 19.76) | 0.52 (0.49 - 0.56) | 15.61 (14.79 - 16.44) | 8.38 (7.77 - 8.98) | 0.54 (0.49 - 0.59) | 6.81 (6.27 - 7.35) | 3.69 (3.3 - 4.09) | 0.54 (0.47 - 0.62) |

| Table S6: ASRw, ASRuc, UC mentions and ratio of ASRw and ASRuc for 2019 and 2014 stratified by top 10 causes and COVID-19 | | | | | | | | |
| --- | --- | --- | --- | --- | --- | --- | --- | --- |
| Cause | 2019 | | | | 2024 | | | |
|  | ASRw per 100k persons | ASRuc per 100k persons | UC mentions | ASRw/ASRuc ratio | ASRw per 100k persons | ASRuc per 100k persons | UC mentions | ASRw/ASRuc ratio |
| COPD | 27.82 (26.7 - 28.94) | 36.55 (35.27 - 37.83) | 3,179 | 0.76 (0.72 - 0.8) | 24.82 (23.81 - 25.84) | 32.04 (30.89 - 33.19) | 3,026 | 0.77 (0.73 - 0.82) |
| COVID-19 | 0 (0 - 0) | 0 (0 - 0) | - |  | 7.12 (6.58 - 7.66) | 12.79 (12.07 - 13.52) | 1,212 | 0.56 (0.51 - 0.61) |
| Cerebrovascular disease | 38.5 (37.19 - 39.81) | 54.34 (52.78 - 55.9) | 4,733 | 0.71 (0.68 - 0.74) | 33.72 (32.54 - 34.9) | 44.97 (43.61 - 46.33) | 4,251 | 0.75 (0.72 - 0.79) |
| Colorectal cancer | 13.22 (12.46 - 13.99) | 24 (22.96 - 25.03) | 2,100 | 0.55 (0.51 - 0.59) | 12.03 (11.32 - 12.73) | 22.25 (21.29 - 23.21) | 2,103 | 0.54 (0.5 - 0.58) |
| Dementia & Alzheimer disease | 40.97 (39.62 - 42.32) | 45.03 (43.61 - 46.44) | 3,934 | 0.91 (0.87 - 0.95) | 43.65 (42.31 - 45) | 53.1 (51.62 - 54.58) | 5,006 | 0.82 (0.79 - 0.86) |
| Diabetes mellitus | 29.03 (27.89 - 30.17) | 30.72 (29.55 - 31.9) | 2,656 | 0.94 (0.89 - 1) | 30.85 (29.72 - 31.98) | 34.25 (33.06 - 35.44) | 3,238 | 0.9 (0.86 - 0.95) |
| Hypertensive diseases | 55.38 (53.81 - 56.96) | 48.39 (46.92 - 49.86) | 4,229 | 1.14 (1.1 - 1.19) | 58.16 (56.61 - 59.71) | 49.79 (48.35 - 51.22) | 4,682 | 1.17 (1.12 - 1.21) |
| Ischaemic heart disease | 89.23 (87.23 - 91.23) | 152.84 (150.23 - 155.45) | 13,336 | 0.58 (0.57 - 0.6) | 72.62 (70.89 - 74.35) | 123.82 (121.56 - 126.08) | 11,691 | 0.59 (0.57 - 0.6) |
| Lung cancer | 23.48 (22.46 - 24.51) | 45.95 (44.52 - 47.38) | 3,991 | 0.51 (0.48 - 0.54) | 22.73 (21.76 - 23.69) | 44.58 (43.23 - 45.93) | 4,224 | 0.51 (0.48 - 0.54) |
| Other heart diseases | 46.57 (45.13 - 48.01) | 29.17 (28.03 - 30.31) | 2,535 | 1.6 (1.52 - 1.68) | 39 (37.73 - 40.26) | 21.93 (20.98 - 22.88) | 2,067 | 1.78 (1.68 - 1.88) |
| Residual - external causes | 11.34 (10.63 - 12.04) | 21.3 (20.33 - 22.27) | 1,874 | 0.53 (0.49 - 0.57) | 11.66 (10.96 - 12.35) | 22.13 (21.18 - 23.09) | 2,067 | 0.53 (0.49 - 0.57) |

| Table S7: High-level causes and their ASRw, ASRuc, UC mentions and ASRw/ASRuc ratio for 2019 and 2024 | | | | | |
| --- | --- | --- | --- | --- | --- |
| High-level cause | year | ASRw per 100k persons | ASRuc per 100k persons | UC mentions | ratio ASRw to ASRuc |
| Blood diseases | 2019 | 7.46 (6.88 - 8.03) | 3.31 (2.92 - 3.69) | 293 | 2.26 (1.96 - 2.59) |
| Blood diseases | 2020-2023 avg | 8.86 (8.25 - 9.48) | 3.51 (3.12 - 3.89) | 319 | 2.65 (2.31 - 3.03) |
| Blood diseases | 2024 | 8.46 (7.87 - 9.06) | 2.86 (2.52 - 3.2) | 271 | 2.96 (2.57 - 3.4) |
| Cardiovascular diseases | 2019 | 247.5 (244.18 - 250.83) | 345.53 (341.6 - 349.46) | 30129 | 0.72 (0.7 - 0.73) |
| Cardiovascular diseases | 2020-2023 avg | 247.54 (244.29 - 250.8) | 326.94 (323.2 - 330.69) | 29646 | 0.76 (0.74 - 0.77) |
| Cardiovascular diseases | 2024 | 224.25 (221.21 - 227.3) | 298.1 (294.59 - 301.6) | 28108 | 0.75 (0.74 - 0.77) |
| Congenital conditions | 2019 | 1.88 (1.6 - 2.17) | 2.93 (2.57 - 3.28) | 260 | 0.64 (0.53 - 0.78) |
| Congenital conditions | 2020-2023 avg | 1.9 (1.62 - 2.19) | 2.87 (2.52 - 3.22) | 260 | 0.66 (0.55 - 0.8) |
| Congenital conditions | 2024 | 2.11 (1.81 - 2.41) | 3.09 (2.72 - 3.45) | 276 | 0.68 (0.57 - 0.82) |
| Digestive diseases | 2019 | 33 (31.79 - 34.22) | 27.77 (26.66 - 28.88) | 2422 | 1.19 (1.13 - 1.25) |
| Digestive diseases | 2020-2023 avg | 32.61 (31.43 - 33.8) | 27.52 (26.44 - 28.61) | 2492 | 1.19 (1.12 - 1.25) |
| Digestive diseases | 2024 | 31.37 (30.23 - 32.51) | 26.89 (25.83 - 27.94) | 2527 | 1.17 (1.11 - 1.23) |
| Endocrine disorders | 2019 | 55.49 (53.91 - 57.07) | 41.04 (39.69 - 42.4) | 3553 | 1.35 (1.29 - 1.41) |
| Endocrine disorders | 2020-2023 avg | 63.05 (61.4 - 64.7) | 49.43 (47.97 - 50.89) | 4474 | 1.28 (1.23 - 1.33) |
| Endocrine disorders | 2024 | 59.53 (57.97 - 61.1) | 47.68 (46.28 - 49.08) | 4504 | 1.25 (1.2 - 1.3) |
| External causes | 2019 | 27.53 (26.43 - 28.63) | 52.48 (50.96 - 54) | 4614 | 0.52 (0.5 - 0.55) |
| External causes | 2020-2023 avg | 30.09 (28.95 - 31.22) | 56.85 (55.29 - 58.41) | 5145 | 0.53 (0.5 - 0.55) |
| External causes | 2024 | 30.8 (29.67 - 31.93) | 57.92 (56.37 - 59.47) | 5410 | 0.53 (0.51 - 0.56) |
| Factors influencing health status and contact with health services | 2019 | 15.68 (14.84 - 16.52) | 0 (0 - 0) | 0 |  |
| Factors influencing health status and contact with health services | 2020-2023 avg | 16.65 (15.8 - 17.49) | 0 (0 - 0) | 0 |  |
| Factors influencing health status and contact with health services | 2024 | 15.43 (14.63 - 16.23) | 0 (0 - 0) | 0 |  |
| Genitourinary diseases | 2019 | 49.65 (48.17 - 51.14) | 21.08 (20.11 - 22.05) | 1848 | 2.36 (2.23 - 2.49) |
| Genitourinary diseases | 2020-2023 avg | 53.73 (52.22 - 55.25) | 21.3 (20.35 - 22.25) | 1930 | 2.55 (2.42 - 2.7) |
| Genitourinary diseases | 2024 | 51.03 (49.57 - 52.48) | 19.86 (18.95 - 20.76) | 1866 | 2.57 (2.43 - 2.71) |
| Hearing and vision diseases | 2019 | 0.58 (0.42 - 0.74) | 0.01 (-0.01 - 0.04) | 1 | 48.32 (6.68 - 349.76) |
| Hearing and vision diseases | 2020-2023 avg | 0.72 (0.55 - 0.9) | 0.01 (-0.01 - 0.03) | 1 | 53.09 (8.63 - 344.54) |
| Hearing and vision diseases | 2024 | 0.73 (0.56 - 0.91) | 0.03 (0 - 0.06) | 3 | 26.52 (8.25 - 85.24) |
| Infectious diseases | 2019 | 19.99 (19.05 - 20.94) | 11.75 (11.03 - 12.48) | 1032 | 1.7 (1.57 - 1.84) |
| Infectious diseases | 2020-2023 avg | 26.23 (25.17 - 27.29) | 11.14 (10.45 - 11.83) | 1010 | 2.38 (2.21 - 2.56) |
| Infectious diseases | 2024 | 25.68 (24.65 - 26.71) | 12.76 (12.03 - 13.49) | 1195 | 2.01 (1.88 - 2.16) |
| Injuries | 2019 | 9.11 (8.48 - 9.75) | 0 (0 - 0) | 0 |  |
| Injuries | 2020-2023 avg | 8.93 (8.31 - 9.55) | 0 (0 - 0) | 0 |  |
| Injuries | 2024 | 9.97 (9.33 - 10.62) | 0 (0 - 0) | 0 |  |
| Maternal conditions | 2019 | 0.1 (0.03 - 0.16) | 0.06 (0.01 - 0.11) | 5 | 1.71 (0.57 - 5.14) |
| Maternal conditions | 2020-2023 avg | 0.04 (0 - 0.08) | 0.03 (-0.01 - 0.06) | 2 | 1.54 (0.3 - 8.01) |
| Maternal conditions | 2024 | 0.1 (0.03 - 0.16) | 0.02 (-0.01 - 0.05) | 2 | 4.38 (0.93 - 20.62) |
| Mental & behavioural disorders | 2019 | 16.42 (15.57 - 17.28) | 14.79 (13.98 - 15.6) | 1295 | 1.11 (1.03 - 1.2) |
| Mental & behavioural disorders | 2020-2023 avg | 19.6 (18.68 - 20.52) | 17.38 (16.52 - 18.24) | 1572 | 1.13 (1.05 - 1.21) |
| Mental & behavioural disorders | 2024 | 20.55 (19.63 - 21.47) | 18.45 (17.58 - 19.33) | 1726 | 1.11 (1.04 - 1.19) |
| Musculoskeletal conditions | 2019 | 8.96 (8.33 - 9.59) | 4.22 (3.78 - 4.65) | 368 | 2.13 (1.88 - 2.41) |
| Musculoskeletal conditions | 2020-2023 avg | 9.47 (8.84 - 10.11) | 4.13 (3.71 - 4.55) | 376 | 2.32 (2.05 - 2.62) |
| Musculoskeletal conditions | 2024 | 9.53 (8.9 - 10.16) | 4.37 (3.95 - 4.8) | 412 | 2.18 (1.94 - 2.45) |
| Neoplasms | 2019 | 131.35 (128.93 - 133.77) | 245.08 (241.77 - 248.39) | 21325 | 0.54 (0.52 - 0.55) |
| Neoplasms | 2020-2023 avg | 130.64 (128.27 - 133.01) | 241.13 (237.92 - 244.35) | 21842 | 0.54 (0.53 - 0.55) |
| Neoplasms | 2024 | 126.89 (124.6 - 129.18) | 237.32 (234.19 - 240.44) | 22431 | 0.53 (0.52 - 0.55) |
| Nervous system diseases | 2019 | 72.52 (70.72 - 74.32) | 68.73 (66.97 - 70.48) | 5990 | 1.06 (1.02 - 1.09) |
| Nervous system diseases | 2020-2023 avg | 75.92 (74.12 - 77.73) | 70.1 (68.37 - 71.83) | 6365 | 1.09 (1.05 - 1.12) |
| Nervous system diseases | 2024 | 74.21 (72.46 - 75.96) | 78.07 (76.27 - 79.86) | 7359 | 0.95 (0.92 - 0.98) |
| Perinatal conditions | 2019 | 1.16 (0.94 - 1.38) | 1.76 (1.49 - 2.03) | 161 | 0.66 (0.51 - 0.84) |
| Perinatal conditions | 2020-2023 avg | 1.06 (0.84 - 1.27) | 1.57 (1.31 - 1.83) | 141 | 0.67 (0.52 - 0.87) |
| Perinatal conditions | 2024 | 1.19 (0.95 - 1.42) | 1.64 (1.36 - 1.91) | 135 | 0.73 (0.56 - 0.94) |
| Respiratory diseases | 2019 | 88.97 (86.97 - 90.96) | 60.72 (59.07 - 62.37) | 5283 | 1.47 (1.41 - 1.52) |
| Respiratory diseases | 2020-2023 avg | 110.91 (108.73 - 113.09) | 115.91 (113.68 - 118.14) | 10482 | 0.97 (0.94 - 0.99) |
| Respiratory diseases | 2024 | 86.95 (85.05 - 88.84) | 71.05 (69.34 - 72.77) | 6701 | 1.22 (1.18 - 1.26) |
| Skin diseases | 2019 | 3.92 (3.5 - 4.34) | 1.1 (0.88 - 1.33) | 94 | 3.55 (2.82 - 4.47) |
| Skin diseases | 2020-2023 avg | 3.84 (3.43 - 4.24) | 0.97 (0.77 - 1.17) | 88 | 4.14 (3.24 - 5.29) |
| Skin diseases | 2024 | 3.81 (3.41 - 4.2) | 1.01 (0.81 - 1.21) | 95 | 3.77 (3 - 4.73) |

| Table S8: Top 10 causes most co-occurring with COVID-19 by year | | | | | |
| --- | --- | --- | --- | --- | --- |
| Rank | 2024 | 2023 | 2022 | 2021 | 2020 |
| 1 | Pneumonia | Pneumonia | Pneumonia | Pneumonia | Residual - infections |
| 2 | Residual - infections | Residual - infections | Residual - infections | Residual - infections | Pneumonia |
| 3 | Hypertensive diseases | Hypertensive diseases | Hypertensive diseases | Hypertensive diseases | Hypertensive diseases |
| 4 | Renal failure | Ischaemic heart disease | Renal failure | Renal failure | Renal failure |
| 5 | Ischaemic heart disease | Renal failure | Ischaemic heart disease | Ischaemic heart disease | Ischaemic heart disease |
| 6 | Dementia & Alzheimer disease | Diabetes mellitus | Dementia & Alzheimer disease | Dementia & Alzheimer disease | Dementia & Alzheimer disease |
| 7 | Diabetes mellitus | Other heart diseases | Other heart diseases | Other heart diseases | Atrial fibrillation |
| 8 | Other heart diseases | Dementia & Alzheimer disease | Diabetes mellitus | Atrial fibrillation | Diabetes mellitus |
| 9 | Artery diseases | Atrial fibrillation | Atrial fibrillation | Diabetes mellitus | Other heart diseases |
| 10 | Atrial fibrillation | Artery diseases | Artery diseases | Artery diseases | Cerebrovascular disease |

#### **Sensitivity analyses**

##### *Alternative weighting*

| Table S9: Outcomes of different weighting schemes by cause and year | | | | | | | | | | | | | | | | | | |
| --- | --- | --- | --- | --- | --- | --- | --- | --- | --- | --- | --- | --- | --- | --- | --- | --- | --- | --- |
|  | 2019 (95% CI) | | | 2020 (95% CI) | | | 2021 (95% CI) | | | 2022 (95% CI) | | | 2023 (95% CI) | | | 2024 (95% CI) | | |
| Cause | ASRmain | ASRequal | ASRdouble | ASRmain | ASRequal | ASRdouble | ASRmain | ASRequal | ASRdouble | ASRmain | ASRequal | ASRdouble | ASRmain | ASRequal | ASRdouble | ASRmain | ASRequal | ASRdouble |
| **Infectious diseases** | 19.994 (19.05 - 20.938) | 22.235 (21.239 - 23.232) | 19.599 (18.664 - 20.534) | 26.713 (25.63 - 27.796) | 31.467 (30.292 - 32.642) | 26.277 (25.203 - 27.35) | 26.616 (25.543 - 27.688) | 31.431 (30.266 - 32.597) | 26.555 (25.483 - 27.626) | 26.42 (25.359 - 27.481) | 31.575 (30.416 - 32.735) | 26.575 (25.511 - 27.639) | 25.182 (24.151 - 26.212) | 29.287 (28.176 - 30.398) | 25.387 (24.353 - 26.422) | 25.683 (24.652 - 26.713) | 29.937 (28.824 - 31.049) | 25.892 (24.858 - 26.927) |
| HIV disease | 0.247 (0.143 - 0.351) | 0.163 (0.078 - 0.247) | 0.215 (0.118 - 0.311) | 0.224 (0.126 - 0.322) | 0.142 (0.064 - 0.221) | 0.191 (0.1 - 0.282) | 0.238 (0.137 - 0.339) | 0.164 (0.08 - 0.248) | 0.202 (0.109 - 0.295) | 0.244 (0.142 - 0.345) | 0.163 (0.08 - 0.247) | 0.202 (0.11 - 0.295) | 0.277 (0.169 - 0.386) | 0.196 (0.106 - 0.287) | 0.24 (0.14 - 0.341) | 0.204 (0.112 - 0.297) | 0.147 (0.068 - 0.225) | 0.184 (0.096 - 0.272) |
| Intestinal infections | 1.482 (1.224 - 1.74) | 1.445 (1.19 - 1.7) | 1.507 (1.247 - 1.766) | 1.502 (1.244 - 1.759) | 1.434 (1.183 - 1.685) | 1.495 (1.239 - 1.751) | 1.604 (1.341 - 1.868) | 1.454 (1.203 - 1.705) | 1.571 (1.31 - 1.831) | 1.464 (1.215 - 1.714) | 1.416 (1.17 - 1.662) | 1.48 (1.229 - 1.731) | 1.564 (1.307 - 1.822) | 1.352 (1.113 - 1.592) | 1.492 (1.241 - 1.744) | 1.855 (1.578 - 2.132) | 1.628 (1.368 - 1.887) | 1.794 (1.522 - 2.067) |
| Residual - infections | 5.088 (4.612 - 5.564) | 5.729 (5.223 - 6.235) | 5.285 (4.799 - 5.771) | 10.51 (9.832 - 11.189) | 13.215 (12.454 - 13.976) | 11.118 (10.419 - 11.816) | 9.718 (9.071 - 10.366) | 12.486 (11.752 - 13.221) | 10.717 (10.037 - 11.397) | 9.453 (8.818 - 10.087) | 12.372 (11.647 - 13.097) | 10.544 (9.875 - 11.214) | 8.26 (7.669 - 8.851) | 10.183 (9.527 - 10.838) | 9.082 (8.463 - 9.701) | 8.694 (8.095 - 9.294) | 10.759 (10.092 - 11.425) | 9.562 (8.934 - 10.191) |
| Septicaemia | 8.993 (8.36 - 9.627) | 10.894 (10.197 - 11.592) | 9.343 (8.697 - 9.989) | 8.743 (8.124 - 9.363) | 11.048 (10.352 - 11.744) | 9.266 (8.628 - 9.903) | 9.181 (8.551 - 9.812) | 11.418 (10.715 - 12.121) | 9.848 (9.196 - 10.501) | 9.666 (9.024 - 10.309) | 12.04 (11.323 - 12.756) | 10.325 (9.661 - 10.988) | 10.285 (9.627 - 10.943) | 12.684 (11.954 - 13.414) | 10.965 (10.285 - 11.644) | 10.233 (9.583 - 10.884) | 12.602 (11.88 - 13.324) | 10.949 (10.276 - 11.622) |
| Tuberculosis | 0.331 (0.211 - 0.451) | 0.307 (0.191 - 0.423) | 0.337 (0.216 - 0.458) | 0.338 (0.215 - 0.46) | 0.316 (0.197 - 0.434) | 0.352 (0.227 - 0.476) | 0.297 (0.183 - 0.412) | 0.279 (0.168 - 0.389) | 0.29 (0.177 - 0.403) | 0.268 (0.162 - 0.375) | 0.232 (0.132 - 0.331) | 0.269 (0.162 - 0.375) | 0.279 (0.17 - 0.388) | 0.231 (0.132 - 0.329) | 0.261 (0.156 - 0.366) | 0.21 (0.118 - 0.303) | 0.187 (0.1 - 0.275) | 0.2 (0.11 - 0.29) |
| Viral hepatitis | 1.361 (1.115 - 1.607) | 1.117 (0.894 - 1.34) | 1.209 (0.977 - 1.44) | 1.223 (0.992 - 1.454) | 1.077 (0.861 - 1.294) | 1.156 (0.931 - 1.38) | 0.919 (0.72 - 1.119) | 0.902 (0.705 - 1.099) | 0.912 (0.713 - 1.11) | 0.932 (0.732 - 1.131) | 0.896 (0.701 - 1.092) | 0.925 (0.726 - 1.124) | 0.883 (0.69 - 1.076) | 0.824 (0.638 - 1.01) | 0.867 (0.676 - 1.058) | 0.764 (0.586 - 0.941) | 0.795 (0.613 - 0.976) | 0.755 (0.578 - 0.932) |
| **Neoplasms** | 131.351 (128.928 - 133.775) | 112.803 (110.558 - 115.049) | 149.7 (147.113 - 152.287) | 132.621 (130.21 - 135.032) | 113.373 (111.145 - 115.602) | 150.003 (147.439 - 152.566) | 130.645 (128.268 - 133.023) | 111.626 (109.429 - 113.824) | 146.786 (144.266 - 149.306) | 131.239 (128.872 - 133.606) | 110.471 (108.3 - 112.642) | 146.278 (143.78 - 148.776) | 128.067 (125.749 - 130.386) | 108.068 (105.938 - 110.198) | 143.153 (140.702 - 145.605) | 126.891 (124.605 - 129.178) | 106.167 (104.075 - 108.259) | 141.737 (139.32 - 144.154) |
| Bladder cancer | 3.6 (3.201 - 4) | 2.603 (2.263 - 2.942) | 3.42 (3.03 - 3.809) | 3.522 (3.13 - 3.913) | 2.517 (2.186 - 2.848) | 3.317 (2.937 - 3.698) | 3.624 (3.226 - 4.022) | 2.619 (2.281 - 2.957) | 3.422 (3.035 - 3.808) | 3.744 (3.344 - 4.143) | 2.686 (2.347 - 3.025) | 3.48 (3.094 - 3.865) | 3.403 (3.026 - 3.78) | 2.418 (2.099 - 2.736) | 3.179 (2.815 - 3.544) | 3.104 (2.747 - 3.461) | 2.157 (1.859 - 2.454) | 2.885 (2.541 - 3.229) |
| Brain cancer | 3.274 (2.894 - 3.654) | 2.724 (2.378 - 3.071) | 3.748 (3.342 - 4.155) | 3.551 (3.156 - 3.945) | 2.898 (2.541 - 3.254) | 3.993 (3.575 - 4.412) | 3.277 (2.901 - 3.654) | 2.633 (2.295 - 2.971) | 3.648 (3.251 - 4.045) | 3.633 (3.239 - 4.028) | 2.923 (2.57 - 3.277) | 4.027 (3.612 - 4.442) | 3.268 (2.899 - 3.638) | 2.627 (2.296 - 2.958) | 3.615 (3.227 - 4.004) | 3.237 (2.872 - 3.603) | 2.597 (2.269 - 2.925) | 3.589 (3.204 - 3.974) |
| Breast cancer | 11.063 (10.361 - 11.766) | 8.893 (8.263 - 9.523) | 11.411 (10.697 - 12.124) | 10.885 (10.194 - 11.575) | 8.579 (7.967 - 9.191) | 11.004 (10.311 - 11.698) | 10.643 (9.964 - 11.321) | 8.448 (7.844 - 9.053) | 10.824 (10.14 - 11.508) | 10.363 (9.698 - 11.028) | 8.084 (7.496 - 8.671) | 10.289 (9.626 - 10.951) | 10.575 (9.908 - 11.242) | 8.329 (7.737 - 8.92) | 10.529 (9.864 - 11.193) | 10.964 (10.291 - 11.637) | 8.434 (7.843 - 9.026) | 10.855 (10.184 - 11.525) |
| Cancer secondary site | 31.652 (30.463 - 32.841) | 38.419 (37.109 - 39.729) | 28.285 (27.16 - 29.409) | 31.136 (29.969 - 32.304) | 38.155 (36.862 - 39.448) | 28.242 (27.13 - 29.354) | 29.866 (28.729 - 31.003) | 36.668 (35.408 - 37.928) | 27.167 (26.082 - 28.251) | 28.728 (27.621 - 29.835) | 35.353 (34.126 - 36.581) | 26.232 (25.175 - 27.29) | 27.412 (26.339 - 28.484) | 33.983 (32.789 - 35.177) | 25.29 (24.26 - 26.32) | 27.118 (26.061 - 28.175) | 34.11 (32.925 - 35.295) | 25.542 (24.516 - 26.567) |
| Cancer unknown primary | 15.211 (14.387 - 16.036) | 16.832 (15.965 - 17.699) | 14.485 (13.681 - 15.289) | 16.279 (15.436 - 17.123) | 18.116 (17.227 - 19.005) | 15.471 (14.649 - 16.292) | 15.507 (14.688 - 16.325) | 17.321 (16.456 - 18.186) | 14.999 (14.194 - 15.804) | 14.862 (14.066 - 15.657) | 16.488 (15.65 - 17.327) | 14.349 (13.567 - 15.131) | 15.027 (14.232 - 15.821) | 16.938 (16.094 - 17.781) | 14.612 (13.829 - 15.395) | 16.779 (15.948 - 17.61) | 18.949 (18.066 - 19.831) | 16.236 (15.418 - 17.054) |
| Cervical cancer | 0.929 (0.725 - 1.133) | 0.696 (0.52 - 0.872) | 0.948 (0.743 - 1.154) | 0.898 (0.7 - 1.097) | 0.674 (0.502 - 0.846) | 0.898 (0.699 - 1.096) | 0.802 (0.617 - 0.988) | 0.625 (0.461 - 0.79) | 0.826 (0.638 - 1.015) | 0.752 (0.573 - 0.93) | 0.526 (0.377 - 0.676) | 0.726 (0.55 - 0.901) | 0.806 (0.623 - 0.99) | 0.581 (0.425 - 0.737) | 0.785 (0.604 - 0.967) | 0.806 (0.623 - 0.989) | 0.59 (0.434 - 0.746) | 0.796 (0.615 - 0.978) |
| Colorectal cancer | 13.224 (12.457 - 13.991) | 9.635 (8.98 - 10.29) | 12.905 (12.148 - 13.663) | 13.213 (12.453 - 13.973) | 9.428 (8.786 - 10.07) | 12.713 (11.967 - 13.458) | 12.772 (12.028 - 13.516) | 9.221 (8.59 - 9.853) | 12.312 (11.582 - 13.042) | 12.06 (11.344 - 12.777) | 8.653 (8.046 - 9.259) | 11.621 (10.918 - 12.325) | 12.329 (11.61 - 13.048) | 8.75 (8.145 - 9.355) | 11.832 (11.128 - 12.536) | 12.027 (11.323 - 12.731) | 8.362 (7.775 - 8.949) | 11.424 (10.738 - 12.109) |
| Gallbladder cancer | 1.564 (1.297 - 1.831) | 1.059 (0.84 - 1.279) | 1.503 (1.241 - 1.764) | 1.719 (1.446 - 1.993) | 1.202 (0.972 - 1.431) | 1.682 (1.411 - 1.953) | 1.759 (1.483 - 2.035) | 1.228 (0.998 - 1.458) | 1.722 (1.45 - 1.995) | 1.588 (1.327 - 1.849) | 1.129 (0.909 - 1.349) | 1.584 (1.324 - 1.845) | 1.319 (1.084 - 1.555) | 0.854 (0.665 - 1.044) | 1.22 (0.994 - 1.447) | 1.68 (1.417 - 1.943) | 1.177 (0.957 - 1.397) | 1.656 (1.395 - 1.916) |
| Hodgkin lymphoma | 0.265 (0.156 - 0.375) | 0.231 (0.13 - 0.333) | 0.271 (0.161 - 0.381) | 0.291 (0.178 - 0.404) | 0.226 (0.127 - 0.326) | 0.284 (0.173 - 0.395) | 0.245 (0.143 - 0.347) | 0.21 (0.116 - 0.305) | 0.247 (0.145 - 0.349) | 0.238 (0.135 - 0.342) | 0.185 (0.095 - 0.275) | 0.221 (0.122 - 0.319) | 0.264 (0.158 - 0.369) | 0.194 (0.103 - 0.284) | 0.249 (0.146 - 0.351) | 0.269 (0.164 - 0.375) | 0.184 (0.096 - 0.271) | 0.234 (0.135 - 0.332) |
| Kidney cancer | 2.759 (2.408 - 3.11) | 1.987 (1.689 - 2.285) | 2.666 (2.321 - 3.011) | 2.415 (2.088 - 2.741) | 1.736 (1.459 - 2.012) | 2.277 (1.961 - 2.593) | 2.49 (2.163 - 2.817) | 1.826 (1.546 - 2.106) | 2.379 (2.06 - 2.699) | 2.737 (2.395 - 3.078) | 1.909 (1.624 - 2.194) | 2.538 (2.21 - 2.866) | 2.405 (2.085 - 2.725) | 1.714 (1.444 - 1.984) | 2.247 (1.938 - 2.556) | 2.551 (2.226 - 2.876) | 1.761 (1.491 - 2.031) | 2.354 (2.042 - 2.667) |
| Larynx cancer | 0.913 (0.712 - 1.114) | 0.693 (0.518 - 0.869) | 0.901 (0.702 - 1.101) | 0.942 (0.739 - 1.144) | 0.734 (0.555 - 0.913) | 0.949 (0.746 - 1.153) | 0.727 (0.55 - 0.904) | 0.568 (0.412 - 0.725) | 0.725 (0.548 - 0.902) | 0.896 (0.702 - 1.09) | 0.671 (0.503 - 0.839) | 0.873 (0.682 - 1.065) | 0.856 (0.669 - 1.044) | 0.628 (0.467 - 0.789) | 0.825 (0.641 - 1.01) | 0.889 (0.697 - 1.08) | 0.628 (0.467 - 0.789) | 0.822 (0.638 - 1.006) |
| Liver cancer | 5.144 (4.663 - 5.624) | 3.481 (3.085 - 3.876) | 4.964 (4.492 - 5.436) | 4.914 (4.448 - 5.38) | 3.24 (2.862 - 3.618) | 4.648 (4.195 - 5.101) | 4.899 (4.439 - 5.359) | 3.168 (2.798 - 3.538) | 4.531 (4.089 - 4.973) | 5.017 (4.555 - 5.479) | 3.212 (2.842 - 3.582) | 4.589 (4.147 - 5.031) | 4.573 (4.137 - 5.008) | 2.916 (2.567 - 3.264) | 4.182 (3.765 - 4.599) | 4.562 (4.129 - 4.996) | 2.839 (2.497 - 3.181) | 4.135 (3.722 - 4.548) |
| Lung cancer | 23.482 (22.457 - 24.507) | 16.445 (15.588 - 17.303) | 23.31 (22.289 - 24.331) | 23.477 (22.462 - 24.492) | 16.154 (15.313 - 16.996) | 22.899 (21.897 - 23.901) | 23.319 (22.316 - 24.323) | 15.925 (15.096 - 16.754) | 22.553 (21.566 - 23.539) | 23.18 (22.185 - 24.175) | 15.582 (14.766 - 16.397) | 22.209 (21.235 - 23.183) | 22.774 (21.796 - 23.752) | 15.146 (14.348 - 15.943) | 21.702 (20.748 - 22.657) | 22.729 (21.763 - 23.694) | 14.643 (13.868 - 15.419) | 21.143 (20.212 - 22.074) |
| Malignant melanoma-skin | 2.363 (2.037 - 2.69) | 1.85 (1.561 - 2.138) | 2.446 (2.114 - 2.778) | 2.485 (2.155 - 2.815) | 1.841 (1.557 - 2.125) | 2.431 (2.105 - 2.757) | 2.483 (2.153 - 2.812) | 1.931 (1.641 - 2.222) | 2.53 (2.197 - 2.863) | 2.422 (2.101 - 2.743) | 1.775 (1.501 - 2.05) | 2.357 (2.041 - 2.673) | 2.475 (2.153 - 2.797) | 1.852 (1.574 - 2.13) | 2.446 (2.126 - 2.766) | 2.488 (2.167 - 2.808) | 1.806 (1.533 - 2.079) | 2.405 (2.09 - 2.72) |
| Mesothelioma | 0.61 (0.445 - 0.775) | 0.463 (0.32 - 0.607) | 0.644 (0.475 - 0.814) | 0.56 (0.403 - 0.718) | 0.405 (0.272 - 0.539) | 0.574 (0.415 - 0.732) | 0.562 (0.407 - 0.717) | 0.392 (0.262 - 0.522) | 0.555 (0.401 - 0.71) | 0.511 (0.362 - 0.659) | 0.34 (0.219 - 0.461) | 0.481 (0.337 - 0.625) | 0.536 (0.385 - 0.687) | 0.376 (0.249 - 0.502) | 0.531 (0.381 - 0.682) | 0.528 (0.381 - 0.675) | 0.374 (0.251 - 0.497) | 0.531 (0.384 - 0.678) |
| Non-Hodgkin lymphomas | 3.828 (3.415 - 4.241) | 2.892 (2.533 - 3.251) | 3.819 (3.406 - 4.231) | 3.87 (3.457 - 4.284) | 2.963 (2.602 - 3.324) | 3.898 (3.483 - 4.312) | 3.814 (3.41 - 4.219) | 2.886 (2.534 - 3.238) | 3.761 (3.359 - 4.163) | 3.745 (3.346 - 4.144) | 2.727 (2.386 - 3.068) | 3.632 (3.238 - 4.025) | 4.044 (3.632 - 4.456) | 2.94 (2.588 - 3.292) | 3.936 (3.529 - 4.343) | 3.538 (3.158 - 3.918) | 2.459 (2.142 - 2.776) | 3.335 (2.966 - 3.704) |
| Non-melanoma-skin | 1.163 (0.935 - 1.391) | 0.956 (0.749 - 1.162) | 1.134 (0.909 - 1.359) | 1.131 (0.911 - 1.351) | 0.913 (0.715 - 1.111) | 1.089 (0.872 - 1.305) | 1.233 (1.002 - 1.464) | 1.046 (0.834 - 1.259) | 1.194 (0.967 - 1.421) | 1.246 (1.017 - 1.476) | 1.01 (0.803 - 1.217) | 1.19 (0.965 - 1.414) | 1.195 (0.971 - 1.42) | 0.988 (0.784 - 1.193) | 1.162 (0.94 - 1.383) | 1.266 (1.036 - 1.496) | 1.002 (0.798 - 1.206) | 1.199 (0.975 - 1.422) |
| Oesophagus cancer | 2.174 (1.863 - 2.485) | 1.573 (1.309 - 1.837) | 2.195 (1.883 - 2.508) | 2.501 (2.17 - 2.833) | 1.806 (1.524 - 2.087) | 2.508 (2.177 - 2.84) | 2.305 (1.988 - 2.621) | 1.612 (1.347 - 1.876) | 2.237 (1.925 - 2.548) | 2.309 (1.993 - 2.624) | 1.573 (1.312 - 1.833) | 2.192 (1.885 - 2.5) | 2.22 (1.915 - 2.525) | 1.54 (1.286 - 1.795) | 2.148 (1.847 - 2.448) | 2.142 (1.845 - 2.439) | 1.428 (1.185 - 1.671) | 2.025 (1.736 - 2.314) |
| Oral cancers | 3.389 (3.001 - 3.777) | 2.443 (2.114 - 2.772) | 3.337 (2.952 - 3.721) | 3.256 (2.879 - 3.633) | 2.421 (2.096 - 2.746) | 3.248 (2.871 - 3.624) | 3.352 (2.972 - 3.731) | 2.372 (2.052 - 2.691) | 3.244 (2.871 - 3.618) | 3.471 (3.087 - 3.855) | 2.477 (2.152 - 2.802) | 3.363 (2.984 - 3.741) | 3.084 (2.726 - 3.443) | 2.205 (1.901 - 2.508) | 2.992 (2.639 - 3.346) | 3.088 (2.733 - 3.443) | 2.141 (1.846 - 2.437) | 2.938 (2.592 - 3.284) |
| Other blood cancers | 9.807 (9.143 - 10.472) | 7.35 (6.776 - 7.925) | 9.736 (9.074 - 10.397) | 9.935 (9.273 - 10.596) | 7.519 (6.943 - 8.094) | 9.929 (9.267 - 10.59) | 9.578 (8.932 - 10.224) | 7.306 (6.742 - 7.869) | 9.445 (8.804 - 10.086) | 10.226 (9.564 - 10.887) | 7.594 (7.024 - 8.164) | 10.054 (9.399 - 10.71) | 9.33 (8.704 - 9.956) | 6.88 (6.342 - 7.418) | 9.085 (8.467 - 9.703) | 9.269 (8.65 - 9.887) | 6.624 (6.101 - 7.146) | 8.84 (8.236 - 9.444) |
| Ovarian cancer | 2.918 (2.556 - 3.279) | 2.157 (1.847 - 2.467) | 2.995 (2.63 - 3.361) | 2.881 (2.524 - 3.237) | 2.091 (1.788 - 2.394) | 2.925 (2.566 - 3.284) | 2.501 (2.173 - 2.829) | 1.765 (1.489 - 2.04) | 2.451 (2.126 - 2.776) | 2.766 (2.424 - 3.108) | 1.967 (1.678 - 2.256) | 2.734 (2.394 - 3.075) | 2.575 (2.247 - 2.903) | 1.811 (1.537 - 2.086) | 2.54 (2.215 - 2.866) | 2.781 (2.443 - 3.12) | 2.001 (1.714 - 2.288) | 2.792 (2.453 - 3.131) |
| Pancreatic cancer | 10.439 (9.756 - 11.123) | 7.397 (6.822 - 7.972) | 10.567 (9.879 - 11.254) | 10.596 (9.916 - 11.277) | 7.499 (6.926 - 8.072) | 10.681 (9.997 - 11.365) | 10.535 (9.86 - 11.211) | 7.356 (6.791 - 7.92) | 10.5 (9.826 - 11.175) | 10.596 (9.923 - 11.269) | 7.445 (6.881 - 8.009) | 10.571 (9.899 - 11.243) | 10.876 (10.2 - 11.551) | 7.655 (7.089 - 8.222) | 10.853 (10.178 - 11.528) | 11.031 (10.356 - 11.706) | 7.58 (7.02 - 8.139) | 10.858 (10.189 - 11.528) |
| Prostate cancer | 9.285 (8.64 - 9.931) | 7.014 (6.453 - 7.575) | 8.898 (8.266 - 9.53) | 9.384 (8.742 - 10.025) | 6.977 (6.424 - 7.53) | 8.85 (8.227 - 9.473) | 9.005 (8.38 - 9.629) | 6.887 (6.341 - 7.433) | 8.658 (8.046 - 9.271) | 9.404 (8.77 - 10.038) | 7.136 (6.584 - 7.689) | 8.975 (8.355 - 9.594) | 9.233 (8.609 - 9.857) | 6.974 (6.432 - 7.516) | 8.82 (8.21 - 9.429) | 8.911 (8.305 - 9.518) | 6.452 (5.937 - 6.968) | 8.283 (7.699 - 8.868) |
| Residual - benign/in situ/uncertain neoplasms | 4.474 (4.027 - 4.922) | 4.267 (3.83 - 4.704) | 4.428 (3.983 - 4.874) | 4.826 (4.367 - 5.285) | 4.668 (4.216 - 5.119) | 4.843 (4.383 - 5.303) | 4.803 (4.347 - 5.258) | 4.649 (4.201 - 5.097) | 4.792 (4.337 - 5.248) | 5.494 (5.008 - 5.979) | 5.266 (4.792 - 5.74) | 5.522 (5.035 - 6.008) | 5.461 (4.981 - 5.941) | 5.345 (4.87 - 5.819) | 5.493 (5.011 - 5.974) | 6.195 (5.689 - 6.7) | 6.326 (5.815 - 6.837) | 6.132 (5.629 - 6.635) |
| Residual - malignant neoplasms | 6.261 (5.731 - 6.791) | 4.689 (4.231 - 5.148) | 6.275 (5.744 - 6.805) | 6.544 (6.008 - 7.079) | 4.868 (4.405 - 5.331) | 6.483 (5.949 - 7.017) | 6.819 (6.275 - 7.362) | 4.984 (4.519 - 5.449) | 6.684 (6.146 - 7.223) | 6.576 (6.046 - 7.106) | 4.787 (4.334 - 5.239) | 6.401 (5.878 - 6.924) | 6.837 (6.3 - 7.374) | 4.995 (4.536 - 5.454) | 6.63 (6.101 - 7.159) | 6.334 (5.823 - 6.846) | 4.53 (4.098 - 4.962) | 6.058 (5.558 - 6.558) |
| Stomach cancer | 4.421 (3.975 - 4.867) | 3.212 (2.831 - 3.592) | 4.465 (4.016 - 4.913) | 4.46 (4.018 - 4.903) | 3.167 (2.794 - 3.54) | 4.402 (3.962 - 4.842) | 4.099 (3.677 - 4.522) | 2.948 (2.59 - 3.306) | 4.062 (3.642 - 4.483) | 4.087 (3.67 - 4.504) | 2.866 (2.517 - 3.215) | 3.972 (3.561 - 4.383) | 4.024 (3.612 - 4.435) | 2.839 (2.493 - 3.186) | 3.935 (3.527 - 4.342) | 3.865 (3.466 - 4.264) | 2.652 (2.321 - 2.982) | 3.699 (3.309 - 4.089) |
| Thyroid cancer | 0.629 (0.461 - 0.797) | 0.467 (0.323 - 0.612) | 0.618 (0.452 - 0.784) | 0.545 (0.39 - 0.7) | 0.414 (0.279 - 0.548) | 0.54 (0.386 - 0.693) | 0.552 (0.396 - 0.707) | 0.439 (0.3 - 0.577) | 0.549 (0.394 - 0.704) | 0.473 (0.332 - 0.614) | 0.376 (0.25 - 0.502) | 0.475 (0.333 - 0.616) | 0.48 (0.339 - 0.62) | 0.362 (0.24 - 0.485) | 0.45 (0.314 - 0.586) | 0.497 (0.354 - 0.64) | 0.382 (0.257 - 0.508) | 0.477 (0.336 - 0.617) |
| Uterus cancer | 1.824 (1.537 - 2.11) | 1.417 (1.165 - 1.67) | 1.861 (1.572 - 2.15) | 1.817 (1.535 - 2.099) | 1.333 (1.092 - 1.575) | 1.782 (1.502 - 2.061) | 1.846 (1.565 - 2.127) | 1.351 (1.11 - 1.592) | 1.794 (1.517 - 2.071) | 1.788 (1.51 - 2.066) | 1.301 (1.064 - 1.537) | 1.738 (1.464 - 2.012) | 1.661 (1.397 - 1.926) | 1.176 (0.954 - 1.399) | 1.592 (1.333 - 1.851) | 1.685 (1.42 - 1.95) | 1.191 (0.968 - 1.414) | 1.62 (1.36 - 1.88) |
| **Blood diseases** | 7.459 (6.884 - 8.035) | 8.948 (8.318 - 9.579) | 7.666 (7.082 - 8.249) | 8.148 (7.551 - 8.745) | 10.14 (9.474 - 10.806) | 8.312 (7.709 - 8.915) | 8.612 (8.003 - 9.222) | 10.514 (9.841 - 11.188) | 8.934 (8.312 - 9.555) | 9.186 (8.56 - 9.812) | 11.077 (10.389 - 11.765) | 9.443 (8.808 - 10.078) | 9.501 (8.867 - 10.134) | 11.299 (10.609 - 11.99) | 9.732 (9.092 - 10.372) | 8.465 (7.874 - 9.056) | 10.461 (9.804 - 11.118) | 8.737 (8.136 - 9.338) |
| Anaemias | 4.204 (3.772 - 4.635) | 5.439 (4.948 - 5.931) | 4.735 (4.276 - 5.193) | 4.824 (4.365 - 5.283) | 6.418 (5.888 - 6.947) | 5.404 (4.918 - 5.89) | 4.768 (4.316 - 5.221) | 6.308 (5.787 - 6.829) | 5.437 (4.953 - 5.921) | 4.975 (4.515 - 5.435) | 6.575 (6.046 - 7.104) | 5.617 (5.128 - 6.107) | 5.363 (4.887 - 5.839) | 6.941 (6.4 - 7.483) | 6.015 (5.511 - 6.519) | 4.814 (4.368 - 5.259) | 6.403 (5.889 - 6.917) | 5.436 (4.963 - 5.91) |
| Residual - blood diseases | 1.881 (1.592 - 2.171) | 2.035 (1.734 - 2.337) | 1.954 (1.659 - 2.249) | 1.69 (1.418 - 1.962) | 2.023 (1.725 - 2.321) | 1.815 (1.533 - 2.096) | 2.15 (1.844 - 2.456) | 2.348 (2.029 - 2.668) | 2.259 (1.946 - 2.573) | 2.432 (2.109 - 2.755) | 2.537 (2.207 - 2.867) | 2.502 (2.174 - 2.83) | 2.353 (2.039 - 2.668) | 2.353 (2.039 - 2.668) | 2.348 (2.034 - 2.662) | 2.006 (1.718 - 2.293) | 2.282 (1.975 - 2.589) | 2.128 (1.832 - 2.424) |
| **Endocrine disorders** | 55.491 (53.914 - 57.068) | 58.804 (57.181 - 60.428) | 54.104 (52.546 - 55.661) | 61.923 (60.274 - 63.572) | 66.553 (64.844 - 68.262) | 60.651 (59.019 - 62.283) | 64.65 (62.978 - 66.322) | 68.205 (66.487 - 69.922) | 63.868 (62.206 - 65.53) | 64.332 (62.674 - 65.989) | 67.609 (65.91 - 69.308) | 63.861 (62.209 - 65.512) | 61.297 (59.692 - 62.902) | 64.103 (62.461 - 65.745) | 60.569 (58.973 - 62.164) | 59.533 (57.965 - 61.1) | 62.601 (60.993 - 64.209) | 58.777 (57.219 - 60.334) |
| Amyloidosis | 0.421 (0.281 - 0.56) | 0.326 (0.204 - 0.449) | 0.36 (0.231 - 0.489) | 0.449 (0.308 - 0.59) | 0.428 (0.291 - 0.566) | 0.458 (0.316 - 0.599) | 0.622 (0.458 - 0.786) | 0.49 (0.345 - 0.635) | 0.587 (0.428 - 0.747) | 0.746 (0.568 - 0.924) | 0.627 (0.464 - 0.79) | 0.707 (0.534 - 0.88) | 0.8 (0.617 - 0.982) | 0.609 (0.45 - 0.769) | 0.726 (0.552 - 0.9) | 0.947 (0.751 - 1.142) | 0.705 (0.537 - 0.874) | 0.87 (0.683 - 1.058) |
| Dehydration disorders | 3.613 (3.212 - 4.014) | 4.816 (4.352 - 5.28) | 3.97 (3.55 - 4.391) | 3.757 (3.353 - 4.162) | 5.124 (4.651 - 5.597) | 4.117 (3.693 - 4.54) | 4.049 (3.632 - 4.467) | 5.44 (4.956 - 5.924) | 4.503 (4.062 - 4.943) | 4.167 (3.746 - 4.587) | 5.674 (5.183 - 6.165) | 4.742 (4.293 - 5.191) | 3.991 (3.581 - 4.401) | 5.376 (4.9 - 5.852) | 4.461 (4.027 - 4.894) | 4.121 (3.709 - 4.533) | 5.514 (5.037 - 5.992) | 4.573 (4.139 - 5.008) |
| Diabetes mellitus | 29.028 (27.886 - 30.17) | 28.212 (27.086 - 29.338) | 28.659 (27.524 - 29.794) | 32.681 (31.481 - 33.88) | 32.864 (31.662 - 34.066) | 32.744 (31.544 - 33.945) | 33.821 (32.61 - 35.032) | 32.523 (31.335 - 33.711) | 33.276 (32.074 - 34.477) | 33.926 (32.722 - 35.13) | 31.799 (30.633 - 32.964) | 33.039 (31.851 - 34.227) | 31.938 (30.779 - 33.097) | 29.885 (28.764 - 31.006) | 31.053 (29.91 - 32.196) | 30.849 (29.72 - 31.978) | 29.306 (28.205 - 30.407) | 30.236 (29.118 - 31.354) |
| Disorders of thyorid gland | 1.406 (1.155 - 1.657) | 1.967 (1.67 - 2.263) | 1.683 (1.409 - 1.957) | 1.438 (1.187 - 1.688) | 2.072 (1.771 - 2.372) | 1.729 (1.454 - 2.004) | 1.445 (1.196 - 1.695) | 2.086 (1.786 - 2.386) | 1.742 (1.468 - 2.016) | 1.595 (1.335 - 1.856) | 2.367 (2.049 - 2.684) | 1.975 (1.685 - 2.265) | 1.578 (1.321 - 1.835) | 2.283 (1.974 - 2.592) | 1.925 (1.64 - 2.209) | 1.478 (1.231 - 1.726) | 2.17 (1.87 - 2.469) | 1.804 (1.53 - 2.077) |
| Malnutrition | 3.189 (2.813 - 3.565) | 4.093 (3.667 - 4.519) | 3.353 (2.967 - 3.739) | 3.18 (2.809 - 3.551) | 4.088 (3.667 - 4.509) | 3.267 (2.89 - 3.643) | 3.251 (2.878 - 3.624) | 4.127 (3.706 - 4.548) | 3.365 (2.985 - 3.745) | 3.467 (3.084 - 3.851) | 4.357 (3.927 - 4.787) | 3.61 (3.219 - 4.001) | 3.153 (2.789 - 3.516) | 3.942 (3.535 - 4.349) | 3.345 (2.97 - 3.719) | 2.387 (2.074 - 2.7) | 3.002 (2.651 - 3.353) | 2.536 (2.213 - 2.859) |
| Metabolic disorders | 3.424 (3.033 - 3.816) | 4.041 (3.616 - 4.466) | 3.717 (3.31 - 4.125) | 3.561 (3.165 - 3.957) | 4.485 (4.041 - 4.929) | 4.034 (3.613 - 4.455) | 4.143 (3.72 - 4.566) | 4.901 (4.441 - 5.36) | 4.524 (4.082 - 4.965) | 4.314 (3.886 - 4.742) | 5.173 (4.704 - 5.642) | 4.792 (4.34 - 5.243) | 4.534 (4.097 - 4.971) | 5.339 (4.865 - 5.812) | 5.001 (4.543 - 5.46) | 4.845 (4.398 - 5.292) | 6.04 (5.541 - 6.539) | 5.465 (4.991 - 5.94) |
| Obesity | 4.588 (4.135 - 5.04) | 4.182 (3.75 - 4.614) | 4.448 (4.002 - 4.894) | 5.822 (5.317 - 6.327) | 5.009 (4.54 - 5.478) | 5.548 (5.054 - 6.041) | 7.123 (6.569 - 7.678) | 6.527 (5.997 - 7.058) | 7.042 (6.491 - 7.593) | 6.229 (5.712 - 6.746) | 5.665 (5.172 - 6.158) | 6.228 (5.711 - 6.744) | 5.707 (5.218 - 6.195) | 5.257 (4.788 - 5.726) | 5.686 (5.198 - 6.173) | 5.61 (5.128 - 6.091) | 4.965 (4.512 - 5.418) | 5.431 (4.957 - 5.904) |
| Residual - endocrine | 0.497 (0.348 - 0.647) | 0.66 (0.488 - 0.832) | 0.569 (0.409 - 0.729) | 0.515 (0.365 - 0.666) | 0.72 (0.541 - 0.898) | 0.61 (0.446 - 0.774) | 0.556 (0.4 - 0.711) | 0.696 (0.522 - 0.869) | 0.623 (0.458 - 0.787) | 0.623 (0.461 - 0.785) | 0.807 (0.622 - 0.992) | 0.711 (0.538 - 0.884) | 0.713 (0.54 - 0.887) | 0.869 (0.677 - 1.061) | 0.805 (0.621 - 0.99) | 0.582 (0.427 - 0.736) | 0.799 (0.618 - 0.98) | 0.695 (0.526 - 0.863) |
| **Mental & behavioural disorders** | 16.423 (15.57 - 17.276) | 16.659 (15.799 - 17.519) | 16.13 (15.284 - 16.975) | 18.546 (17.646 - 19.446) | 18.905 (17.996 - 19.814) | 18.161 (17.27 - 19.051) | 19.475 (18.56 - 20.391) | 19.709 (18.788 - 20.63) | 19.144 (18.237 - 20.051) | 20.798 (19.856 - 21.74) | 21.257 (20.304 - 22.209) | 20.317 (19.386 - 21.248) | 19.579 (18.671 - 20.486) | 19.918 (19.002 - 20.833) | 19.244 (18.345 - 20.143) | 20.548 (19.627 - 21.47) | 20.841 (19.913 - 21.769) | 20.135 (19.223 - 21.047) |
| Alcohol induced diseases | 7.201 (6.637 - 7.766) | 4.926 (4.459 - 5.392) | 6.605 (6.064 - 7.146) | 8.033 (7.441 - 8.625) | 5.491 (5.002 - 5.98) | 7.3 (6.735 - 7.864) | 8.65 (8.04 - 9.259) | 5.885 (5.383 - 6.388) | 7.855 (7.275 - 8.436) | 9.094 (8.471 - 9.717) | 6.415 (5.892 - 6.939) | 8.349 (7.752 - 8.946) | 8.609 (8.009 - 9.21) | 5.884 (5.388 - 6.38) | 7.833 (7.261 - 8.406) | 8.764 (8.162 - 9.365) | 6.05 (5.55 - 6.549) | 7.932 (7.36 - 8.505) |
| Mood disorders | 2.939 (2.579 - 3.3) | 3.83 (3.418 - 4.242) | 2.944 (2.583 - 3.306) | 3.096 (2.728 - 3.463) | 4.102 (3.68 - 4.525) | 3.173 (2.801 - 3.545) | 2.93 (2.576 - 3.285) | 3.861 (3.454 - 4.268) | 2.985 (2.627 - 3.343) | 3.182 (2.813 - 3.551) | 4.141 (3.72 - 4.561) | 3.181 (2.813 - 3.55) | 2.954 (2.6 - 3.307) | 3.937 (3.529 - 4.345) | 3.047 (2.689 - 3.406) | 3.004 (2.651 - 3.357) | 3.99 (3.583 - 4.397) | 3.083 (2.725 - 3.441) |
| Residual - mental/behavioural | 1.54 (1.279 - 1.801) | 1.988 (1.691 - 2.285) | 1.677 (1.404 - 1.949) | 1.749 (1.472 - 2.026) | 2.302 (1.984 - 2.62) | 1.941 (1.649 - 2.233) | 1.917 (1.63 - 2.205) | 2.472 (2.146 - 2.799) | 2.111 (1.81 - 2.413) | 2.188 (1.883 - 2.494) | 2.812 (2.466 - 3.159) | 2.373 (2.055 - 2.691) | 2.114 (1.816 - 2.413) | 2.629 (2.296 - 2.962) | 2.259 (1.95 - 2.568) | 2.43 (2.112 - 2.748) | 2.952 (2.602 - 3.301) | 2.627 (2.297 - 2.957) |
| Schizophrenia | 0.813 (0.622 - 1.003) | 1.05 (0.833 - 1.266) | 0.814 (0.623 - 1.004) | 0.852 (0.66 - 1.044) | 1.146 (0.923 - 1.37) | 0.897 (0.7 - 1.095) | 0.91 (0.713 - 1.108) | 1.165 (0.942 - 1.389) | 0.925 (0.725 - 1.124) | 1.011 (0.803 - 1.219) | 1.313 (1.075 - 1.55) | 1.022 (0.813 - 1.232) | 0.889 (0.697 - 1.082) | 1.182 (0.96 - 1.404) | 0.944 (0.745 - 1.142) | 0.858 (0.669 - 1.048) | 1.148 (0.929 - 1.367) | 0.908 (0.714 - 1.103) |
| Substance use disorders | 2.732 (2.384 - 3.079) | 3.354 (2.969 - 3.74) | 2.949 (2.588 - 3.31) | 3.268 (2.891 - 3.646) | 3.934 (3.52 - 4.349) | 3.405 (3.02 - 3.791) | 3.526 (3.136 - 3.916) | 4.374 (3.94 - 4.809) | 3.795 (3.39 - 4.199) | 3.748 (3.348 - 4.148) | 4.624 (4.18 - 5.068) | 3.937 (3.528 - 4.347) | 3.76 (3.362 - 4.157) | 4.675 (4.231 - 5.118) | 3.931 (3.524 - 4.337) | 4.146 (3.732 - 4.56) | 5.025 (4.57 - 5.48) | 4.321 (3.899 - 4.743) |
| **Nervous system diseases** | 72.523 (70.725 - 74.321) | 73.6 (71.789 - 75.411) | 72.363 (70.567 - 74.159) | 77.273 (75.436 - 79.11) | 81.273 (79.389 - 83.157) | 77.59 (75.749 - 79.43) | 75.01 (73.21 - 76.81) | 77.711 (75.879 - 79.543) | 75.497 (73.691 - 77.303) | 77.432 (75.615 - 79.25) | 80.204 (78.355 - 82.054) | 78.397 (76.568 - 80.225) | 73.985 (72.219 - 75.75) | 74.737 (72.962 - 76.511) | 75.193 (73.413 - 76.974) | 74.209 (72.457 - 75.96) | 74.297 (72.545 - 76.05) | 75.59 (73.822 - 77.358) |
| Cerebral palsy | 0.226 (0.127 - 0.324) | 0.233 (0.133 - 0.333) | 0.232 (0.132 - 0.332) | 0.277 (0.167 - 0.387) | 0.264 (0.157 - 0.371) | 0.271 (0.162 - 0.38) | 0.282 (0.172 - 0.392) | 0.258 (0.153 - 0.363) | 0.284 (0.174 - 0.394) | 0.316 (0.199 - 0.434) | 0.291 (0.178 - 0.404) | 0.35 (0.226 - 0.474) | 0.33 (0.212 - 0.449) | 0.251 (0.148 - 0.354) | 0.303 (0.189 - 0.416) | 0.246 (0.143 - 0.349) | 0.196 (0.104 - 0.287) | 0.234 (0.134 - 0.334) |
| Dementia & Alzheimer disease | 40.974 (39.623 - 42.325) | 41.41 (40.053 - 42.768) | 42.823 (41.443 - 44.204) | 42.877 (41.511 - 44.242) | 45.997 (44.582 - 47.412) | 45.773 (44.362 - 47.184) | 41.963 (40.617 - 43.308) | 43.587 (42.216 - 44.959) | 44.491 (43.105 - 45.876) | 43.069 (41.714 - 44.424) | 44.83 (43.448 - 46.212) | 46.001 (44.601 - 47.401) | 42.386 (41.048 - 43.724) | 42.281 (40.945 - 43.618) | 44.777 (43.402 - 46.152) | 43.652 (42.308 - 44.996) | 42.797 (41.466 - 44.127) | 45.943 (44.565 - 47.322) |
| Epilepsy | 3.315 (2.93 - 3.699) | 3.768 (3.359 - 4.177) | 3.519 (3.123 - 3.914) | 3.716 (3.313 - 4.119) | 4.29 (3.857 - 4.723) | 4.011 (3.593 - 4.43) | 3.521 (3.13 - 3.912) | 4.184 (3.759 - 4.61) | 3.778 (3.374 - 4.182) | 3.924 (3.515 - 4.332) | 4.471 (4.035 - 4.908) | 4.161 (3.74 - 4.582) | 3.515 (3.13 - 3.899) | 3.966 (3.557 - 4.375) | 3.712 (3.317 - 4.107) | 3.503 (3.121 - 3.886) | 4.029 (3.619 - 4.438) | 3.755 (3.359 - 4.151) |
| Inflammatory diseases - CNS | 0.486 (0.34 - 0.633) | 0.459 (0.316 - 0.602) | 0.502 (0.353 - 0.651) | 0.488 (0.342 - 0.634) | 0.45 (0.31 - 0.591) | 0.487 (0.341 - 0.634) | 0.502 (0.355 - 0.648) | 0.465 (0.324 - 0.607) | 0.51 (0.363 - 0.658) | 0.545 (0.391 - 0.698) | 0.524 (0.374 - 0.675) | 0.554 (0.399 - 0.709) | 0.543 (0.392 - 0.693) | 0.527 (0.379 - 0.676) | 0.557 (0.405 - 0.71) | 0.492 (0.35 - 0.634) | 0.484 (0.344 - 0.625) | 0.502 (0.358 - 0.645) |
| Multiple sclerosis | 1.066 (0.848 - 1.285) | 0.854 (0.659 - 1.049) | 1.096 (0.875 - 1.318) | 1.098 (0.879 - 1.317) | 0.858 (0.664 - 1.052) | 1.091 (0.872 - 1.309) | 1.104 (0.885 - 1.322) | 0.879 (0.684 - 1.073) | 1.137 (0.916 - 1.359) | 1.141 (0.919 - 1.364) | 0.863 (0.671 - 1.055) | 1.127 (0.907 - 1.347) | 1.118 (0.903 - 1.334) | 0.842 (0.655 - 1.029) | 1.094 (0.881 - 1.307) | 0.842 (0.657 - 1.027) | 0.617 (0.459 - 0.776) | 0.797 (0.617 - 0.977) |
| Parkinson disease | 8.452 (7.835 - 9.069) | 7.544 (6.96 - 8.128) | 8.582 (7.959 - 9.204) | 9.21 (8.573 - 9.847) | 8.486 (7.874 - 9.098) | 9.492 (8.845 - 10.139) | 8.897 (8.275 - 9.518) | 7.928 (7.341 - 8.514) | 9.063 (8.436 - 9.69) | 9.104 (8.481 - 9.727) | 8.125 (7.536 - 8.714) | 9.246 (8.618 - 9.874) | 8.614 (8.013 - 9.216) | 7.367 (6.812 - 7.923) | 8.753 (8.147 - 9.359) | 8.718 (8.119 - 9.317) | 7.319 (6.77 - 7.868) | 8.74 (8.14 - 9.34) |
| Residual - nervous system | 9.436 (8.788 - 10.085) | 11.126 (10.422 - 11.831) | 9.402 (8.754 - 10.049) | 9.369 (8.728 - 10.01) | 11.553 (10.841 - 12.265) | 9.722 (9.069 - 10.375) | 9.174 (8.544 - 9.804) | 11.277 (10.579 - 11.975) | 9.521 (8.88 - 10.163) | 10.14 (9.481 - 10.798) | 12.221 (11.499 - 12.944) | 10.344 (9.68 - 11.009) | 9.562 (8.928 - 10.196) | 11.409 (10.716 - 12.101) | 9.749 (9.108 - 10.39) | 8.892 (8.286 - 9.497) | 10.82 (10.151 - 11.488) | 9.221 (8.604 - 9.838) |
| Systemic atrophies - CNS | 1.544 (1.282 - 1.806) | 1.256 (1.02 - 1.491) | 1.711 (1.436 - 1.987) | 1.711 (1.436 - 1.985) | 1.457 (1.204 - 1.71) | 1.944 (1.651 - 2.236) | 1.569 (1.309 - 1.828) | 1.378 (1.135 - 1.621) | 1.821 (1.542 - 2.1) | 1.636 (1.372 - 1.9) | 1.387 (1.144 - 1.63) | 1.828 (1.549 - 2.106) | 1.413 (1.171 - 1.655) | 1.211 (0.987 - 1.436) | 1.596 (1.338 - 1.854) | 1.748 (1.478 - 2.017) | 1.462 (1.215 - 1.709) | 1.925 (1.641 - 2.208) |
| **Hearing and vision diseases** | 0.584 (0.422 - 0.745) | 0.818 (0.627 - 1.009) | 0.646 (0.476 - 0.816) | 0.693 (0.52 - 0.866) | 0.985 (0.778 - 1.191) | 0.776 (0.593 - 0.96) | 0.646 (0.479 - 0.812) | 0.888 (0.693 - 1.083) | 0.697 (0.524 - 0.87) | 0.776 (0.594 - 0.958) | 1.06 (0.848 - 1.273) | 0.832 (0.644 - 1.02) | 0.775 (0.594 - 0.955) | 1.065 (0.853 - 1.277) | 0.835 (0.648 - 1.023) | 0.734 (0.56 - 0.908) | 1.001 (0.798 - 1.205) | 0.788 (0.607 - 0.968) |
| Hearing and vision diseases | 0.44 (0.3 - 0.58) | 0.65 (0.48 - 0.82) | 0.532 (0.378 - 0.686) | 0.518 (0.368 - 0.668) | 0.782 (0.598 - 0.966) | 0.64 (0.473 - 0.807) | 0.496 (0.35 - 0.642) | 0.721 (0.545 - 0.897) | 0.585 (0.427 - 0.744) | 0.572 (0.415 - 0.728) | 0.834 (0.645 - 1.023) | 0.681 (0.511 - 0.852) | 0.575 (0.419 - 0.731) | 0.845 (0.656 - 1.033) | 0.688 (0.518 - 0.858) | 0.557 (0.405 - 0.708) | 0.801 (0.619 - 0.983) | 0.653 (0.488 - 0.817) |
| **Cardiovascular diseases** | 247.501 (244.177 - 250.826) | 236.567 (233.316 - 239.817) | 268.727 (265.263 - 272.191) | 256.934 (253.584 - 260.284) | 248.562 (245.266 - 251.857) | 277.031 (273.553 - 280.509) | 249.328 (246.046 - 252.609) | 243.126 (239.885 - 246.367) | 268.095 (264.692 - 271.497) | 248.848 (245.591 - 252.104) | 242.234 (239.021 - 245.447) | 267.622 (264.245 - 270.999) | 235.07 (231.925 - 238.215) | 227.021 (223.93 - 230.111) | 252.666 (249.406 - 255.927) | 224.254 (221.211 - 227.298) | 215.313 (212.331 - 218.295) | 239.453 (236.309 - 242.598) |
| Artery diseases | 29.973 (28.816 - 31.13) | 37.557 (36.261 - 38.852) | 32.128 (30.93 - 33.326) | 28.864 (27.74 - 29.987) | 36.74 (35.472 - 38.008) | 31.262 (30.093 - 32.432) | 28.177 (27.074 - 29.28) | 35.413 (34.177 - 36.65) | 30.71 (29.558 - 31.861) | 27.825 (26.735 - 28.914) | 35.002 (33.781 - 36.224) | 30.248 (29.112 - 31.384) | 24.596 (23.578 - 25.613) | 31.008 (29.866 - 32.15) | 27.048 (25.982 - 28.115) | 21.935 (20.984 - 22.887) | 27.918 (26.845 - 28.992) | 24.36 (23.357 - 25.363) |
| Atrial fibrillation | 24.766 (23.716 - 25.816) | 28.075 (26.957 - 29.193) | 26.656 (25.567 - 27.746) | 26.453 (25.38 - 27.526) | 30.104 (28.959 - 31.249) | 28.726 (27.608 - 29.844) | 25.591 (24.54 - 26.642) | 30.487 (29.339 - 31.634) | 28.454 (27.346 - 29.562) | 26.737 (25.67 - 27.805) | 31.531 (30.371 - 32.69) | 29.538 (28.415 - 30.66) | 26.163 (25.112 - 27.214) | 30.605 (29.469 - 31.742) | 28.849 (27.746 - 29.953) | 25.973 (24.936 - 27.009) | 29.522 (28.417 - 30.627) | 28.263 (27.182 - 29.344) |
| Cerebrovascular disease | 38.5 (37.189 - 39.812) | 34.665 (33.42 - 35.91) | 39.645 (38.314 - 40.977) | 38.769 (37.466 - 40.072) | 35.078 (33.839 - 36.317) | 39.64 (38.323 - 40.957) | 37.512 (36.238 - 38.785) | 34.505 (33.284 - 35.727) | 38.635 (37.342 - 39.928) | 37.06 (35.804 - 38.317) | 34.237 (33.029 - 35.445) | 38.187 (36.911 - 39.463) | 34.86 (33.648 - 36.071) | 32.025 (30.864 - 33.186) | 35.78 (34.553 - 37.007) | 33.722 (32.543 - 34.902) | 30.904 (29.775 - 32.033) | 34.42 (33.228 - 35.612) |
| Chronic rheumatic heart diseases | 2.848 (2.491 - 3.205) | 2.234 (1.917 - 2.55) | 2.584 (2.244 - 2.924) | 3.196 (2.82 - 3.572) | 2.29 (1.972 - 2.608) | 2.773 (2.423 - 3.122) | 2.792 (2.445 - 3.14) | 2.065 (1.766 - 2.364) | 2.456 (2.13 - 2.782) | 2.971 (2.614 - 3.328) | 2.301 (1.988 - 2.615) | 2.679 (2.341 - 3.018) | 3.174 (2.807 - 3.54) | 2.33 (2.015 - 2.644) | 2.77 (2.427 - 3.113) | 2.97 (2.62 - 3.321) | 2.019 (1.73 - 2.307) | 2.482 (2.162 - 2.802) |
| Heart failure (specified) | 7.736 (7.147 - 8.324) | 8.893 (8.263 - 9.523) | 8.156 (7.552 - 8.759) | 7.569 (6.994 - 8.145) | 8.868 (8.245 - 9.492) | 8.216 (7.616 - 8.816) | 6.96 (6.411 - 7.508) | 8.379 (7.777 - 8.981) | 7.607 (7.034 - 8.18) | 7.121 (6.569 - 7.672) | 8.59 (7.984 - 9.196) | 7.904 (7.323 - 8.485) | 7.373 (6.816 - 7.93) | 8.955 (8.341 - 9.568) | 8.12 (7.536 - 8.705) | 7.22 (6.674 - 7.766) | 8.796 (8.194 - 9.399) | 8.065 (7.488 - 8.642) |
| Hypertensive diseases | 55.385 (53.813 - 56.957) | 59.24 (57.614 - 60.866) | 57.295 (55.696 - 58.894) | 61.056 (59.426 - 62.686) | 66.713 (65.008 - 68.418) | 63.724 (62.059 - 65.39) | 62.33 (60.691 - 63.97) | 67.518 (65.811 - 69.226) | 65.046 (63.371 - 66.721) | 61.163 (59.55 - 62.776) | 66.681 (64.997 - 68.366) | 63.957 (62.308 - 65.607) | 59.945 (58.356 - 61.534) | 64.896 (63.243 - 66.549) | 62.438 (60.817 - 64.06) | 58.162 (56.612 - 59.712) | 62.901 (61.29 - 64.513) | 60.611 (59.029 - 62.194) |
| Ischaemic heart disease | 89.231 (87.235 - 91.227) | 65.19 (63.483 - 66.896) | 84.127 (82.189 - 86.065) | 89.626 (87.646 - 91.607) | 66.436 (64.731 - 68.142) | 84.729 (82.804 - 86.655) | 83.716 (81.812 - 85.619) | 62.866 (61.217 - 64.515) | 79.15 (77.3 - 81.001) | 84.521 (82.623 - 86.419) | 62.754 (61.119 - 64.39) | 79.473 (77.632 - 81.314) | 77.939 (76.129 - 79.748) | 56.725 (55.181 - 58.268) | 72.498 (70.754 - 74.243) | 72.621 (70.89 - 74.351) | 51.876 (50.413 - 53.339) | 66.79 (65.13 - 68.45) |
| Non-rheumatic valve disorders | 8.934 (8.303 - 9.566) | 7.544 (6.964 - 8.125) | 8.614 (7.994 - 9.234) | 9.397 (8.756 - 10.038) | 7.992 (7.402 - 8.583) | 9.145 (8.514 - 9.777) | 9.067 (8.439 - 9.694) | 7.915 (7.329 - 8.501) | 8.913 (8.291 - 9.535) | 9.171 (8.544 - 9.798) | 8.033 (7.446 - 8.619) | 9.074 (8.45 - 9.697) | 8.983 (8.367 - 9.599) | 7.858 (7.282 - 8.434) | 8.818 (8.207 - 9.428) | 8.731 (8.129 - 9.333) | 7.308 (6.758 - 7.858) | 8.338 (7.75 - 8.926) |
| Other heart diseases | 46.573 (45.131 - 48.014) | 54.618 (53.057 - 56.179) | 49.5 (48.013 - 50.987) | 47.018 (45.586 - 48.45) | 56.223 (54.657 - 57.789) | 50.491 (49.007 - 51.975) | 44.852 (43.461 - 46.243) | 53.881 (52.356 - 55.406) | 48.17 (46.729 - 49.612) | 45.229 (43.842 - 46.617) | 54.263 (52.743 - 55.784) | 48.607 (47.168 - 50.046) | 43.052 (41.706 - 44.397) | 51.249 (49.78 - 52.718) | 46.196 (44.801 - 47.59) | 38.996 (37.726 - 40.265) | 46.724 (45.335 - 48.113) | 41.77 (40.456 - 43.083) |
| Phlebitis & thrombophlebitis | 1.514 (1.254 - 1.775) | 1.67 (1.397 - 1.943) | 1.638 (1.367 - 1.909) | 1.731 (1.456 - 2.007) | 1.928 (1.638 - 2.219) | 1.889 (1.601 - 2.177) | 1.576 (1.315 - 1.836) | 1.791 (1.514 - 2.069) | 1.758 (1.483 - 2.033) | 1.463 (1.213 - 1.713) | 1.709 (1.439 - 1.979) | 1.612 (1.35 - 1.874) | 1.321 (1.086 - 1.557) | 1.59 (1.332 - 1.848) | 1.521 (1.268 - 1.774) | 1.293 (1.063 - 1.523) | 1.512 (1.263 - 1.762) | 1.454 (1.209 - 1.698) |
| Pulmonary heart diseases | 10.014 (9.346 - 10.683) | 11.978 (11.247 - 12.709) | 10.675 (9.985 - 11.366) | 10.167 (9.5 - 10.834) | 12.68 (11.935 - 13.425) | 10.949 (10.257 - 11.641) | 9.667 (9.021 - 10.313) | 11.994 (11.274 - 12.714) | 10.543 (9.868 - 11.218) | 9.523 (8.885 - 10.161) | 11.84 (11.129 - 12.552) | 10.33 (9.665 - 10.994) | 9.133 (8.514 - 9.753) | 11.078 (10.395 - 11.761) | 9.842 (9.199 - 10.486) | 8.524 (7.931 - 9.116) | 10.683 (10.019 - 11.346) | 9.36 (8.739 - 9.981) |
| Residual - cardiovascular | 1.894 (1.603 - 2.185) | 2.396 (2.069 - 2.723) | 2.024 (1.724 - 2.324) | 2.11 (1.808 - 2.413) | 2.685 (2.344 - 3.027) | 2.254 (1.941 - 2.567) | 1.906 (1.619 - 2.192) | 2.444 (2.119 - 2.769) | 2.079 (1.78 - 2.379) | 1.993 (1.702 - 2.284) | 2.59 (2.259 - 2.922) | 2.193 (1.888 - 2.497) | 1.744 (1.474 - 2.015) | 2.269 (1.96 - 2.578) | 1.95 (1.663 - 2.236) | 1.982 (1.696 - 2.268) | 2.502 (2.181 - 2.823) | 2.128 (1.831 - 2.424) |
| Transient cerebral ischaemic attack | 0.735 (0.554 - 0.916) | 1.005 (0.793 - 1.216) | 0.867 (0.671 - 1.064) | 0.798 (0.613 - 0.984) | 1.078 (0.862 - 1.293) | 0.918 (0.719 - 1.117) | 0.766 (0.584 - 0.948) | 1.057 (0.843 - 1.271) | 0.892 (0.696 - 1.089) | 0.787 (0.604 - 0.97) | 1.095 (0.879 - 1.311) | 0.936 (0.737 - 1.136) | 0.684 (0.514 - 0.854) | 0.994 (0.789 - 1.199) | 0.837 (0.649 - 1.025) | 0.616 (0.457 - 0.775) | 0.896 (0.704 - 1.087) | 0.738 (0.564 - 0.913) |
| **Respiratory diseases** | 88.966 (86.971 - 90.961) | 96.224 (94.15 - 98.299) | 87.125 (85.151 - 89.099) | 118.75 (116.469 - 121.031) | 116.016 (113.762 - 118.271) | 117.658 (115.387 - 119.929) | 115.954 (113.717 - 118.192) | 111.581 (109.387 - 113.776) | 115.807 (113.571 - 118.044) | 113.104 (110.907 - 115.302) | 109.344 (107.183 - 111.506) | 111.832 (109.647 - 114.018) | 95.833 (93.825 - 97.842) | 97.777 (95.748 - 99.806) | 95.028 (93.027 - 97.028) | 86.949 (85.054 - 88.844) | 91.265 (89.323 - 93.206) | 86.124 (84.238 - 88.011) |
| Asthma | 0.834 (0.64 - 1.027) | 0.871 (0.673 - 1.068) | 0.893 (0.693 - 1.093) | 0.987 (0.78 - 1.194) | 1.103 (0.883 - 1.322) | 1.055 (0.841 - 1.269) | 1.011 (0.802 - 1.219) | 1.131 (0.91 - 1.351) | 1.077 (0.862 - 1.292) | 1.076 (0.862 - 1.291) | 1.122 (0.903 - 1.341) | 1.111 (0.894 - 1.329) | 1.015 (0.808 - 1.221) | 1.097 (0.883 - 1.312) | 1.059 (0.848 - 1.271) | 0.941 (0.744 - 1.138) | 1.06 (0.85 - 1.269) | 1.005 (0.801 - 1.209) |
| Bronchiectasis | 0.051 (0.004 - 0.098) | 0.054 (0.006 - 0.103) | 0.06 (0.009 - 0.111) | 0.055 (0.006 - 0.105) | 0.059 (0.008 - 0.111) | 0.058 (0.007 - 0.108) | 0.038 (-0.003 - 0.08) | 0.038 (-0.003 - 0.079) | 0.04 (-0.002 - 0.082) | 0.057 (0.008 - 0.106) | 0.053 (0.006 - 0.1) | 0.059 (0.009 - 0.109) | 0.027 (-0.007 - 0.062) | 0.028 (-0.007 - 0.064) | 0.029 (-0.007 - 0.065) | 0.038 (0 - 0.076) | 0.038 (0 - 0.077) | 0.041 (0.001 - 0.08) |
| COPD | 27.82 (26.703 - 28.936) | 25.595 (24.523 - 26.666) | 28.311 (27.184 - 29.437) | 27.493 (26.394 - 28.593) | 25.472 (24.414 - 26.53) | 27.898 (26.79 - 29.006) | 25.305 (24.258 - 26.351) | 24.317 (23.292 - 25.342) | 26.05 (24.988 - 27.111) | 26.638 (25.57 - 27.706) | 25.479 (24.434 - 26.524) | 27.304 (26.223 - 28.386) | 25.614 (24.576 - 26.652) | 23.979 (22.975 - 24.983) | 26.152 (25.103 - 27.201) | 24.823 (23.811 - 25.835) | 23.154 (22.176 - 24.131) | 25.407 (24.383 - 26.431) |
| COVID-19 | 0 (0 - 0) | 0 (0 - 0) | 0 (0 - 0) | 36.486 (35.221 - 37.751) | 21.001 (20.041 - 21.961) | 30.554 (29.397 - 31.712) | 44.057 (42.677 - 45.437) | 25.062 (24.022 - 26.103) | 36.711 (35.451 - 37.97) | 36.894 (35.641 - 38.146) | 21.458 (20.503 - 22.413) | 30.4 (29.263 - 31.537) | 16.149 (15.323 - 16.975) | 9.613 (8.976 - 10.25) | 13.288 (12.538 - 14.037) | 7.123 (6.581 - 7.665) | 4.151 (3.738 - 4.565) | 5.676 (5.192 - 6.16) |
| Influenza | 1.909 (1.617 - 2.202) | 1.188 (0.958 - 1.419) | 1.666 (1.393 - 1.939) | 1.818 (1.536 - 2.099) | 1.156 (0.932 - 1.38) | 1.602 (1.337 - 1.866) | 0.047 (0.003 - 0.09) | 0.033 (-0.004 - 0.07) | 0.036 (-0.003 - 0.074) | 1.974 (1.683 - 2.265) | 1.174 (0.95 - 1.397) | 1.68 (1.412 - 1.948) | 2.592 (2.261 - 2.924) | 1.535 (1.28 - 1.79) | 2.169 (1.866 - 2.472) | 3.61 (3.224 - 3.996) | 2.114 (1.819 - 2.409) | 2.941 (2.593 - 3.29) |
| Other ALRI | 0.676 (0.503 - 0.849) | 0.766 (0.583 - 0.95) | 0.712 (0.535 - 0.889) | 0.469 (0.327 - 0.612) | 0.542 (0.389 - 0.695) | 0.496 (0.349 - 0.643) | 0.29 (0.179 - 0.401) | 0.348 (0.227 - 0.47) | 0.32 (0.204 - 0.437) | 0.414 (0.282 - 0.547) | 0.48 (0.337 - 0.623) | 0.441 (0.304 - 0.578) | 0.467 (0.327 - 0.607) | 0.537 (0.387 - 0.687) | 0.491 (0.347 - 0.635) | 0.423 (0.291 - 0.554) | 0.499 (0.355 - 0.642) | 0.448 (0.312 - 0.584) |
| Other diseases of pleura | 2.381 (2.056 - 2.706) | 3.353 (2.967 - 3.739) | 2.721 (2.373 - 3.069) | 2.162 (1.854 - 2.47) | 3.091 (2.723 - 3.459) | 2.478 (2.148 - 2.808) | 2.321 (2.004 - 2.637) | 3.313 (2.935 - 3.692) | 2.671 (2.331 - 3.011) | 2.344 (2.027 - 2.661) | 3.338 (2.96 - 3.716) | 2.718 (2.377 - 3.06) | 2.425 (2.105 - 2.746) | 3.426 (3.045 - 3.807) | 2.81 (2.465 - 3.155) | 2.239 (1.935 - 2.544) | 3.204 (2.84 - 3.569) | 2.616 (2.287 - 2.945) |
| Other interstitial respiratory diseases | 6.717 (6.169 - 7.264) | 8.481 (7.866 - 9.096) | 7.22 (6.653 - 7.788) | 6.525 (5.991 - 7.06) | 8.531 (7.919 - 9.142) | 7.075 (6.518 - 7.631) | 6.69 (6.153 - 7.228) | 8.583 (7.974 - 9.192) | 7.252 (6.692 - 7.812) | 5.907 (5.405 - 6.41) | 7.37 (6.81 - 7.931) | 6.357 (5.837 - 6.878) | 5.356 (4.881 - 5.831) | 6.609 (6.081 - 7.136) | 5.751 (5.259 - 6.244) | 5.065 (4.607 - 5.522) | 6.263 (5.754 - 6.772) | 5.448 (4.973 - 5.922) |
| Pneumonia | 31.07 (29.892 - 32.249) | 37.563 (36.268 - 38.858) | 32.027 (30.831 - 33.223) | 33.872 (32.654 - 35.09) | 42.518 (41.153 - 43.883) | 35.292 (34.049 - 36.535) | 31.722 (30.552 - 32.893) | 40.355 (39.034 - 41.675) | 33.123 (31.926 - 34.319) | 28.747 (27.641 - 29.854) | 36.074 (34.834 - 37.314) | 30.303 (29.166 - 31.44) | 28.971 (27.866 - 30.075) | 35.476 (34.253 - 36.698) | 30.639 (29.504 - 31.775) | 27.85 (26.777 - 28.924) | 34.304 (33.112 - 35.495) | 29.513 (28.408 - 30.618) |
| Pneumonitis | 5.749 (5.242 - 6.257) | 6.973 (6.415 - 7.532) | 5.787 (5.277 - 6.296) | 5.925 (5.416 - 6.435) | 7.3 (6.734 - 7.865) | 6.013 (5.5 - 6.526) | 6.252 (5.734 - 6.77) | 7.62 (7.048 - 8.193) | 6.189 (5.674 - 6.705) | 6.094 (5.584 - 6.604) | 7.489 (6.924 - 8.055) | 6.252 (5.735 - 6.769) | 6.266 (5.752 - 6.779) | 7.518 (6.956 - 8.08) | 6.34 (5.823 - 6.856) | 6.774 (6.245 - 7.303) | 8.078 (7.5 - 8.656) | 6.962 (6.426 - 7.498) |
| Residual - respiratory | 2.563 (2.226 - 2.901) | 2.96 (2.597 - 3.323) | 2.673 (2.328 - 3.018) | 2.49 (2.16 - 2.821) | 3.067 (2.701 - 3.433) | 2.672 (2.33 - 3.014) | 2.056 (1.758 - 2.354) | 2.625 (2.289 - 2.962) | 2.312 (1.996 - 2.628) | 2.221 (1.912 - 2.529) | 2.802 (2.456 - 3.148) | 2.445 (2.122 - 2.769) | 2.876 (2.528 - 3.224) | 3.462 (3.08 - 3.844) | 3.067 (2.708 - 3.427) | 2.558 (2.233 - 2.883) | 3.192 (2.829 - 3.555) | 2.809 (2.469 - 3.15) |
| **Digestive diseases** | 33.002 (31.79 - 34.215) | 34.581 (33.34 - 35.823) | 32.929 (31.718 - 34.141) | 33.194 (31.989 - 34.399) | 34.939 (33.703 - 36.176) | 33.462 (32.252 - 34.671) | 32.797 (31.607 - 33.987) | 34.723 (33.499 - 35.948) | 32.944 (31.752 - 34.137) | 32.977 (31.789 - 34.164) | 35.011 (33.788 - 36.235) | 33.145 (31.955 - 34.335) | 31.484 (30.333 - 32.634) | 33.048 (31.87 - 34.227) | 31.862 (30.705 - 33.02) | 31.368 (30.23 - 32.507) | 32.921 (31.755 - 34.087) | 31.534 (30.393 - 32.676) |
| Cirrhosis of the liver | 6.637 (6.093 - 7.182) | 6.411 (5.876 - 6.946) | 6.854 (6.301 - 7.407) | 6.51 (5.976 - 7.044) | 6.207 (5.686 - 6.729) | 6.665 (6.125 - 7.206) | 6.284 (5.763 - 6.805) | 6.055 (5.543 - 6.566) | 6.448 (5.921 - 6.976) | 6.51 (5.982 - 7.038) | 6.142 (5.63 - 6.655) | 6.546 (6.017 - 7.076) | 5.979 (5.478 - 6.48) | 5.515 (5.034 - 5.996) | 6.07 (5.565 - 6.575) | 5.733 (5.247 - 6.219) | 5.29 (4.823 - 5.756) | 5.733 (5.247 - 6.219) |
| Diseases of oesophagus, stomach & duodenum | 3.229 (2.848 - 3.609) | 3.593 (3.192 - 3.994) | 3.443 (3.05 - 3.836) | 3.413 (3.027 - 3.799) | 3.697 (3.294 - 4.099) | 3.664 (3.264 - 4.064) | 3.438 (3.053 - 3.822) | 3.846 (3.439 - 4.253) | 3.742 (3.341 - 4.143) | 3.144 (2.778 - 3.51) | 3.636 (3.243 - 4.03) | 3.485 (3.099 - 3.87) | 3.103 (2.742 - 3.464) | 3.488 (3.106 - 3.871) | 3.393 (3.015 - 3.77) | 3.197 (2.833 - 3.561) | 3.494 (3.113 - 3.874) | 3.439 (3.061 - 3.816) |
| Diseases of peritoneum | 1.184 (0.955 - 1.414) | 1.558 (1.295 - 1.821) | 1.299 (1.058 - 1.539) | 1.304 (1.064 - 1.543) | 1.681 (1.409 - 1.953) | 1.414 (1.165 - 1.664) | 1.225 (0.995 - 1.456) | 1.612 (1.348 - 1.876) | 1.353 (1.111 - 1.596) | 1.176 (0.952 - 1.4) | 1.542 (1.285 - 1.798) | 1.288 (1.054 - 1.523) | 1.28 (1.047 - 1.512) | 1.653 (1.389 - 1.917) | 1.411 (1.167 - 1.654) | 1.064 (0.854 - 1.274) | 1.381 (1.143 - 1.62) | 1.141 (0.924 - 1.358) |
| Disorders of gallbladder, biliary tract & pancreas | 3.574 (3.175 - 3.973) | 3.437 (3.046 - 3.829) | 3.656 (3.253 - 4.06) | 3.309 (2.929 - 3.688) | 3.244 (2.868 - 3.62) | 3.389 (3.005 - 3.774) | 3.607 (3.213 - 4.002) | 3.48 (3.093 - 3.868) | 3.627 (3.231 - 4.022) | 3.443 (3.06 - 3.826) | 3.375 (2.996 - 3.755) | 3.518 (3.13 - 3.905) | 3.329 (2.953 - 3.704) | 3.214 (2.846 - 3.582) | 3.42 (3.04 - 3.8) | 3.285 (2.916 - 3.655) | 3.174 (2.812 - 3.537) | 3.334 (2.962 - 3.706) |
| Other diseases of intestines | 6.528 (5.989 - 7.067) | 6.536 (5.996 - 7.075) | 6.653 (6.109 - 7.197) | 6.483 (5.951 - 7.015) | 6.497 (5.965 - 7.03) | 6.669 (6.129 - 7.209) | 6.126 (5.611 - 6.641) | 6.154 (5.639 - 6.67) | 6.276 (5.755 - 6.797) | 6.319 (5.799 - 6.838) | 6.454 (5.929 - 6.98) | 6.491 (5.964 - 7.018) | 6.261 (5.747 - 6.775) | 6.254 (5.74 - 6.768) | 6.431 (5.91 - 6.952) | 6.14 (5.636 - 6.643) | 6.236 (5.729 - 6.744) | 6.364 (5.852 - 6.877) |
| Other diseases of liver | 5.367 (4.879 - 5.855) | 6.362 (5.831 - 6.894) | 5.388 (4.899 - 5.877) | 4.997 (4.53 - 5.465) | 6.153 (5.634 - 6.672) | 5.208 (4.73 - 5.685) | 5.185 (4.713 - 5.658) | 6.374 (5.85 - 6.898) | 5.423 (4.94 - 5.906) | 5.123 (4.655 - 5.591) | 6.352 (5.831 - 6.873) | 5.362 (4.883 - 5.84) | 4.732 (4.288 - 5.177) | 5.911 (5.413 - 6.408) | 4.985 (4.528 - 5.441) | 4.738 (4.295 - 5.18) | 5.884 (5.391 - 6.377) | 5.024 (4.569 - 5.48) |
| Residual - digestive | 4.737 (4.279 - 5.195) | 5.412 (4.922 - 5.902) | 5.032 (4.56 - 5.505) | 5.475 (4.986 - 5.965) | 6.128 (5.61 - 6.646) | 5.787 (5.284 - 6.29) | 4.925 (4.465 - 5.385) | 5.605 (5.114 - 6.096) | 5.262 (4.786 - 5.738) | 5.515 (5.03 - 6.001) | 6.214 (5.699 - 6.73) | 5.852 (5.352 - 6.353) | 5.419 (4.941 - 5.897) | 6.017 (5.513 - 6.52) | 5.717 (5.226 - 6.207) | 5.489 (5.012 - 5.965) | 6.138 (5.635 - 6.642) | 5.841 (5.35 - 6.333) |
| **Skin diseases** | 3.922 (3.503 - 4.342) | 4.894 (4.426 - 5.363) | 4.02 (3.595 - 4.445) | 3.867 (3.456 - 4.277) | 4.961 (4.496 - 5.426) | 3.989 (3.572 - 4.407) | 3.749 (3.347 - 4.151) | 4.771 (4.318 - 5.225) | 3.92 (3.509 - 4.331) | 3.708 (3.311 - 4.105) | 4.844 (4.39 - 5.298) | 3.987 (3.575 - 4.399) | 4.032 (3.62 - 4.444) | 5.017 (4.558 - 5.477) | 4.19 (3.77 - 4.61) | 3.807 (3.411 - 4.204) | 4.82 (4.374 - 5.266) | 3.961 (3.557 - 4.365) |
| Infections of skin | 0.335 (0.214 - 0.457) | 0.401 (0.267 - 0.534) | 0.367 (0.24 - 0.495) | 0.294 (0.18 - 0.407) | 0.395 (0.264 - 0.527) | 0.329 (0.208 - 0.449) | 0.32 (0.203 - 0.438) | 0.395 (0.265 - 0.526) | 0.365 (0.239 - 0.49) | 0.242 (0.141 - 0.343) | 0.313 (0.198 - 0.428) | 0.285 (0.175 - 0.395) | 0.299 (0.186 - 0.411) | 0.34 (0.221 - 0.46) | 0.328 (0.211 - 0.446) | 0.307 (0.194 - 0.419) | 0.367 (0.244 - 0.49) | 0.34 (0.222 - 0.459) |
| Residual - skin diseases | 2.733 (2.382 - 3.084) | 3.587 (3.186 - 3.988) | 3.061 (2.69 - 3.432) | 2.742 (2.397 - 3.088) | 3.697 (3.296 - 4.099) | 3.102 (2.734 - 3.469) | 2.652 (2.314 - 2.991) | 3.541 (3.15 - 3.931) | 3.004 (2.644 - 3.364) | 2.652 (2.316 - 2.987) | 3.636 (3.243 - 4.029) | 3.107 (2.744 - 3.471) | 2.925 (2.573 - 3.276) | 3.801 (3.401 - 4.201) | 3.283 (2.911 - 3.654) | 2.717 (2.383 - 3.052) | 3.61 (3.224 - 3.995) | 3.067 (2.711 - 3.422) |
| **Musculoskeletal conditions** | 8.96 (8.326 - 9.593) | 10.725 (10.032 - 11.419) | 9.273 (8.629 - 9.917) | 9.252 (8.618 - 9.886) | 11.349 (10.647 - 12.052) | 9.714 (9.064 - 10.365) | 9.306 (8.672 - 9.939) | 11.38 (10.68 - 12.08) | 9.789 (9.14 - 10.439) | 9.534 (8.897 - 10.171) | 11.827 (11.117 - 12.537) | 10.065 (9.41 - 10.72) | 9.807 (9.164 - 10.449) | 11.731 (11.028 - 12.434) | 10.244 (9.587 - 10.9) | 9.528 (8.9 - 10.155) | 11.601 (10.908 - 12.293) | 10.04 (9.396 - 10.685) |
| Infectious arthropathies | 0.106 (0.039 - 0.174) | 0.089 (0.027 - 0.151) | 0.104 (0.037 - 0.171) | 0.097 (0.032 - 0.162) | 0.087 (0.026 - 0.148) | 0.098 (0.033 - 0.163) | 0.16 (0.076 - 0.244) | 0.13 (0.054 - 0.206) | 0.159 (0.075 - 0.244) | 0.159 (0.077 - 0.24) | 0.124 (0.052 - 0.197) | 0.152 (0.072 - 0.233) | 0.25 (0.147 - 0.353) | 0.174 (0.088 - 0.259) | 0.216 (0.121 - 0.311) | 0.198 (0.108 - 0.289) | 0.14 (0.064 - 0.216) | 0.172 (0.088 - 0.257) |
| Osteoarthritis | 0.578 (0.418 - 0.738) | 0.765 (0.581 - 0.95) | 0.67 (0.498 - 0.843) | 0.547 (0.392 - 0.701) | 0.733 (0.555 - 0.911) | 0.63 (0.465 - 0.796) | 0.542 (0.389 - 0.695) | 0.744 (0.565 - 0.923) | 0.643 (0.477 - 0.81) | 0.588 (0.43 - 0.747) | 0.768 (0.587 - 0.949) | 0.655 (0.488 - 0.822) | 0.5 (0.356 - 0.645) | 0.67 (0.503 - 0.838) | 0.58 (0.424 - 0.736) | 0.436 (0.302 - 0.571) | 0.593 (0.436 - 0.749) | 0.501 (0.357 - 0.645) |
| Osteopathies & chondropathies | 2.885 (2.527 - 3.244) | 3.735 (3.326 - 4.143) | 3.25 (2.869 - 3.63) | 2.82 (2.471 - 3.169) | 3.705 (3.305 - 4.105) | 3.242 (2.868 - 3.617) | 2.628 (2.292 - 2.964) | 3.545 (3.155 - 3.935) | 3.071 (2.708 - 3.434) | 2.666 (2.33 - 3.003) | 3.597 (3.206 - 3.988) | 3.127 (2.762 - 3.492) | 2.654 (2.319 - 2.988) | 3.45 (3.068 - 3.831) | 3.05 (2.692 - 3.409) | 2.524 (2.201 - 2.848) | 3.383 (3.008 - 3.758) | 2.944 (2.594 - 3.293) |
| Residual - musculoskeletal | 2.762 (2.41 - 3.115) | 3.46 (3.066 - 3.854) | 3.17 (2.793 - 3.547) | 2.911 (2.555 - 3.268) | 3.783 (3.377 - 4.19) | 3.377 (2.993 - 3.761) | 2.937 (2.581 - 3.293) | 3.798 (3.393 - 4.203) | 3.364 (2.983 - 3.746) | 3.19 (2.821 - 3.558) | 4.22 (3.796 - 4.645) | 3.712 (3.314 - 4.11) | 3.465 (3.083 - 3.847) | 4.372 (3.943 - 4.801) | 3.936 (3.529 - 4.343) | 3.54 (3.158 - 3.921) | 4.51 (4.079 - 4.941) | 4.024 (3.617 - 4.431) |
| Rheumatoid arthritis | 0.499 (0.349 - 0.65) | 0.526 (0.371 - 0.68) | 0.488 (0.34 - 0.637) | 0.601 (0.439 - 0.763) | 0.69 (0.516 - 0.863) | 0.658 (0.488 - 0.828) | 0.661 (0.492 - 0.83) | 0.733 (0.555 - 0.911) | 0.698 (0.525 - 0.872) | 0.551 (0.398 - 0.703) | 0.671 (0.503 - 0.839) | 0.615 (0.454 - 0.776) | 0.605 (0.446 - 0.764) | 0.668 (0.5 - 0.835) | 0.63 (0.467 - 0.793) | 0.643 (0.479 - 0.807) | 0.712 (0.54 - 0.885) | 0.688 (0.519 - 0.858) |
| Systemic connective tissue disorders | 0.596 (0.432 - 0.76) | 0.56 (0.401 - 0.719) | 0.581 (0.419 - 0.743) | 0.561 (0.404 - 0.718) | 0.558 (0.401 - 0.715) | 0.567 (0.409 - 0.725) | 0.729 (0.552 - 0.906) | 0.66 (0.491 - 0.828) | 0.706 (0.531 - 0.88) | 0.638 (0.473 - 0.803) | 0.612 (0.45 - 0.773) | 0.63 (0.466 - 0.794) | 0.681 (0.512 - 0.849) | 0.617 (0.456 - 0.778) | 0.669 (0.502 - 0.837) | 0.617 (0.457 - 0.778) | 0.565 (0.412 - 0.718) | 0.609 (0.45 - 0.768) |
| **Genitourinary diseases** | 49.654 (48.166 - 51.142) | 57.866 (56.26 - 59.472) | 48.776 (47.301 - 50.25) | 55.338 (53.784 - 56.892) | 64.365 (62.689 - 66.041) | 54.979 (53.43 - 56.528) | 54.032 (52.504 - 55.56) | 64.14 (62.475 - 65.806) | 53.721 (52.197 - 55.245) | 53.881 (52.366 - 55.396) | 64.698 (63.038 - 66.358) | 54.082 (52.564 - 55.599) | 51.685 (50.209 - 53.162) | 62.45 (60.828 - 64.073) | 51.94 (50.46 - 53.42) | 51.026 (49.573 - 52.479) | 61.118 (59.528 - 62.709) | 51.319 (49.862 - 52.777) |
| Glomerular diseases | 0.212 (0.115 - 0.309) | 0.196 (0.102 - 0.289) | 0.213 (0.116 - 0.31) | 0.138 (0.061 - 0.216) | 0.138 (0.06 - 0.217) | 0.144 (0.064 - 0.224) | 0.203 (0.109 - 0.297) | 0.209 (0.114 - 0.304) | 0.205 (0.111 - 0.3) | 0.142 (0.063 - 0.22) | 0.157 (0.075 - 0.239) | 0.149 (0.069 - 0.229) | 0.187 (0.097 - 0.276) | 0.18 (0.092 - 0.268) | 0.187 (0.097 - 0.277) | 0.195 (0.105 - 0.284) | 0.176 (0.092 - 0.26) | 0.193 (0.104 - 0.281) |
| Hyperplasia of prostate | 0.451 (0.309 - 0.592) | 0.543 (0.389 - 0.698) | 0.488 (0.341 - 0.634) | 0.572 (0.415 - 0.729) | 0.727 (0.549 - 0.905) | 0.649 (0.482 - 0.817) | 0.61 (0.447 - 0.773) | 0.753 (0.572 - 0.934) | 0.664 (0.494 - 0.834) | 0.61 (0.449 - 0.772) | 0.733 (0.557 - 0.91) | 0.681 (0.511 - 0.852) | 0.559 (0.404 - 0.714) | 0.667 (0.498 - 0.835) | 0.61 (0.449 - 0.771) | 0.538 (0.388 - 0.687) | 0.666 (0.5 - 0.832) | 0.593 (0.436 - 0.75) |
| Renal failure | 31.765 (30.575 - 32.955) | 39.579 (38.25 - 40.908) | 34.706 (33.462 - 35.95) | 35.034 (33.797 - 36.27) | 44.008 (42.622 - 45.394) | 38.722 (37.422 - 40.022) | 34.005 (32.793 - 35.217) | 43.285 (41.917 - 44.653) | 37.572 (36.298 - 38.847) | 33.766 (32.567 - 34.966) | 43.869 (42.502 - 45.236) | 37.865 (36.595 - 39.135) | 32.123 (30.959 - 33.287) | 42.048 (40.716 - 43.379) | 36.164 (34.93 - 37.399) | 31.812 (30.665 - 32.959) | 41.184 (39.879 - 42.489) | 35.595 (34.382 - 36.809) |
| Renal tubulo-interstitial diseases | 0.858 (0.662 - 1.055) | 0.946 (0.74 - 1.153) | 0.895 (0.694 - 1.096) | 0.981 (0.774 - 1.188) | 1.089 (0.871 - 1.307) | 1.048 (0.835 - 1.262) | 1.015 (0.806 - 1.225) | 1.108 (0.889 - 1.327) | 1.055 (0.841 - 1.269) | 0.936 (0.736 - 1.137) | 1.056 (0.843 - 1.269) | 1.013 (0.805 - 1.221) | 0.954 (0.754 - 1.154) | 1.043 (0.834 - 1.252) | 0.985 (0.782 - 1.188) | 0.958 (0.758 - 1.157) | 1.038 (0.831 - 1.246) | 1.007 (0.802 - 1.211) |
| Residual - genitourinary | 5.802 (5.294 - 6.31) | 6.496 (5.959 - 7.034) | 6.228 (5.702 - 6.754) | 7.322 (6.757 - 7.887) | 7.53 (6.957 - 8.103) | 7.686 (7.107 - 8.265) | 6.823 (6.281 - 7.365) | 7.807 (7.227 - 8.388) | 7.459 (6.892 - 8.026) | 7.018 (6.472 - 7.565) | 7.911 (7.331 - 8.49) | 7.647 (7.077 - 8.217) | 6.703 (6.171 - 7.235) | 7.832 (7.257 - 8.407) | 7.457 (6.896 - 8.018) | 7.02 (6.481 - 7.559) | 7.986 (7.411 - 8.56) | 7.794 (7.226 - 8.362) |
| Urolithiasis | 0.209 (0.113 - 0.306) | 0.188 (0.096 - 0.279) | 0.196 (0.103 - 0.29) | 0.234 (0.132 - 0.336) | 0.242 (0.139 - 0.345) | 0.242 (0.139 - 0.346) | 0.246 (0.143 - 0.35) | 0.253 (0.147 - 0.358) | 0.247 (0.143 - 0.352) | 0.244 (0.141 - 0.347) | 0.239 (0.137 - 0.34) | 0.242 (0.14 - 0.345) | 0.284 (0.174 - 0.393) | 0.24 (0.139 - 0.341) | 0.257 (0.153 - 0.362) | 0.219 (0.124 - 0.313) | 0.212 (0.119 - 0.305) | 0.219 (0.124 - 0.314) |
| **Maternal conditions** | 0.096 (0.032 - 0.16) | 0.103 (0.037 - 0.169) | 0.09 (0.028 - 0.152) | 0.05 (0.004 - 0.096) | 0.048 (0.003 - 0.094) | 0.039 (-0.002 - 0.08) | 0.045 (0.001 - 0.089) | 0.045 (0.001 - 0.09) | 0.041 (-0.001 - 0.083) | 0.038 (-0.002 - 0.078) | 0.03 (-0.006 - 0.065) | 0.027 (-0.007 - 0.061) | 0.033 (-0.005 - 0.07) | 0.035 (-0.004 - 0.073) | 0.031 (-0.005 - 0.068) | 0.095 (0.029 - 0.161) | 0.107 (0.037 - 0.177) | 0.083 (0.022 - 0.145) |
| Maternal conditions | 0.092 (0.029 - 0.154) | 0.097 (0.033 - 0.161) | 0.085 (0.025 - 0.145) | 0.049 (0.003 - 0.094) | 0.047 (0.002 - 0.092) | 0.038 (-0.002 - 0.078) | 0.044 (0 - 0.087) | 0.044 (0 - 0.087) | 0.04 (-0.002 - 0.081) | 0.037 (-0.003 - 0.077) | 0.028 (-0.007 - 0.063) | 0.025 (-0.008 - 0.059) | 0.033 (-0.005 - 0.07) | 0.035 (-0.004 - 0.073) | 0.031 (-0.005 - 0.068) | 0.094 (0.028 - 0.159) | 0.103 (0.034 - 0.171) | 0.079 (0.019 - 0.139) |
| **Perinatal conditions** | 1.156 (0.936 - 1.376) | 1.145 (0.925 - 1.364) | 1.348 (1.11 - 1.586) | 1.222 (0.994 - 1.45) | 1.224 (0.996 - 1.452) | 1.414 (1.169 - 1.659) | 1.096 (0.879 - 1.314) | 1.1 (0.883 - 1.318) | 1.298 (1.061 - 1.534) | 0.892 (0.699 - 1.085) | 0.888 (0.695 - 1.081) | 1.015 (0.809 - 1.222) | 1.013 (0.803 - 1.222) | 1.025 (0.814 - 1.237) | 1.203 (0.974 - 1.432) | 1.186 (0.952 - 1.421) | 1.203 (0.967 - 1.439) | 1.354 (1.103 - 1.605) |
| Perinatal conditions | 1.141 (0.922 - 1.36) | 1.132 (0.914 - 1.35) | 1.339 (1.102 - 1.577) | 1.208 (0.981 - 1.435) | 1.207 (0.98 - 1.434) | 1.402 (1.158 - 1.646) | 1.088 (0.871 - 1.305) | 1.093 (0.876 - 1.31) | 1.294 (1.057 - 1.53) | 0.88 (0.688 - 1.071) | 0.877 (0.686 - 1.069) | 1.009 (0.803 - 1.214) | 1.005 (0.796 - 1.214) | 1.016 (0.806 - 1.226) | 1.196 (0.968 - 1.424) | 1.181 (0.947 - 1.416) | 1.196 (0.96 - 1.431) | 1.348 (1.098 - 1.598) |
| **Congenital conditions** | 1.883 (1.596 - 2.169) | 1.645 (1.377 - 1.912) | 1.978 (1.685 - 2.271) | 1.972 (1.68 - 2.263) | 1.784 (1.507 - 2.061) | 2.089 (1.789 - 2.389) | 1.844 (1.562 - 2.127) | 1.629 (1.364 - 1.895) | 1.915 (1.627 - 2.203) | 1.921 (1.635 - 2.206) | 1.666 (1.4 - 1.932) | 2.003 (1.711 - 2.295) | 1.875 (1.593 - 2.156) | 1.629 (1.366 - 1.892) | 1.943 (1.656 - 2.23) | 2.11 (1.809 - 2.412) | 1.915 (1.627 - 2.203) | 2.233 (1.922 - 2.544) |
| Congenital conditions | 1.791 (1.512 - 2.07) | 1.487 (1.233 - 1.742) | 1.844 (1.561 - 2.127) | 1.847 (1.565 - 2.129) | 1.597 (1.335 - 1.859) | 1.934 (1.645 - 2.222) | 1.747 (1.472 - 2.022) | 1.459 (1.208 - 1.711) | 1.765 (1.489 - 2.042) | 1.82 (1.542 - 2.098) | 1.479 (1.228 - 1.73) | 1.835 (1.556 - 2.114) | 1.777 (1.503 - 2.051) | 1.45 (1.202 - 1.698) | 1.786 (1.511 - 2.061) | 1.971 (1.679 - 2.263) | 1.701 (1.429 - 1.972) | 2.055 (1.756 - 2.354) |
| **Ill-defined causes** | 27.008 (25.91 - 28.105) | 25.391 (24.327 - 26.455) | 34.327 (33.09 - 35.565) | 29.75 (28.612 - 30.889) | 27.777 (26.677 - 28.878) | 37.604 (36.324 - 38.885) | 32.289 (31.11 - 33.468) | 30.157 (29.017 - 31.297) | 40.81 (39.485 - 42.136) | 30.809 (29.664 - 31.954) | 29.056 (27.944 - 30.168) | 39.245 (37.952 - 40.537) | 28.804 (27.701 - 29.907) | 27.174 (26.103 - 28.246) | 36.698 (35.453 - 37.943) | 29.143 (28.044 - 30.243) | 27.244 (26.181 - 28.306) | 36.855 (35.619 - 38.091) |
| Ill-defined causes | 27.008 (25.91 - 28.105) | 25.143 (24.084 - 26.202) | 34.041 (32.809 - 35.273) | 29.75 (28.612 - 30.889) | 27.523 (26.428 - 28.619) | 37.304 (36.029 - 38.58) | 32.289 (31.11 - 33.468) | 29.823 (28.689 - 30.956) | 40.414 (39.095 - 41.734) | 30.809 (29.664 - 31.954) | 28.776 (27.669 - 29.883) | 38.916 (37.629 - 40.203) | 28.804 (27.701 - 29.907) | 26.957 (25.89 - 28.024) | 36.445 (35.204 - 37.685) | 29.143 (28.044 - 30.243) | 26.962 (25.905 - 28.019) | 36.527 (35.296 - 37.757) |
| **Injuries** | 9.113 (8.479 - 9.748) | 11.431 (10.72 - 12.143) | 8.543 (7.928 - 9.159) | 7.934 (7.346 - 8.523) | 10.147 (9.482 - 10.812) | 7.654 (7.077 - 8.232) | 9.2 (8.571 - 9.829) | 11.764 (11.052 - 12.475) | 8.876 (8.258 - 9.495) | 9.261 (8.632 - 9.889) | 11.867 (11.156 - 12.578) | 8.968 (8.35 - 9.587) | 9.33 (8.704 - 9.957) | 11.907 (11.2 - 12.615) | 8.981 (8.366 - 9.595) | 9.973 (9.328 - 10.618) | 12.756 (12.027 - 13.484) | 9.63 (8.997 - 10.263) |
| Burns | 0.028 (-0.007 - 0.063) | 0.037 (-0.003 - 0.078) | 0.029 (-0.007 - 0.065) | 0.036 (-0.004 - 0.075) | 0.045 (0.001 - 0.09) | 0.034 (-0.004 - 0.073) | 0.062 (0.011 - 0.114) | 0.072 (0.017 - 0.127) | 0.052 (0.005 - 0.099) | 0.025 (-0.007 - 0.057) | 0.033 (-0.004 - 0.071) | 0.026 (-0.007 - 0.059) | 0.07 (0.015 - 0.125) | 0.085 (0.024 - 0.145) | 0.062 (0.01 - 0.114) | 0.047 (0.003 - 0.091) | 0.055 (0.007 - 0.103) | 0.04 (-0.001 - 0.081) |
| Dislocations | 0.039 (-0.001 - 0.08) | 0.054 (0.006 - 0.102) | 0.043 (0 - 0.085) | 0.042 (-0.001 - 0.085) | 0.059 (0.008 - 0.109) | 0.046 (0.001 - 0.091) | 0.046 (0.002 - 0.091) | 0.071 (0.016 - 0.127) | 0.059 (0.008 - 0.109) | 0.051 (0.004 - 0.098) | 0.063 (0.011 - 0.116) | 0.048 (0.002 - 0.094) | 0.068 (0.015 - 0.121) | 0.097 (0.034 - 0.161) | 0.077 (0.021 - 0.134) | 0.04 (-0.001 - 0.082) | 0.057 (0.008 - 0.107) | 0.046 (0.002 - 0.09) |
| Drowning/submersion injuries | 0.011 (-0.011 - 0.033) | 0.011 (-0.011 - 0.033) | 0.007 (-0.01 - 0.025) | 0.02 (-0.009 - 0.048) | 0.022 (-0.009 - 0.053) | 0.016 (-0.01 - 0.042) | 0.021 (-0.009 - 0.051) | 0.025 (-0.008 - 0.058) | 0.019 (-0.01 - 0.047) | 0.021 (-0.008 - 0.051) | 0.03 (-0.005 - 0.064) | 0.023 (-0.008 - 0.054) | 0.015 (-0.011 - 0.041) | 0.017 (-0.01 - 0.044) | 0.012 (-0.011 - 0.035) | 0.028 (-0.006 - 0.061) | 0.032 (-0.004 - 0.068) | 0.023 (-0.008 - 0.054) |
| Hip fracture | 1.198 (0.968 - 1.428) | 1.714 (1.439 - 1.99) | 1.365 (1.119 - 1.611) | 0.908 (0.71 - 1.106) | 1.341 (1.099 - 1.582) | 1.084 (0.867 - 1.301) | 1.221 (0.992 - 1.45) | 1.752 (1.478 - 2.027) | 1.401 (1.156 - 1.647) | 1.272 (1.039 - 1.505) | 1.81 (1.532 - 2.088) | 1.444 (1.196 - 1.692) | 1.145 (0.926 - 1.365) | 1.67 (1.405 - 1.935) | 1.343 (1.106 - 1.581) | 1.276 (1.045 - 1.507) | 1.845 (1.567 - 2.122) | 1.48 (1.231 - 1.729) |
| Humerus fracture | 0.192 (0.1 - 0.284) | 0.279 (0.169 - 0.39) | 0.224 (0.125 - 0.323) | 0.206 (0.113 - 0.3) | 0.307 (0.193 - 0.421) | 0.249 (0.146 - 0.352) | 0.221 (0.124 - 0.318) | 0.317 (0.201 - 0.433) | 0.253 (0.149 - 0.357) | 0.189 (0.1 - 0.278) | 0.283 (0.174 - 0.391) | 0.231 (0.133 - 0.329) | 0.21 (0.115 - 0.304) | 0.292 (0.181 - 0.403) | 0.232 (0.133 - 0.331) | 0.182 (0.095 - 0.269) | 0.262 (0.158 - 0.367) | 0.21 (0.116 - 0.304) |
| Internal & crush injuries | 0.435 (0.297 - 0.573) | 0.575 (0.416 - 0.734) | 0.442 (0.303 - 0.581) | 0.412 (0.279 - 0.546) | 0.552 (0.397 - 0.706) | 0.426 (0.29 - 0.563) | 0.442 (0.304 - 0.58) | 0.598 (0.437 - 0.759) | 0.467 (0.325 - 0.609) | 0.41 (0.277 - 0.542) | 0.568 (0.412 - 0.724) | 0.449 (0.31 - 0.587) | 0.506 (0.36 - 0.651) | 0.675 (0.507 - 0.844) | 0.524 (0.376 - 0.672) | 0.498 (0.354 - 0.641) | 0.674 (0.507 - 0.841) | 0.526 (0.379 - 0.673) |
| Medical-related injuries (consequences) | 0.24 (0.136 - 0.345) | 0.35 (0.224 - 0.476) | 0.281 (0.168 - 0.394) | 0.185 (0.095 - 0.274) | 0.281 (0.17 - 0.392) | 0.231 (0.131 - 0.332) | 0.259 (0.153 - 0.365) | 0.403 (0.271 - 0.535) | 0.335 (0.215 - 0.456) | 0.232 (0.131 - 0.333) | 0.361 (0.235 - 0.486) | 0.3 (0.185 - 0.414) | 0.303 (0.19 - 0.416) | 0.471 (0.33 - 0.611) | 0.392 (0.264 - 0.52) | 0.407 (0.278 - 0.536) | 0.614 (0.455 - 0.773) | 0.503 (0.359 - 0.646) |
| Other fractures | 1.158 (0.932 - 1.384) | 1.566 (1.303 - 1.829) | 1.22 (0.988 - 1.452) | 1.036 (0.823 - 1.248) | 1.412 (1.164 - 1.659) | 1.105 (0.886 - 1.324) | 1.118 (0.9 - 1.337) | 1.569 (1.31 - 1.828) | 1.243 (1.013 - 1.474) | 1.18 (0.957 - 1.404) | 1.645 (1.381 - 1.91) | 1.304 (1.068 - 1.539) | 1.143 (0.923 - 1.362) | 1.588 (1.329 - 1.846) | 1.252 (1.023 - 1.482) | 1.085 (0.871 - 1.298) | 1.556 (1.301 - 1.812) | 1.246 (1.017 - 1.474) |
| Poisoning - alcohol | 0.075 (0.019 - 0.131) | 0.09 (0.029 - 0.152) | 0.066 (0.013 - 0.119) | 0.063 (0.011 - 0.115) | 0.081 (0.022 - 0.14) | 0.061 (0.01 - 0.112) | 0.105 (0.038 - 0.172) | 0.123 (0.05 - 0.195) | 0.089 (0.027 - 0.151) | 0.105 (0.038 - 0.172) | 0.123 (0.051 - 0.196) | 0.089 (0.028 - 0.151) | 0.115 (0.045 - 0.184) | 0.143 (0.065 - 0.221) | 0.106 (0.039 - 0.173) | 0.097 (0.033 - 0.162) | 0.125 (0.052 - 0.198) | 0.094 (0.031 - 0.158) |
| Poisoning - opioid | 0.003 (-0.009 - 0.016) | 0.005 (-0.01 - 0.019) | 0.004 (-0.009 - 0.016) | 0.011 (-0.011 - 0.033) | 0.014 (-0.011 - 0.038) | 0.01 (-0.011 - 0.032) | 0.008 (-0.011 - 0.026) | 0.01 (-0.011 - 0.032) | 0.008 (-0.011 - 0.026) | 0.014 (-0.01 - 0.039) | 0.019 (-0.009 - 0.047) | 0.015 (-0.01 - 0.04) | 0.029 (-0.007 - 0.064) | 0.036 (-0.003 - 0.076) | 0.027 (-0.007 - 0.062) | 0.039 (-0.002 - 0.079) | 0.055 (0.007 - 0.103) | 0.043 (0.001 - 0.086) |
| Poisoning - other substances | 0.124 (0.051 - 0.197) | 0.147 (0.067 - 0.226) | 0.108 (0.039 - 0.176) | 0.134 (0.058 - 0.21) | 0.161 (0.077 - 0.245) | 0.118 (0.046 - 0.19) | 0.148 (0.068 - 0.228) | 0.183 (0.094 - 0.272) | 0.137 (0.06 - 0.213) | 0.188 (0.098 - 0.278) | 0.225 (0.127 - 0.324) | 0.166 (0.082 - 0.25) | 0.151 (0.071 - 0.232) | 0.2 (0.108 - 0.293) | 0.154 (0.073 - 0.235) | 0.306 (0.192 - 0.419) | 0.391 (0.263 - 0.519) | 0.296 (0.184 - 0.407) |
| Residual - injuries | 2.803 (2.451 - 3.156) | 3.721 (3.315 - 4.128) | 2.866 (2.509 - 3.223) | 2.431 (2.105 - 2.757) | 3.291 (2.912 - 3.67) | 2.559 (2.225 - 2.894) | 2.805 (2.458 - 3.152) | 3.741 (3.341 - 4.142) | 2.893 (2.541 - 3.246) | 2.809 (2.462 - 3.156) | 3.773 (3.372 - 4.175) | 2.93 (2.576 - 3.284) | 2.87 (2.524 - 3.217) | 3.835 (3.434 - 4.235) | 2.967 (2.615 - 3.32) | 3.005 (2.651 - 3.359) | 4.031 (3.621 - 4.441) | 3.121 (2.761 - 3.482) |
| Soft tissue injuries | 0.029 (-0.008 - 0.066) | 0.037 (-0.004 - 0.079) | 0.029 (-0.008 - 0.065) | 0.019 (-0.01 - 0.048) | 0.022 (-0.009 - 0.053) | 0.016 (-0.01 - 0.042) | 0.021 (-0.009 - 0.051) | 0.028 (-0.007 - 0.062) | 0.021 (-0.009 - 0.051) | 0.021 (-0.009 - 0.051) | 0.027 (-0.007 - 0.061) | 0.021 (-0.009 - 0.051) | 0.022 (-0.008 - 0.053) | 0.03 (-0.006 - 0.065) | 0.023 (-0.008 - 0.053) | 0.03 (-0.005 - 0.065) | 0.039 (-0.001 - 0.08) | 0.031 (-0.005 - 0.066) |
| Spinal cord injury | 0.09 (0.027 - 0.153) | 0.125 (0.051 - 0.199) | 0.097 (0.032 - 0.163) | 0.093 (0.03 - 0.157) | 0.127 (0.053 - 0.201) | 0.1 (0.034 - 0.165) | 0.098 (0.031 - 0.165) | 0.126 (0.05 - 0.201) | 0.095 (0.03 - 0.16) | 0.094 (0.032 - 0.157) | 0.125 (0.053 - 0.198) | 0.097 (0.033 - 0.161) | 0.13 (0.057 - 0.204) | 0.168 (0.084 - 0.252) | 0.128 (0.055 - 0.202) | 0.1 (0.036 - 0.164) | 0.129 (0.056 - 0.202) | 0.099 (0.035 - 0.162) |
| Tibia & ankle fracture | 0.074 (0.017 - 0.131) | 0.106 (0.037 - 0.175) | 0.085 (0.023 - 0.146) | 0.1 (0.034 - 0.167) | 0.143 (0.064 - 0.222) | 0.113 (0.043 - 0.184) | 0.107 (0.038 - 0.175) | 0.157 (0.074 - 0.24) | 0.127 (0.053 - 0.201) | 0.11 (0.042 - 0.179) | 0.154 (0.073 - 0.235) | 0.122 (0.05 - 0.194) | 0.101 (0.036 - 0.166) | 0.14 (0.064 - 0.217) | 0.11 (0.042 - 0.178) | 0.099 (0.035 - 0.162) | 0.142 (0.066 - 0.218) | 0.114 (0.046 - 0.182) |
| Traumatic brain injury | 1.164 (0.937 - 1.39) | 1.443 (1.191 - 1.696) | 1.08 (0.861 - 1.298) | 0.878 (0.681 - 1.074) | 1.137 (0.914 - 1.361) | 0.867 (0.672 - 1.062) | 0.983 (0.776 - 1.19) | 1.256 (1.022 - 1.49) | 0.953 (0.749 - 1.156) | 1.082 (0.867 - 1.297) | 1.366 (1.125 - 1.607) | 1.031 (0.821 - 1.241) | 0.994 (0.789 - 1.2) | 1.267 (1.036 - 1.499) | 0.96 (0.759 - 1.161) | 1.132 (0.915 - 1.35) | 1.425 (1.181 - 1.668) | 1.074 (0.863 - 1.286) |
| **External causes** | 27.528 (26.426 - 28.63) | 23.102 (22.093 - 24.111) | 31.123 (29.952 - 32.294) | 28.807 (27.688 - 29.927) | 23.531 (22.519 - 24.543) | 31.926 (30.747 - 33.105) | 28.646 (27.535 - 29.757) | 23.663 (22.653 - 24.673) | 31.774 (30.603 - 32.944) | 31.395 (30.237 - 32.552) | 25.939 (24.887 - 26.991) | 34.782 (33.564 - 36) | 31.495 (30.343 - 32.647) | 26.255 (25.203 - 27.307) | 35.081 (33.865 - 36.297) | 30.801 (29.669 - 31.932) | 25.345 (24.318 - 26.372) | 33.976 (32.787 - 35.165) |
| Accidental poisoning - alcohol | 0.028 (-0.007 - 0.062) | 0.023 (-0.009 - 0.054) | 0.031 (-0.006 - 0.068) | 0.022 (-0.009 - 0.053) | 0.019 (-0.009 - 0.047) | 0.026 (-0.007 - 0.059) | 0.118 (0.046 - 0.189) | 0.085 (0.025 - 0.146) | 0.122 (0.049 - 0.194) | 0.066 (0.013 - 0.119) | 0.047 (0.002 - 0.092) | 0.068 (0.015 - 0.122) | 0.112 (0.043 - 0.182) | 0.089 (0.027 - 0.151) | 0.125 (0.051 - 0.198) | 0.155 (0.073 - 0.236) | 0.112 (0.043 - 0.181) | 0.157 (0.075 - 0.238) |
| Accidental poisoning - drugs | 0.163 (0.079 - 0.247) | 0.152 (0.071 - 0.233) | 0.175 (0.088 - 0.261) | 0.136 (0.059 - 0.213) | 0.136 (0.059 - 0.213) | 0.143 (0.064 - 0.222) | 0.243 (0.14 - 0.345) | 0.205 (0.111 - 0.3) | 0.26 (0.154 - 0.366) | 0.226 (0.127 - 0.325) | 0.193 (0.102 - 0.285) | 0.256 (0.151 - 0.362) | 0.22 (0.123 - 0.317) | 0.199 (0.107 - 0.291) | 0.257 (0.152 - 0.362) | 0.418 (0.284 - 0.551) | 0.359 (0.236 - 0.483) | 0.468 (0.327 - 0.609) |
| Accidental threats to breathing | 0.976 (0.769 - 1.183) | 0.822 (0.632 - 1.012) | 1.027 (0.815 - 1.239) | 0.953 (0.749 - 1.157) | 0.725 (0.547 - 0.903) | 0.951 (0.747 - 1.154) | 1.063 (0.849 - 1.278) | 0.892 (0.696 - 1.089) | 1.106 (0.888 - 1.325) | 1.268 (1.035 - 1.5) | 1.06 (0.847 - 1.274) | 1.313 (1.076 - 1.55) | 1.075 (0.862 - 1.288) | 1.002 (0.796 - 1.207) | 1.143 (0.923 - 1.362) | 0.882 (0.692 - 1.073) | 0.797 (0.616 - 0.979) | 0.921 (0.726 - 1.115) |
| Drowning | 0.215 (0.118 - 0.313) | 0.179 (0.091 - 0.268) | 0.248 (0.144 - 0.352) | 0.176 (0.089 - 0.264) | 0.153 (0.071 - 0.236) | 0.198 (0.105 - 0.292) | 0.211 (0.115 - 0.307) | 0.166 (0.081 - 0.251) | 0.23 (0.13 - 0.329) | 0.218 (0.123 - 0.313) | 0.186 (0.098 - 0.274) | 0.254 (0.151 - 0.357) | 0.273 (0.166 - 0.379) | 0.223 (0.127 - 0.319) | 0.307 (0.194 - 0.421) | 0.246 (0.145 - 0.347) | 0.21 (0.116 - 0.303) | 0.286 (0.177 - 0.396) |
| Falls | 6.171 (5.647 - 6.695) | 4.926 (4.459 - 5.394) | 6.249 (5.722 - 6.776) | 6.524 (5.991 - 7.057) | 5.132 (4.659 - 5.605) | 6.515 (5.982 - 7.048) | 6.041 (5.531 - 6.552) | 4.765 (4.312 - 5.217) | 5.952 (5.445 - 6.458) | 6.655 (6.123 - 7.188) | 5.341 (4.864 - 5.818) | 6.673 (6.139 - 7.206) | 6.467 (5.946 - 6.988) | 5.098 (4.636 - 5.561) | 6.421 (5.902 - 6.94) | 6.809 (6.279 - 7.339) | 5.373 (4.901 - 5.844) | 6.784 (6.255 - 7.313) |
| Homicide & violence | 0.26 (0.153 - 0.366) | 0.251 (0.147 - 0.356) | 0.338 (0.216 - 0.459) | 0.216 (0.119 - 0.313) | 0.206 (0.112 - 0.301) | 0.276 (0.167 - 0.385) | 0.187 (0.097 - 0.277) | 0.169 (0.084 - 0.255) | 0.228 (0.129 - 0.328) | 0.195 (0.103 - 0.287) | 0.186 (0.096 - 0.275) | 0.25 (0.147 - 0.354) | 0.303 (0.191 - 0.416) | 0.275 (0.168 - 0.382) | 0.374 (0.249 - 0.498) | 0.278 (0.169 - 0.386) | 0.265 (0.159 - 0.371) | 0.355 (0.232 - 0.477) |
| Medical-related injuries (external) | 1.216 (0.983 - 1.449) | 0.619 (0.453 - 0.785) | 0.951 (0.745 - 1.156) | 1.161 (0.934 - 1.387) | 0.589 (0.428 - 0.751) | 0.894 (0.695 - 1.093) | 1.863 (1.579 - 2.147) | 0.967 (0.763 - 1.172) | 1.401 (1.155 - 1.647) | 2.084 (1.784 - 2.384) | 1.13 (0.909 - 1.351) | 1.61 (1.346 - 1.873) | 2.289 (1.98 - 2.599) | 1.244 (1.016 - 1.473) | 1.75 (1.479 - 2.021) | 2.155 (1.857 - 2.453) | 1.217 (0.993 - 1.441) | 1.681 (1.418 - 1.944) |
| RTI - cyclists | 0.196 (0.102 - 0.29) | 0.162 (0.077 - 0.248) | 0.225 (0.124 - 0.326) | 0.189 (0.099 - 0.28) | 0.151 (0.07 - 0.232) | 0.208 (0.113 - 0.303) | 0.208 (0.113 - 0.303) | 0.173 (0.087 - 0.26) | 0.236 (0.135 - 0.337) | 0.182 (0.095 - 0.269) | 0.171 (0.087 - 0.254) | 0.227 (0.131 - 0.324) | 0.284 (0.175 - 0.393) | 0.248 (0.146 - 0.349) | 0.337 (0.218 - 0.455) | 0.217 (0.123 - 0.312) | 0.191 (0.102 - 0.279) | 0.249 (0.147 - 0.35) |
| RTI - motor vehicle occupants | 0.657 (0.488 - 0.825) | 0.552 (0.398 - 0.707) | 0.753 (0.573 - 0.934) | 0.838 (0.647 - 1.029) | 0.757 (0.575 - 0.938) | 1.007 (0.798 - 1.216) | 0.834 (0.643 - 1.025) | 0.737 (0.558 - 0.917) | 0.987 (0.78 - 1.194) | 0.817 (0.63 - 1.005) | 0.714 (0.539 - 0.89) | 0.957 (0.753 - 1.16) | 0.721 (0.546 - 0.896) | 0.657 (0.489 - 0.824) | 0.885 (0.691 - 1.079) | 0.613 (0.453 - 0.774) | 0.548 (0.395 - 0.7) | 0.737 (0.56 - 0.913) |
| RTI - motorcyclists | 0.295 (0.182 - 0.409) | 0.254 (0.149 - 0.359) | 0.349 (0.226 - 0.473) | 0.321 (0.204 - 0.438) | 0.288 (0.177 - 0.398) | 0.383 (0.255 - 0.511) | 0.359 (0.235 - 0.483) | 0.32 (0.202 - 0.437) | 0.436 (0.299 - 0.573) | 0.273 (0.165 - 0.382) | 0.227 (0.128 - 0.325) | 0.311 (0.195 - 0.427) | 0.362 (0.237 - 0.486) | 0.329 (0.21 - 0.448) | 0.447 (0.308 - 0.585) | 0.359 (0.235 - 0.482) | 0.322 (0.205 - 0.439) | 0.433 (0.298 - 0.569) |
| RTI - pedestrians | 0.289 (0.174 - 0.404) | 0.239 (0.135 - 0.344) | 0.334 (0.21 - 0.457) | 0.201 (0.108 - 0.295) | 0.172 (0.086 - 0.258) | 0.236 (0.135 - 0.337) | 0.195 (0.104 - 0.286) | 0.178 (0.091 - 0.266) | 0.241 (0.139 - 0.343) | 0.218 (0.121 - 0.316) | 0.184 (0.094 - 0.274) | 0.255 (0.149 - 0.36) | 0.274 (0.167 - 0.381) | 0.238 (0.139 - 0.338) | 0.326 (0.21 - 0.443) | 0.192 (0.103 - 0.281) | 0.17 (0.086 - 0.254) | 0.232 (0.134 - 0.33) |
| Residual - external causes | 11.336 (10.629 - 12.043) | 8.438 (7.829 - 9.048) | 11.485 (10.774 - 12.196) | 12.587 (11.847 - 13.327) | 8.666 (8.053 - 9.28) | 12.093 (11.368 - 12.818) | 11.505 (10.801 - 12.209) | 8.34 (7.741 - 8.94) | 11.527 (10.822 - 12.232) | 12.45 (11.722 - 13.179) | 8.897 (8.281 - 9.513) | 12.308 (11.584 - 13.032) | 12.171 (11.455 - 12.888) | 8.772 (8.163 - 9.381) | 12.117 (11.401 - 12.833) | 11.656 (10.96 - 12.352) | 8.12 (7.539 - 8.702) | 11.231 (10.547 - 11.915) |
| Suicide | 6.367 (5.839 - 6.895) | 5.719 (5.219 - 6.219) | 7.75 (7.168 - 8.332) | 6.245 (5.724 - 6.766) | 5.609 (5.116 - 6.103) | 7.506 (6.936 - 8.077) | 6.212 (5.695 - 6.729) | 5.498 (5.012 - 5.984) | 7.461 (6.894 - 8.027) | 7.122 (6.571 - 7.673) | 6.348 (5.827 - 6.868) | 8.606 (8 - 9.213) | 7.209 (6.657 - 7.76) | 6.474 (5.952 - 6.997) | 8.782 (8.173 - 9.39) | 7.277 (6.725 - 7.829) | 6.483 (5.962 - 7.005) | 8.803 (8.196 - 9.41) |
| **Factors influencing health status and contact with health services** | 15.678 (14.84 - 16.516) | 20.75 (19.786 - 21.714) | 15.85 (15.007 - 16.692) | 16.835 (15.976 - 17.694) | 22.421 (21.43 - 23.412) | 17.184 (16.317 - 18.052) | 16.987 (16.129 - 17.845) | 22.762 (21.769 - 23.755) | 17.506 (16.635 - 18.376) | 17.405 (16.543 - 18.266) | 23.296 (22.299 - 24.293) | 17.918 (17.044 - 18.792) | 15.354 (14.551 - 16.157) | 20.652 (19.721 - 21.583) | 15.91 (15.093 - 16.728) | 15.433 (14.634 - 16.231) | 20.823 (19.896 - 21.75) | 16.069 (15.255 - 16.884) |
| Factors influencing health status and contact with health services | 9.369 (8.722 - 10.017) | 14.316 (13.515 - 15.116) | 11.761 (11.035 - 12.486) | 10.062 (9.398 - 10.725) | 15.452 (14.629 - 16.275) | 12.732 (11.985 - 13.479) | 10.193 (9.528 - 10.857) | 15.688 (14.864 - 16.513) | 12.95 (12.201 - 13.699) | 10.448 (9.781 - 11.115) | 16.088 (15.259 - 16.916) | 13.285 (12.532 - 14.037) | 9.31 (8.685 - 9.935) | 14.302 (13.527 - 15.077) | 11.805 (11.101 - 12.509) | 9.396 (8.773 - 10.019) | 14.465 (13.692 - 15.237) | 11.952 (11.249 - 12.654) |
| Higher order factors in bold. ASR = Age-standardised rate; main = values from the main analysis (UC = 50%, OC = 50%); equal = all causes weighted the same; double = UC is weighted double than a OC in the respective certificate | | | | | | | | | | | | | | | | | | |
| Note that values of higher order and lower order factors can differ even if only one category is present. This is due to exclusion of duplicate causes | | | | | | | | | | | | | | | | | | |

##### *Stringent exclusion criteria*

More stringent exclusion criteria led to the following exclusions:

There was a total of 538,525 death certificates in Austria for the years 2019 to 2024 (2019: 83,386; 2024: 88,486), with 2,502,281 causes (ICDs) total (2019: 369,844; 2024: 424,707). We excluded a total of 720,448 (2019: 110,071, 2024: 124,308) mentions of intermediate or immediate causes.^22^ This included the removal of 53,759 (2019: 9,114; 2024: 9,233) mentions as a UC, which led to the removal of those death certificates and indirectly also OCs that were not intermediate or immediate causes. We additionally excluded 195,431 (2019: 27,390; 2024: 33,999) ill-defined causes, also excluding 20,617 (2019: 2,884; 2024: 3,389) death certificates where ill-defined causes were stated as the UC (see lists below for definitions of intermediate, immediate and ill-defined causes). Note that some intermediate/immediate causes overlap with ill-defined causes, which is why the ill-defined exclusion here is less than in the main analysis. We also excluded 9,101 (2019: 1,306; 2024: 1,761) mentions of injuries in OC, due to co-occurrence with an external cause. We also removed all duplicate causes that were created when mapping ICD codes to the causes list. The final dataset for this sensitivity analysis had 464,148 certificates (2019: 71,388; 2024: 75,864) with overall 1,293,260 causes (2019: 190,299; 2024: 214,711).

The allocation to high-level causes led to further exclusion of OCs due to multiple causes being allocated to the same high-level cause. After excluding duplicates, the final higher-level analysis had a total of 984,153 causes (2019: 143,777; 2024: 163,402).

| Table S10: Sensitivity analysis with stringent exclusion: High-level causes and their ASRw, ASRuc, UC mentions and ASRw/ASRuc ratio for 2019 and 2024 | | | | | |
| --- | --- | --- | --- | --- | --- |
| High-level cause | year | ASRw per 100k persons | ASRuc per 100k persons | UC mentions | ratio ASRw to ASRuc |
| Blood diseases | 2019 | 3.94 (3.52 - 4.36) | 2.28 (1.97 - 2.6) | 202 | 1.73 (1.45 - 2.05) |
| Blood diseases | 2020-2023 avg | 4.64 (4.2 - 5.09) | 2.51 (2.18 - 2.83) | 227 | 1.93 (1.63 - 2.29) |
| Blood diseases | 2024 | 4.38 (3.95 - 4.8) | 2.03 (1.74 - 2.32) | 191 | 2.16 (1.82 - 2.57) |
| Cardiovascular diseases | 2019 | 222.11 (218.96 - 225.26) | 304.43 (300.75 - 308.12) | 26547 | 0.73 (0.72 - 0.74) |
| Cardiovascular diseases | 2020-2023 avg | 225.35 (222.25 - 228.46) | 288.88 (285.37 - 292.4) | 26191 | 0.78 (0.77 - 0.79) |
| Cardiovascular diseases | 2024 | 202.83 (199.94 - 205.73) | 260.4 (257.13 - 263.68) | 24564 | 0.78 (0.76 - 0.79) |
| Congenital conditions | 2019 | 1.94 (1.64 - 2.23) | 2.93 (2.57 - 3.28) | 260 | 0.66 (0.55 - 0.8) |
| Congenital conditions | 2020-2023 avg | 1.96 (1.67 - 2.25) | 2.87 (2.52 - 3.22) | 260 | 0.68 (0.56 - 0.83) |
| Congenital conditions | 2024 | 2.18 (1.87 - 2.49) | 3.09 (2.72 - 3.45) | 276 | 0.71 (0.59 - 0.85) |
| Digestive diseases | 2019 | 27.75 (26.64 - 28.87) | 24.46 (23.41 - 25.5) | 2130 | 1.13 (1.07 - 1.2) |
| Digestive diseases | 2020-2023 avg | 27.28 (26.2 - 28.37) | 23.63 (22.62 - 24.64) | 2138 | 1.16 (1.09 - 1.22) |
| Digestive diseases | 2024 | 26.07 (25.03 - 27.11) | 23.14 (22.16 - 24.12) | 2176 | 1.13 (1.06 - 1.19) |
| Endocrine disorders | 2019 | 55.17 (53.6 - 56.75) | 40.31 (38.96 - 41.66) | 3488 | 1.37 (1.31 - 1.43) |
| Endocrine disorders | 2020-2023 avg | 62.98 (61.34 - 64.63) | 48.64 (47.19 - 50.08) | 4401 | 1.3 (1.25 - 1.35) |
| Endocrine disorders | 2024 | 59.45 (57.89 - 61.02) | 46.79 (45.4 - 48.18) | 4420 | 1.27 (1.22 - 1.32) |
| External causes | 2019 | 27.91 (26.8 - 29.02) | 52.48 (50.96 - 54) | 4614 | 0.53 (0.51 - 0.56) |
| External causes | 2020-2023 avg | 30.55 (29.41 - 31.7) | 56.85 (55.29 - 58.41) | 5144 | 0.54 (0.51 - 0.56) |
| External causes | 2024 | 31.31 (30.17 - 32.46) | 57.92 (56.37 - 59.47) | 5410 | 0.54 (0.52 - 0.57) |
| Factors influencing health status and contact with health services | 2019 | 17.83 (16.93 - 18.72) | 0 (0 - 0) | 0 |  |
| Factors influencing health status and contact with health services | 2020-2023 avg | 18.99 (18.09 - 19.89) | 0 (0 - 0) | 0 |  |
| Factors influencing health status and contact with health services | 2024 | 17.72 (16.86 - 18.57) | 0 (0 - 0) | 0 |  |
| Genitourinary diseases | 2019 | 4.28 (3.84 - 4.72) | 2.13 (1.82 - 2.44) | 186 | 2.01 (1.69 - 2.4) |
| Genitourinary diseases | 2020-2023 avg | 4.84 (4.39 - 5.3) | 2.43 (2.11 - 2.76) | 220 | 1.99 (1.69 - 2.35) |
| Genitourinary diseases | 2024 | 4.67 (4.23 - 5.11) | 2.46 (2.14 - 2.78) | 231 | 1.89 (1.61 - 2.22) |
| Hearing and vision diseases | 2019 | 0.64 (0.47 - 0.81) | 0.01 (-0.01 - 0.04) | 1 | 53.12 (7.35 - 383.86) |
| Hearing and vision diseases | 2020-2023 avg | 0.77 (0.59 - 0.95) | 0.01 (-0.01 - 0.03) | 1 | 55.97 (9.11 - 362.83) |
| Hearing and vision diseases | 2024 | 0.78 (0.6 - 0.96) | 0.03 (0 - 0.06) | 3 | 28.25 (8.8 - 90.68) |
| Infectious diseases | 2019 | 11.1 (10.4 - 11.81) | 8.31 (7.7 - 8.92) | 730 | 1.34 (1.21 - 1.47) |
| Infectious diseases | 2020-2023 avg | 17.7 (16.83 - 18.57) | 8.1 (7.51 - 8.69) | 734 | 2.2 (2.02 - 2.41) |
| Infectious diseases | 2024 | 16.86 (16.03 - 17.7) | 9.11 (8.5 - 9.73) | 854 | 1.85 (1.7 - 2.01) |
| Injuries | 2019 | 6.11 (5.59 - 6.63) | 0 (0 - 0) | 0 |  |
| Injuries | 2020-2023 avg | 5.91 (5.4 - 6.41) | 0 (0 - 0) | 0 |  |
| Injuries | 2024 | 6.44 (5.92 - 6.95) | 0 (0 - 0) | 0 |  |
| Maternal conditions | 2019 | 0.1 (0.03 - 0.16) | 0.06 (0.01 - 0.11) | 5 | 1.72 (0.57 - 5.16) |
| Maternal conditions | 2020-2023 avg | 0.04 (0 - 0.08) | 0.03 (-0.01 - 0.06) | 2 | 1.47 (0.28 - 7.71) |
| Maternal conditions | 2024 | 0.1 (0.03 - 0.17) | 0.02 (-0.01 - 0.05) | 2 | 4.58 (0.98 - 21.4) |
| Mental & behavioural disorders | 2019 | 16.85 (15.99 - 17.71) | 14.79 (13.98 - 15.6) | 1295 | 1.14 (1.06 - 1.23) |
| Mental & behavioural disorders | 2020-2023 avg | 20.3 (19.36 - 21.23) | 17.37 (16.51 - 18.23) | 1572 | 1.17 (1.09 - 1.25) |
| Mental & behavioural disorders | 2024 | 21.33 (20.39 - 22.27) | 18.45 (17.58 - 19.33) | 1726 | 1.16 (1.08 - 1.23) |
| Musculoskeletal conditions | 2019 | 9.38 (8.73 - 10.02) | 4.03 (3.61 - 4.46) | 352 | 2.32 (2.05 - 2.64) |
| Musculoskeletal conditions | 2020-2023 avg | 9.96 (9.31 - 10.62) | 3.98 (3.56 - 4.39) | 362 | 2.53 (2.23 - 2.86) |
| Musculoskeletal conditions | 2024 | 10.2 (9.55 - 10.85) | 4.25 (3.83 - 4.67) | 401 | 2.4 (2.13 - 2.7) |
| Neoplasms | 2019 | 131.96 (129.53 - 134.39) | 245.08 (241.77 - 248.39) | 21325 | 0.54 (0.53 - 0.55) |
| Neoplasms | 2020-2023 avg | 131.42 (129.05 - 133.8) | 241.13 (237.91 - 244.35) | 21842 | 0.55 (0.53 - 0.56) |
| Neoplasms | 2024 | 127.56 (125.27 - 129.86) | 237.32 (234.19 - 240.44) | 22431 | 0.54 (0.53 - 0.55) |
| Nervous system diseases | 2019 | 69.64 (67.88 - 71.4) | 68.02 (66.28 - 69.77) | 5929 | 1.02 (0.99 - 1.06) |
| Nervous system diseases | 2020-2023 avg | 73.54 (71.76 - 75.31) | 69.53 (67.8 - 71.25) | 6314 | 1.06 (1.02 - 1.1) |
| Nervous system diseases | 2024 | 71.92 (70.19 - 73.64) | 77.57 (75.77 - 79.36) | 7312 | 0.93 (0.9 - 0.96) |
| Perinatal conditions | 2019 | 1.09 (0.88 - 1.3) | 1.64 (1.38 - 1.9) | 150 | 0.66 (0.52 - 0.86) |
| Perinatal conditions | 2020-2023 avg | 1.03 (0.82 - 1.24) | 1.53 (1.27 - 1.78) | 137 | 0.68 (0.52 - 0.88) |
| Perinatal conditions | 2024 | 1.17 (0.94 - 1.4) | 1.6 (1.33 - 1.87) | 132 | 0.73 (0.56 - 0.95) |
| Respiratory diseases | 2019 | 48.83 (47.35 - 50.31) | 47.6 (46.13 - 49.06) | 4134 | 1.03 (0.98 - 1.07) |
| Respiratory diseases | 2020-2023 avg | 80.37 (78.51 - 82.22) | 106.59 (104.46 - 108.73) | 9636 | 0.76 (0.74 - 0.79) |
| Respiratory diseases | 2024 | 55.86 (54.34 - 57.38) | 60.18 (58.61 - 61.76) | 5684 | 0.93 (0.89 - 0.96) |
| Skin diseases | 2019 | 1.93 (1.64 - 2.23) | 0.47 (0.32 - 0.62) | 40 | 4.11 (2.91 - 5.82) |
| Skin diseases | 2020-2023 avg | 1.97 (1.68 - 2.26) | 0.48 (0.34 - 0.62) | 43 | 4.3 (3.04 - 6.08) |
| Skin diseases | 2024 | 1.98 (1.69 - 2.27) | 0.54 (0.39 - 0.69) | 51 | 3.67 (2.69 - 5.02) |

| Table S11: Sensitivity analysis: Top 10 causes most co-occurring with COVID-19 by year | | | | | |
| --- | --- | --- | --- | --- | --- |
| Rank | 2024 | 2023 | 2022 | 2021 | 2020 |
| 1 | Residual - infections | Residual - infections | Residual - infections | Residual - infections | Residual - infections |
| 2 | Hypertensive diseases | Hypertensive diseases | Hypertensive diseases | Hypertensive diseases | Hypertensive diseases |
| 3 | Ischaemic heart disease | Ischaemic heart disease | Ischaemic heart disease | Ischaemic heart disease | Ischaemic heart disease |
| 4 | Dementia & Alzheimer disease | Diabetes mellitus | Dementia & Alzheimer disease | Dementia & Alzheimer disease | Dementia & Alzheimer disease |
| 5 | Diabetes mellitus | Dementia & Alzheimer disease | Diabetes mellitus | Diabetes mellitus | Diabetes mellitus |
| 6 | Artery diseases | Artery diseases | Other heart diseases | Other heart diseases | Other heart diseases |
| 7 | Other heart diseases | Other heart diseases | Artery diseases | Artery diseases | Cerebrovascular disease |
| 8 | Pneumonia | COPD | Cerebrovascular disease | Cerebrovascular disease | COPD |
| 9 | Cerebrovascular disease | Cerebrovascular disease | COPD | COPD | Artery diseases |
| 10 | COPD | Factors influencing health status and contact with health services | Factors influencing health status and contact with health services | Factors influencing health status and contact with health services | Factors influencing health status and contact with health services |

| Table S12: Sensitivity analysis: Frequencies of causes most co-occurring with COVID-19 (in 2024) by year, and percentage where COVID-19 was determined the underlying cause. | | | | | |
| --- | --- | --- | --- | --- | --- |
| COVID as underlying cause | 2024 | 2023 | 2022 | 2021 | 2020 |
|  | frequency co-occurring (COVID-19 as UC percentage) | | | | |
| Residual - infections | 1,009 (71.6%) | 1,305 (78.5%) | 2,595 (82.1%) | 2,710 (92%) | 2,128 (97.9%) |
| Hypertensive diseases | 529 (72.6%) | 1,038 (78.4%) | 2,234 (85%) | 2,515 (93.2%) | 1,986 (94.1%) |
| Ischaemic heart disease | 287 (79.1%) | 614 (86%) | 1,300 (87.5%) | 1,470 (95%) | 1,192 (97.1%) |
| Dementia & Alzheimer disease | 286 (86%) | 635 (84.3%) | 1,429 (92.9%) | 1,092 (93.6%) | 1,219 (94%) |
| Diabetes mellitus | 287 (74.9%) | 510 (79.6%) | 1,218 (87.4%) | 1,497 (93.3%) | 1,179 (94.8%) |
| Other heart diseases | 226 (68.6%) | 510 (68.4%) | 1,012 (83.1%) | 868 (88.4%) | 756 (89%) |
| Cerebrovascular disease | 190 (73.7%) | 392 (80.9%) | 841 (88.2%) | 630 (95.6%) | 616 (94.6%) |
| COPD | 149 (69.1%) | 389 (86.6%) | 759 (89.3%) | 817 (93.3%) | 621 (96.9%) |
| Artery diseases | 181 (67.4%) | 444 (71.4%) | 933 (80.2%) | 1,015 (89.5%) | 825 (90.8%) |
| Factors influencing health status and contact with health services | 159 (59.7%) | 293 (66.6%) | 583 (76%) | 671 (89.4%) | 434 (91.2%) |

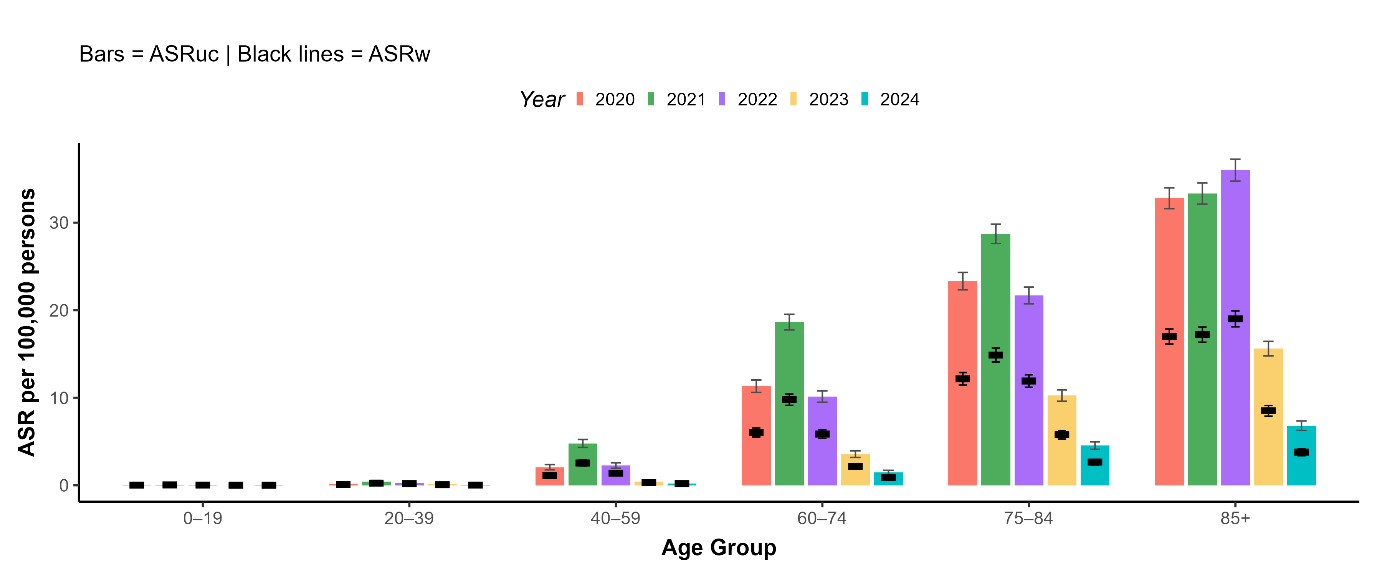
Figure S2: Version of Figure 2 using more stringent exclusion criteria. Stratified age standardised unweighted and weighted COVID-19 death counts from 2020 to 2024. Bars indicate 95% confidence intervals. ASRuc = age standardised underlying causes; ASRw = age standardised weighted counts

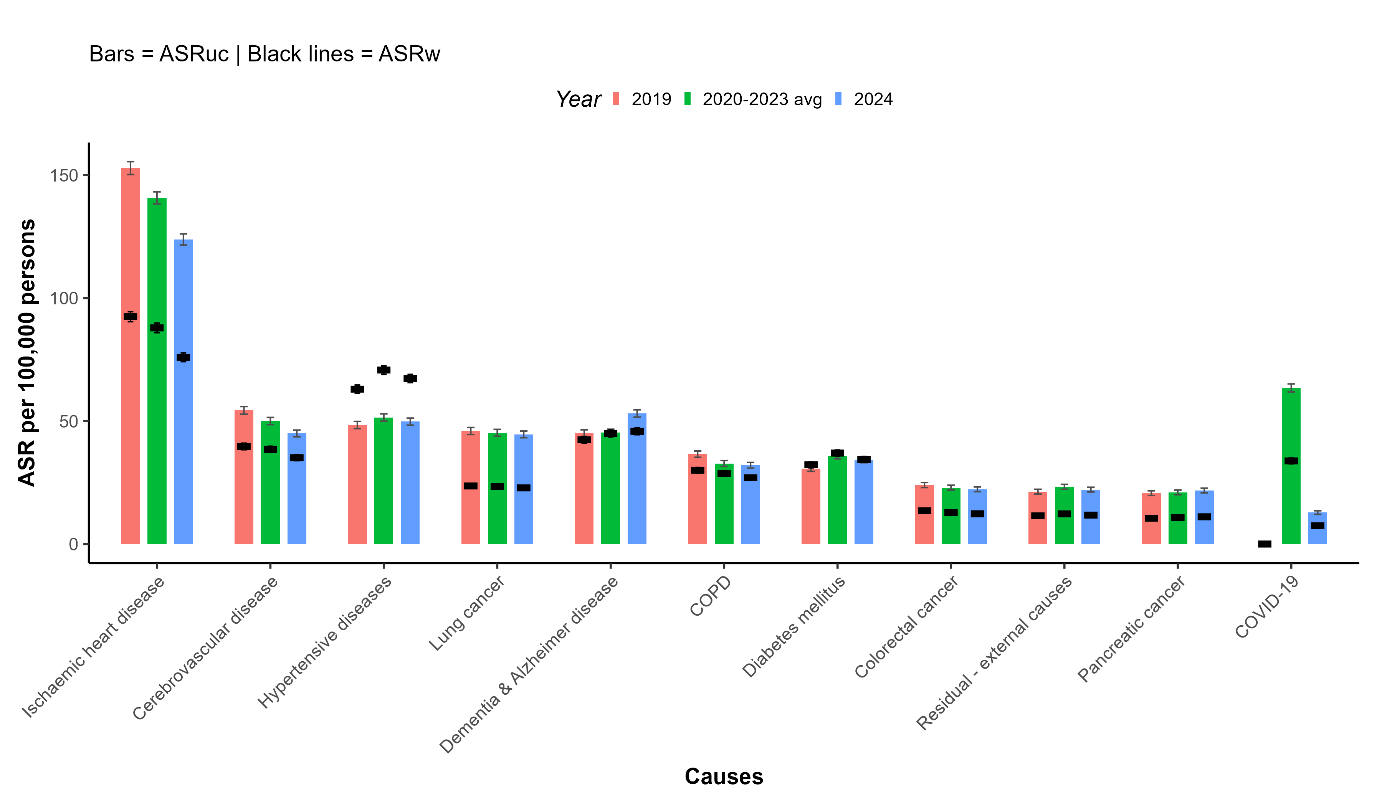
Figure S3: Version of Figure 3 using more stringent exclusion criteria. Age standardised weighted death counts of the top 10 causes of death (as of 2019) and COVID-19 for 2019, 2020-2023 and 2024. Bars indicate 95% confidence intervals. ASRuc = age standardised underlying causes; ASRw = age standardised weighted counts

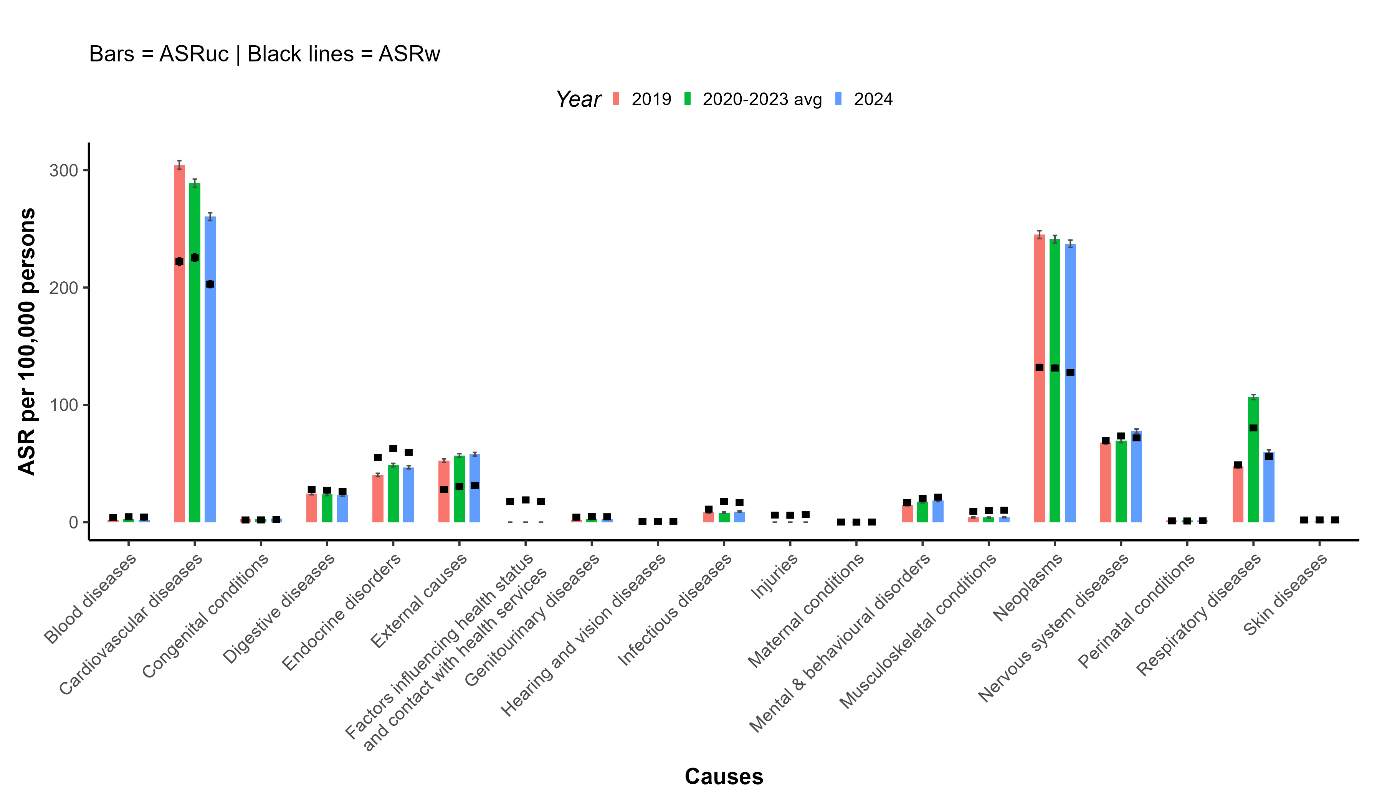

Figure S4: Version of Figure 4 using more stringent exclusion criteria. Age standardised underlying cause (ASRuc) and age standardised weighted counts (ASRw) of the 19 high-level factors for 2019, 2020-2023 and 2024. Bars indicate 95% confidence intervals. COVID-19 is included in “Respiratory diseases”.

#### **List of immediate and intermediate causes**

As listed by Flagg et al. (2021)^4^

1. Sepsis, unspecified (A41.9)
2. Gas gangrene (A48.0)
3. Toxic shock syndrome (A48.3)
4. Secondary and unspecified malignant neoplasm of lymph nodes (C77)
5. Secondary malignant neoplasm of respiratory and digestive organs (C78)
6. Secondary malignant neoplasm of other sites (C79)
7. Iron deficiency anaemia secondary to blood loss (chronic) (D50.0)
8. Acute posthaemorrhagic anaemia (D62)
9. Secondary sideroblastic anaemia due to disease (D64.1)
10. Anaemia, unspecified (D64.9)
11. Disseminated intravascular coagulation [defibrination syndrome] (D65)
12. Secondary thrombocytopenia (D69.5)
13. Haemorrhagic condition, unspecified (D69.9)
14. Secondary polycythaemia (D75.1)
15. Postinfectious hypothyroidism (E03.3)
16. Other hypoglycaemia (E16.1)
17. Hypoglycaemia, unspecified (E16.2)
18. Secondary hyperparathyroidism, not elsewhere classified (E21.1)
19. Secondary hyperaldosteronism (E26.1)
20. Secondary lactase deficiency (E73.1)
21. Secondary systemic amyloidosis (E85.3)
22. Volume depletion (E86)
23. Other disorders of fluid, electrolyte and acid-base balance (E87)
24. Postencephalitic syndrome (F07.1)
25. Postconcussional syndrome (F07.2)
26. Obstructive hydrocephalus (G91.1)
27. Post-traumatic hydrocephalus, unspecified (G91.3)
28. Other hydrocephalus (G91.8)
29. Hydrocephalus, unspecified (G91.9)
30. Toxic encephalopathy (G92)
31. Anoxic brain damage, not elsewhere classified (G93.1)
32. Postviral fatigue syndrome (G93.3)
33. Encephalopathy, unspecified (G93.4)
34. Compression of brain (G93.5)
35. Cerebral oedema (G93.6)
36. Secondary hypertension (I15)
37. Pulmonary embolism (I26)
38. Other secondary pulmonary hypertension (I27.2)
39. Cardiomyopathy, unspecified (I42.9)
40. Paroxysmal tachycardia (I47)
41. Atrial fibrillation and flutter (I48)
42. Other cardiac arrhythmias (I49)
43. Heart failure (I50)
44. Phlebitis and thrombophlebitis (I80)
45. Portal vein thrombosis (I81)
46. Other venous embolism and thrombosis (I82)
47. Other hypotension (I95.8)
48. Hypotension, unspecified (I95.9)
49. Pneumonia, organism unspecified (J18)
50. Adult respiratory distress syndrome (J80)
51. Pulmonary oedema (J81)
52. Pyothorax (J86)
53. Pleural effusion, not elsewhere classified (J90)
54. Other pneumothorax (J93.8)
55. Pneumothorax, unspecified (J93.9)
56. Other pleural conditions (J94)
57. Pulmonary collapse (J98.1)
58. Compensatory emphysema (J98.3)
59. Noninfective gastroenteritis and colitis, unspecified (K52.9)
60. Peritonitis (K65)
61. Other disorders of peritoneum (K66)
62. Hepatic failure, not elsewhere classified (K72)
63. Secondary biliary cirrhosis (K74.4)
64. Abscess of liver (K75.0)
65. Fatty (change of) liver, not elsewhere classified (K76.0)
66. Chronic passive congestion of liver (K76.1)
67. Central haemorrhagic necrosis of liver (K76.2)
68. Perforation of gallbladder (K82.2)
69. Haematemesis (K92.0)
70. Melaena (K92.1)
71. Gastrointestinal haemorrhage, unspecified (K92.2)
72. Cutaneous abscess, furuncle and carbuncle (L02)
73. Cellulitis (L03)
74. Decubitus ulcer and pressure area (L89)
75. Reactive arthropathies (M02)
76. Other secondary gout (M10.4)
77. Secondary multiple arthrosis (M15.3)
78. Post-traumatic coxarthrosis, bilateral (M16.4)
79. Other post-traumatic coxarthrosis (M16.5)
80. Other secondary coxarthrosis, bilateral (M16.6)
81. Other secondary coxarthrosis (M16.7)
82. Post-traumatic gonarthrosis, bilateral (M17.2)
83. Other post-traumatic gonarthrosis (M17.3)
84. Other secondary gonarthrosis, bilateral (M17.4)
85. Other secondary gonarthrosis (M17.5)
86. Post-traumatic arthrosis of first carpometacarpal joints, bilateral (M18.2)
87. Other post-traumatic arthrosis of first carpometacarpal joint (M18.3)
88. Other secondary arthrosis of first carpometacarpal joints, bilateral (M18.4)
89. Other secondary arthrosis of first carpometacarpal joint (M18.5)
90. Post-traumatic arthrosis of other joints (M19.1)
91. Other secondary arthrosis (M19.2)
92. Other secondary scoliosis (M41.5)
93. Postoophorectomy osteoporosis with pathological fracture (M80.1)
94. Postsurgical malabsorption osteoporosis with pathological fracture (M80.3)
95. Postoophorectomy osteoporosis (M81.1)
96. Postsurgical malabsorption osteoporosis (M81.3)
97. Osteomyelitis (M86)
98. Other secondary osteonecrosis (M87.3)
99. Acute renal failure, chronic kidney disease, and unspecified kidney failure (N17–N19)
100. Post-traumatic urethral stricture (N35.0)
101. Postinfective urethral stricture, not elsewhere classified (N35.1)
102. Urinary tract infection, site not specified (N39.0)
103. Pre-existing secondary hypertension complicating pregnancy, childbirth and the puerperium (O10.4)
104. Secondary uterine inertia (O62.1)
105. Respiratory distress of newborn (P22)
106. Fetal blood loss, unspecified (P50.9)
107. Neonatal haemorrhage, unspecified (P54.9)
108. Convulsions of newborn (P90)

#### **List of Causes**

Higher order causes in bold

| **Infectious diseases** |  |
| --- | --- |
| Intestinal infections | A00–A09 |
| Tuberculosis | A15–A19, B90 |
| Septicaemia | A40–A41 |
| Viral hepatitis | B15–B19, B942 |
| HIV disease | B20–B24 |
| Residual - infections | A20–A39, A42–A99, B00–B09, B25–B34, B35–B49, B50–B89, B91–B93, B940–B941, B948–B949, B95–B99 |
| **Neoplasms** |  |
| Oral cancers | C00–C14 |
| Oesophagus cancer | C15 |
| Stomach cancer | C16 |
| Colorectal cancer | C18–C21, C260 |
| Liver cancer | C22 |
| Gallbladder cancer | C23–C24 |
| Pancreatic cancer | C25 |
| Larynx cancer | C32 |
| Lung cancer | C33–C34 |
| Malignant melanoma – skin | C43 |
| Non-melanoma – skin | C44 |
| Mesothelioma | C45 |
| Breast cancer | C50 |
| Cervical cancer | C53 |
| Uterus cancer | C54–C55 |
| Ovarian cancer | C56 |
| Prostate cancer | C61 |
| Kidney cancer | C64 |
| Bladder cancer | C67 |
| Brain cancer | C71 |
| Thyroid cancer | C73 |
| Cancer unknown primary | C76, C80, C97 |
| Cancer secondary site | C77–C79 |
| Hodgkin lymphoma | C81 |
| Non-Hodgkin lymphomas | C82–C86 |
| Other blood cancers | C88, C90–C96, D45–D46, D471, D473–D475 |
| Residual – benign/in situ/uncertain neoplasms | D00–D44, D470, D472, D477–D479, D48 |
| Residual – malignant neoplasms | C17, C261, C268, C269, C30–C31, C37–C39, C40–C41, C46–C49, C51–C52, C57–C58, C60, C62–C63, C65–C66, C68–C70, C72, C74–C75 |
| **Blood diseases** |  |
| Anaemias | D50–D64 |
| Residual – blood diseases | D65–D89 |
| **Endocrine disorders** |  |
| Disorders of thyroid gland | E00–E07 |
| Diabetes mellitus | E10–E14 |
| Malnutrition | E40–E46 |
| Obesity | E66 |
| Amyloidosis | E85 |
| Dehydration disorders | E86–E87 |
| Metabolic disorders | E70–E84, E88–E90 |
| Residual – endocrine | E15–E16, E20–E243, E248–E35, E50–E65, E67–E68 |
| **Mental & behavioural disorders** |  |
| Alcohol-induced diseases | E244, F10, G312, G621, G721, I426, K292, K70, K852, K860 |
| Substance use disorders | F11–F19 |
| Schizophrenia | F20–F29 |
| Mood disorders | F30–F39 |
| Residual – mental/behavioural | F04–F09, F40–F99 |
| **Nervous system diseases** |  |
| Inflammatory diseases – CNS | G00–G09 |
| Systemic atrophies – CNS | G10–G14 |
| Parkinson disease | G20 |
| Dementia & Alzheimer disease | F00–F03, G30, G310, G318 |
| Multiple sclerosis | G35 |
| Epilepsy | G40, G41 |
| Cerebral palsy | G80 |
| Residual – nervous system | G21–G26, G311, G319, G32, G36–G37, G43–G44, G47, G50–G620, G622–G64, G70–G720, G722–G73, G81–G83, G90–G99 |
| **Hearing and vision diseases** | H00–H95 |
| **Cardiovascular diseases** |  |
| Chronic rheumatic heart diseases | I05–I09 |
| Hypertensive diseases | I10–I15 |
| Ischaemic heart disease | I20–I25 |
| Pulmonary heart diseases | I26–I28 |
| Non-rheumatic valve disorders | I34–I36 |
| Atrial fibrillation | I48 |
| Heart failure (specified) | I500, I501 |
| Other heart diseases | I30–I33, I37–I39, I40–I41, I420–I425, I427–I45, I47, I49, I51–I52 |
| Cerebrovascular disease | I60–I69 |
| Artery diseases | I70–I79 |
| Phlebitis & thrombophlebitis | I80 |
| Transient cerebral ischaemic attack | G45–G46 |
| Residual – cardiovascular | I00–I02, I81–I89, I950–I958, I96–I98 |
| **Respiratory diseases** |  |
| Influenza | J09–J11 |
| Pneumonia | J12–J18 |
| Other ALRI | J20–J22 |
| COPD | J40–J44 |
| Asthma | J45–J46 |
| Bronchiectasis | J47 |
| Pneumonitis | J69 |
| Other interstitial respiratory diseases | J80–J84 |
| Other diseases of pleura | J90–J94 |
| COVID-19 | U071 |
| Residual – respiratory | J00–J06, J30–J39, J60–J68, J70, J85–J86, J95, J961–J968, J97–J99 |
| **Digestive diseases** |  |
| Diseases of oesophagus, stomach & duodenum | K20–K291, K293–K31 |
| Other diseases of intestines | K55–K64 |
| Diseases of peritoneum | K65–K67 |
| Cirrhosis of the liver | K74 |
| Other diseases of liver | K71–K73, K75–K77 |
| Disorders of gallbladder, biliary tract & pancreas | K80–K851, K853–K859, K861–K87 |
| Residual – digestive | K00–K14, K35–K38, K40–K46, K50–K52, K90–K93 |
| **Skin diseases** |  |
| Infections of skin | L00–L08 |
| Residual – skin diseases | L10–L99 |
| **Musculoskeletal conditions** |  |
| Infectious arthropathies | M00–M03 |
| Rheumatoid arthritis | M05–M06 |
| Osteoarthritis | M15–M19 |
| Systemic connective tissue disorders | M30–M36 |
| Osteopathies & chondropathies | M80–M94 |
| Residual – musculoskeletal | M07–M14, M20–M25, M40–M79, M95–M99 |
| **Genitourinary diseases** |  |
| Glomerular diseases | N00–N08 |
| Renal tubulo-interstitial diseases | N10–N16 |
| Renal failure | N17–N19 |
| Urolithiasis | N20–N23 |
| Hyperplasia of prostate | N40 |
| Residual – genitourinary | N25–N29, N30–N39, N41–N51, N60–N99 |
| **Maternal conditions** | O00–O99 |
| **Perinatal conditions (incl. SIDS)** | P00–P284, P286–P96, R95 |
| **Congenital conditions** | Q00–Q99 |
| **Ill-defined causes** | I46, I509, I959, I99, J960, J969, P285, R00–R99 (excl. R95), U00-U99 (excl. U071) |
| **Injuries** |  |
| Traumatic brain injury | S020, S021, S027, S029, S06, T902, T905 |
| Spinal cord injury | S140, S141, S147, S240, S241, S247, S340, S341, S347, T060, T061, T093, T903, T913 |
| Internal & crush injuries | S07, S17, S18, S224, S225, S25–S28, S297, S35–S37, S380, S381, S396, S397, S47, S57, S67, S77, S87, S97, T04, T065, T147, T914, T915 |
| Poisoning - other substances | T36–T39, T407–T409, T41–T50, T52–T65, T940, T941, T96, T97 |
| Poisoning – alcohol | T51 |
| Poisoning – opioid | T400–T406 |
| Hip fracture | S72, T931 |
| Tibia & ankle fracture | S82 |
| Humerus fracture | S422, S423, S424, S427 |
| Other fractures | S022–S026, S028, S12, S220–S223, S228, S229, S32, S420–S421, S428–S429, S497, S52, S597, S620–S628, S697, S820, S92, T02, T08, T10, T12, T142, T911, T912, T921, T922, T932 |
| Drowning/submersion injuries | T751 |
| Dislocations | S030–S033, S131–S133, S231–S232, S331–S333, S430–S433, S530, S531, S630–S632, S730, S830, S831, S930, S931, S933, T03, T092, T112, T132, T143 |
| Soft tissue injuries | S034–S035, S134–S136, S16, S230, S233–S235, S290, S335–S337, S390, S434–S437, S46, S532–S534, S56, S633–S637, S66, S731, S76, S832–S837, S86, S932, S934–S936, S96, T064, T095, T115, T135, T146 |
| Burns | T20–T32, T95 |
| Medical-related injuries (consequences) | T80–T88, T983 |
| Residual – injuries | S00, S01, S04, S05, S08–S11, S130, S142–S146, S15, S19, S20, S21, S242–S246, S298, S299, S30, S31, S330, S334, S342–S346, S348, S382, S383, S398, S399, S40, S41, S44, S45, S48, S498, S499, S50, S51, S54, S55, S58, S598, S599, S60, S61, S64, S65, S68, S698, S699, S70, S71, S74, S75, S78, S799, S80, S81, S84, S85, S88–S91, S94, S95, S98, S99, T00, T01, T05, T062, T063, T068, T07, T090, T091, T094, T096, T098, T099, T110, T111, T113, T114, T116, T118, T119, T130, T131, T133, T134, T136, T138, T139, T140, T141, T144, T145, T148, T149, T15–T19, T33–T35, T66–T75 (excl. T751), T78, T79, T900, T901, T904, T908, T909, T910, T918–T920, T924, T928–T930, T933, T934, T936, T938, T939, T980, T981, T982 T980–T982, |
| **External causes** |  |
| RTI – motorcyclists | V203 –V209, V213–V219, V223–V229, V233–V239, V243–V249, V253–V259, V263–V269, V273–V279, V283–V289, V294–V299 |
| RTI – vehicle occupants | V304–V309, V314–V319, V324–V329, V334–V339, V344–V349, V354–V359, V364–V369, V374–V379, V384–V389, V394–V399, V404–V409, V414–V419, V424–V429, V434–V439, V444–V449, V454–V459, V464–V469, V474–V479, V484–V489, V494–V499, V504–V509, V514–V519, V524–V529, V534–V539, V544–V549, V554–V559, V564–V569, V574–V579, V584–V589, V594–V599, V604–V609, V614–V619, V624–V629, V634–V639, V644–V649, V654–V659, V664–V669, V674–V679, V684–V689, V694–V699, V704–V709, V714–V719, V724–V729, V734–V739, V744–V749, V754–V759, V764–V769, V774–V779, V784–V789, V794–V799, V870–V879, V892, Y850 |
| RTI – cyclists | V103 –V109, V113–V119, V123–V129, V133–V139, V143–V149, V153–V159, V163–V169, V173–V179, V183–V189, V194–V199 |
| RTI – pedestrians | V011, V019, V021, V029, V031, V039, V041, V049, V051, V059, V061, V069, V092, V093, V099 |
| Accidental poisoning – alcohol | X45 |
| Accidental poisoning – drugs | X42–X44 |
| Falls | W00–W19 |
| Drowning | V90, V92, W65–W74 |
| Accidental threats to breathing | W75–W84 |
| Suicide | X60–X84, Y870 |
| Homicide & violence | X85–Y09, Y871 |
| Medical-related injuries (external) | Y40–Y84, Y88 |
| Residual – external causes | V010, V020, V030, V040, V050, V060, V090, V091, V100–V102, V110–V112, V120–V122, V130–V132, V140–V142, V150–V152, V160–V162, V170–V172, V180–V182, V190–V193, V200–V202, V210–V212, V220–V222, V230–V232, V240– V242, V250–V252, V260–V262, V270–V272, V280–V282, V290–V293, V300–V303, V310–V313, V320–V323, V330–V333, V340–V343, V350–V353, V360–V363, V370–V373, V380–V383, V390–V393, V400–V403, V410–V413, V420–V423, V430–V433, V440–V443, V450–V453, V460–V463, V470–V473, V480–V483, V490–V493, V500–V503, V510–V513, V520–V523, V530–V533, V540–V543, V550–V553, V560–V563, V570–V573, V580–V583, V590–V593, V600–V603, V610–V613, V620–V623, V630–V633, V640–V643, V650–V653, V660–V663, V670–V673, V680–V683, V690–V693, V700–V703, V710–V713, V720–V723, V730–V733, V740–V743, V750–V753, V760–V763, V770– V773, V780–V783, V790–V793, V80–V86, V88, V890, V891, V893, V899, V91, V93–V99, W20–W64, W85–W99, X00–X39, X40–X41, X46–X49, X50–X59, Y10–Y34, Y35–Y36, Y859, Y86, Y872, Y89, Y90–Y98 |
| **Factors influencing health status and contact with health services** | Z00-Z99 |
| Based on WHO and the Bishop et. al., with small adaptations (noted below)^3,5^ | |

#### **Adaptations to List of Causes in Bishop**^3^

- Included “Factors influencing health status and contact with health services – Z00-Z99” from WHO list
- Added all “U” ICDs expect for COVID-19 (U07.1) to “ill-defined”
- Added COVID-19 (U07.1) and to the higher order category “Respiratory diseases”
- Combined “Hypertension” and “Hypertensive disease” together to “Hypertensive disease” as originally categorized by WHO
- Excluded S028 from “Traumatic brain injury” as it is already present in “Other fractures”
- Added T78 and T79 to „Residual – injuries“ (previously unspecified)
